# Covid-19 in the Phase 3 Trial of mRNA-1273 During the Delta-variant Surge

**DOI:** 10.1101/2021.09.17.21263624

**Authors:** Lindsey R. Baden, Hana M. El Sahly, Brandon Essink, Dean Follmann, Kathleen M. Neuzil, Allison August, Heather Clouting, Gabrielle Fortier, Weiping Deng, Shu Han, Xiaoping Zhao, Brett Leav, Carla Talarico, Bethany Girard, Yamuna D. Paila, Joanne E. Tomassini, Florian Schödel, Rolando Pajon, Honghong Zhou, Rituparna Das, Jacqueline Miller

## Abstract

**Background:** Following emergency use authorization in December 2020, the Coronavirus Efficacy (COVE) trial was amended to an open-label phase, where participants were unblinded and those randomized to placebo were offered vaccination. Emergence of the delta variant of severe acute respiratory syndrome coronavirus 2 (SARS-CoV-2) has been associated with increased incidences of coronavirus disease 2019 (Covid-19) among unvaccinated and vaccinated persons. This exploratory analysis evaluated the incidence and genetic sequences of Covid-19 cases in the ongoing COVE trial during the open-label phase, with a focus on July-August 2021, when delta-variants surged in the US.

**Methods:** Covid-19 cases were identified in participants initially randomized to mRNA-1273 (vaccinated from July-December 2020) and those initially randomized to the placebo (vaccinated December 2020-April 2021) who received at least one dose and were SARS-CoV-2-negative at baseline in the modified-intent-to-treat population were analyzed. Included were Covid-19 cases occurring after 26-Mar-2021 with positive RT-PCR results in nasopharyngeal samples (central lab test) and reported Covid-19 symptoms. Genetic sequencing of Covid-19 cases was also performed.

**Results:** There were 14,746 participants in the earlier mRNA-1273 (mRNA-1273e) group and 11,431 in the later placebo-mRNA1273 (mRNA-1273p) group. Covid-19 cases increased from the start of the open-label phase to July-August 2021. During July and August, 162 Covid-19 cases occurred in the mRNA-1273e group and 88 in the mRNA-1273p group. Of the cases sequenced, 144/149 [97%]) in the mRNA-1273 and 86/88 (99%) in the mRNA-1273p groups were attributed to delta. The incidence rate of Covid-19 was lower for the mRNA-1273p (49.0/1000 person-years) versus mRNA-1273e (77.1/1000 person-years) group [36.4% (95% CI 17.1%-51.5%) reduction]. There were fewer severe Covid-19 cases in the mRNA-1273p (6; 6.2/1000 person-years) than mRNA-1273e (13; 3.3/1000 person-years) [46.0% (95% CI −52.4%-83.2%) reduction]. Three Covid-19 related hospitalizations occurred with two resulting deaths in the mRNA-1273e group.

**Conclusion:** Incidence rates of Covid-19 and severe Covid-19 were lower during the months when delta was the dominant variant (July/August 2021) among COVE participants vaccinated more recently. Analysis of COVID-19 cases from the open-label phase of the COVE study is ongoing.

---

Following emergency use authorization (EUA) of the mRNA-1273 Covid-19 vaccine on 18-Dec-2020, the observer-blind, pivotal Coronavirus Efficacy (COVE) trial was amended to an open-label phase, where participants were unblinded and those randomized to placebo were offered vaccination.^1,2^ The surveillance of Covid-19 through the protocol-defined illness visits remained unchanged through the open label phase. The emergence of the delta variant of severe acute respiratory syndrome coronavirus 2 (SARS-CoV-2) in the US was associated with increased incidence of Covid-19 in the community among unvaccinated as well as vaccinated persons.^3–5^ Herein we describe Covid-19 incidences during a portion of the open-label phase 01July through 27-August-2021, in participants initially randomized to the mRNA-1273 group (vaccinated from July-December 2020) and those initially randomized to the placebo group (vaccinated December 2020-April 2021) in the modified-intent-to treat (mITT) population. Included in this analysis were 14,746 participants in the earlier mRNA-1273 (mRNA-1273e) group and 11,431 in the later placebo-mRNA-1273 (mRNA-1273p) group. The baseline characteristics of the participants were similar between the groups except for the fraction of participants ≥65 years of age, and healthcare workers (Table S1). Median follow-up times from the first dose were 13 months in the mRNA-1273e (including double-blind and open-label phases) and 7.9 months in the mRNA-1273p (only open-label phase) groups.

Covid-19 cases from the beginning of the open-label phase through June 2021 were low with an increase observed in July and August 2021 (Fig. S1). The greatest numbers of cases were observed in Texas, Florida and California (Table S2). While an increased number of breakthrough cases occurred during the open-label period, the incidence rates of Covid-19 starting after the second dose of mRNA-1273 in the trial were overall low for the mRNA-1273e (19.6) cases/1000 person-year, and much lower than the placebo group during the blinded study phase (134) (Table S3). During July and August, 162 Covid-19 cases occurred in the mRNA-1273e group and 88 in the mRNA-1273p group (Table 1 and Fig. S2). Of the cases sequenced, 144/149 (97%) in the mRNA-1273e and 86/87 (99%) in the mRNA-1273p groups were caused by the delta variant (Table S4). The incidence rate of Covid-19 was lower among the mRNA-1273p (49.0/1000 person-years) group than the mRNA-1273e (77.1/1000 person-years) group with a 36.4% (95% CI 17.1%-51.5%) reduction in the observed incidence rates. Incidence rates reductions were more pronounced in the younger than the older age groups. Similar results were seen using an adjusted Cox proportional model (Table S5).

**Table 1.** Number of Covid-19 Cases and Incidence Rates After mRNA-1273 Vaccination During July 1st to August 27, 2021 in the Modified Intent-to-Treat Population.

| Covid-19 Cases† | mRNA-1273e<br>N=14746 |  |  | mRNA-1273p*<br>N=11431 |  |  | mRNA-1273p vs<br>mRNA-1273e |
| --- | --- | --- | --- | --- | --- | --- | --- |
|  | Cases<br>n | Person-yr | Rate/1000<br>Person-yr | Cases<br>n | Person-yr | Rate/1000<br>Person-yr | Reduction of observed<br>incidence rate % (95% CI) |
| All cases | 162 | 2102 | 77.1 | 88 | 1796 | 49.0 | 36.4 (17.1-51.5) |
| ≥18-<65<br>yr | 136 | 1558 | 87.3 | 68 | 1289 | 52.8 | 39.6 (18.6-55.5) |
| ≥65 yr | 26 | 544 | 47.8 | 20 | 507 | 39.5 | 17.4 (-53.9-56.3) |
| Severe | 13 | 2102 | 6.2 | 6 | 1796 | 3.3 | 46.0 (-52.4-83.2) |
| ≥18-<65<br>yr | 7 | 1558 | 4.5 | 4 | 1289 | 3.1 | 30.9 (-171.7- 85.2) |
| ≥65 yr | 6 | 544 | 11.0 | 2 | 507 | 3.9 | 64.2 (-100.2-96.5) |
| mRNA-1273e =mRNA-1273 earlier and includes participants who were originally randomized and received mRNA-1273 in the blinded phase (vaccinated from 27 July 2020 to 16 Dec 2020). mRNA-1273p= placebo-mRNA-1273 treated and includes participants who were originally randomized to placebo in the blinded phase and receiving mRNA-1273 in the open-label phase (vaccinated from 29 Dec 2020 to 30 Apr 2021 post-EUA). Yr=years; CI=confidence interval. N denotes the number of participants with negative SARS-CoV-2 status at study entry (modified intent-to treat) who were at risk pre-vaccination. For this analysis, participants at risk at the start of the analysis period (01-Jul-21 to 27-Aug-21) were included and person-years was calculated for this analysis period. Incidence rate was defined as the number of Covid-19 cases divided by the number of participants at risk at the beginning of the time period (July 1, 2021) and adjusted by person-years in each group. To assess the difference in incidence rates of Covid-19 between the mRNA-1273 group and the placebo-mRNA-1273 group, the 1- ratio of incidence rate was calculated and provided with the 95% confidence intervals calculated using the exact method (Poisson distribution) adjusting for person-years.*Placebo-mRNA-1273 participants not considered at risk in the open-label phase were excluded from this analysis: those who (i) were COVID-19 or SARS-CoV-2 infection during the blinded phase, (ii) did not enter open-label or received off-study COVID-19 vaccine, or (iii) became a case prior to 1st dose of mRNA in the open-label phase. †Covid-19 cases starting 14 days after dose 2. |  |  |  |  |  |  |  |

There were 13 protocol-specified severe Covid-19 cases in the mRNA-1273e (6.2/1000 person-years) and 6 (3.3/1000 person-years) in the mRNA-1273p groups with an estimated 46.0% (95% CI −52.4%-83.2%) reduction of the observed incidence rate for later compared with earlier vaccinations during July-August (Table 1). There were three Covid-19 related hospitalizations all in the mRNA-1273e group, two of which resulted in death, both in men ≥70 years of age with medical comorbidities more than 10 months after vaccination (Table S6).

Overall, lower incidence rates of Covid-19 and fewer severe Covid-19 cases were observed during July-August 2021 when delta was the dominant variant, among participants vaccinated more recently. Limitations include the open-label design, lack of randomization and 2-month analysis duration. Analysis of Covid-19 cases from the open-label phase of the COVE study is ongoing.

## Methods

This is an exploratory analysis of the previously reported phase 3, randomized, observer-blind, placebo-controlled Coronavirus Efficacy (COVE) trial that enrolled medically stable adults at 99 US sites (clintrials.gov NCT04470427).^1,2^ Eligible participants included adults 18 years or older with no known history of SARS-CoV-2 infection, whose circumstances put them at appreciable risk for SARS-CoV-2 infection and/or high risk of severe Covid-19. Participants were randomized 1:1 to receive mRNA-1273 vaccine (100 µg) or placebo and stratified by age and Covid-19 complications risk criteria (≥18 and <65 years and not at risk, ≥18 and <65 years and at risk, and ≥65 years). Following emergency authorization (EUA) issuance of mRNA-1273, the protocol was amended to two-parts (A and B; protocol available at NEJM.org). Part A, was the observer-blind phase, that concluded when participants unblinded, with consideration given to those on placebo to receive mRNA-1273. Part B, is the open-label ongoing phase of the study. Participants continue to be followed for up to two years as originally planned.

The trial is conducted in accordance with the International Council for Technical Requirements for Registration of Pharmaceuticals for Human Use, Good Clinical Practice Guidance, and applicable government regulations. The central Institutional Review Board/Ethics Committee, Advarra, Inc., 6100 Merriweather Drive, Columbia, MD 21044 approved the protocol and consent forms. All participants provided written informed consent.

The primary endpoint was vaccine efficacy of mRNA-1273 at preventing a first occurrence of symptomatic Covid-19 with onset ≥14 days post-second injection, Covid-19 cases defined as having ≥2 of systemic symptoms (fever ≥ 38ºC, chills, myalgia, headache, sore throat, new olfactory and taste disorder[s]), or experienced ≥1 respiratory sign or symptom (cough, shortness of breath, or clinical or radiological evidence of pneumonia), and confirmed by positive RT-PCR for SARS-CoV-2 using NP swab, nasal or saliva sample. Severe Covid-19, a secondary endpoint, was defined as confirmed Covid-19 per the primary endpoint case definition, plus one of the clinical signs indicative of severe systemic illness (respiratory rate ≥30 per minute, heart rate ≥125 beats per minute, SpO2 ≤ 93% on room air at sea level or PaO2/FIO2 <300 mm Hg, OR respiratory failure or Acute Respiratory Distress Syndrome [ARDS, defined as needing high-flow oxygen, non-invasive or mechanical ventilation, or ECMO], evidence of shock [systolic blood pressure <90 mmHg, diastolic BP < 60 mmHg or requiring vasopressors], OR significant acute renal, hepatic or neurologic dysfunction, OR admission to an intensive care unit or death). Assessment of the severity of unsolicited adverse events was based on investigator-medical judgement and defined as mild (events that did not interfere with the participant’s daily activities), moderate (events that caused some interference with the participants daily activities requiring limited or no medical intervention) and severe (events preventing participant’s daily activity requiring intensive therapeutic intervention).

This exploratory analysis evaluated Covid-19 cases in the ongoing COVE trial, with a focus on those identified between 01-Jul-2021 and 27-Aug-in the mITT population that included all randomized participants who received at least one dose of study treatment and were SARS-CoV-2 negative at baseline of study entry. The analysis was performed in participants who were randomized and received mRNA-1273 (mRNA-1273 group; vaccinated from 27-July-2020 to 16-Dec-2020) in the blinded phase, and participants randomized to placebo in the blinded phase and receiving mRNA-1273 in the open-label phase (Placebo-mRNA-1273 group; vaccinated from 29-Dec-2020 to 30-Apr-2021) and were SARS-CoV-2 uninfected at the beginning of the open-label phase. For this analysis, the live, ongoing database was queried for Covid-19 cases that occurred after 26-Mar-2021 which met the following criteria (i) positive RT-PCR results of nasopharyngeal samples tested at the central lab and (ii) reporting of Covid-19 symptoms. The date of Covid-19 was the later date of positive RT-PCR or reporting of symptoms. These Covid-19 cases were incorporated with data in a locked database with a data cutoff date of 26-March-2021. Surveillance for Covid-19 remains unchanged for this ongoing study. All study participants receive weekly contact (via e-diary prompts; supplemented with monthly telephone calls) to inquire about the occurrence of Covid-19 symptoms that would lead to a clinic or home medical staff visit. Genetic sequencing of Covid-19 cases was also performed and the proportion of delta variants (B.1.617.2 and all AY lineages) in cases are also provided as of September 7, 2021.

The number of Covid-19 cases and calculated incidence rates from 01-Jul-2021 through 27-Aug-2021 are presented. Participants at risk at the start of the analysis period (01-Jul-21 to 27-Aug-21) were included and person-years were calculated. Participants not considered at risk in the open-label phase were excluded from this analysis including those who (i) were Covid-19 or SARS-CoV-2 infection cases during the blinded phase, (ii) did not enter the open-label phase or received off-study Covid-19 vaccine, or (iii) became a case prior to the first dose of mRNA in the open-label phase. Incidence rate was defined as the number of Covid-19 cases divided by the number of participants at risk at the beginning of the time period (July 1, 2021) and adjusted by person-years in each group. To assess the difference in incidence rates of Covid-19 between the mRNA-1273 group and the placebo-mRNA-1273 group, 1-ratio of incidence rates, was calculated with 95% confidence intervals.

Limitations of this analysis include a differential distribution between the mRNA-1273e and mRNA-1273p groups due to the amended study from a blinded to open-label phase and cross-over of placebo participants, and lack of randomization. While there also exists a potential for bias attributed to differences in the risk of those remaining on the study; the demographics suggest a general balance in group characteristics between the groups. Although the findings are based on an analysis of incidence rates, findings in the Cox proportional analysis, adjusted by the original risk stratification of the study showed similar results. Additionally, the analysis is limited to a short 2-month duration of July to August and with a longer follow-up times the result and differences between the groups may change. Please note the Covid-19 cases in this analysis are pending adjudication by the endpoint adjudication committee.

## Supporting information

Consort checklist

Protocol

## Data Availability

As the trial is ongoing, access to patient-level data and supporting clinical documents with qualified external researchers may be available upon request and subject to review once the trial is complete.

https://www.ncbi.nlm.nih.gov/pubmed/33378609

