## Supplement for "Covid-19 in the Phase 3 Trial of mRNA-1273 During the Delta-variant Surge"

#### Table S1. Baseline Demographics and Characteristics, Open-Label At-risk participants in Modified-intent-to-treat

| Characteristics n (%) | mRNA-1273e N=14746 | mRNA-1273p N=11431 |
|---|---|---|
| Sex | | |
| Male | 7678 (52.1) | 6012 (52.6) |
| Female | 7068 (47.9) | 5419 (47.4) |
| Age at Screening (yr) | 51.5 | 52.7 |
| Mean (range) | 18-95 | 18-95 |
| Age (yr) and health risk for severe Covid-19* | | |
| ≥18 and <65 and Not at Risk | 8575 (58.2) | 6310 (55.2) |
| ≥18 and <65 and at Risk | 2467 (16.7) | 1905 (16.7) |
| ≥65 | 3704 (25.1) | 3216 (28.1) |
| Ethnicity | | |
| Hispanic or Latino | 2957 (20.1) | 2168 (19.0) |
| Not Hispanic or Latino | 11650 (79.0) | 9161 (80.1) |
| Not reported or unknown | 139 (0.9) | 102 (0.9) |
| Race‡ | | |
| White | 11738 (79.6) | 9146 (80.0) |
| Black or African American | 1463 (9.9) | 1131 (9.9) |
| Asian | 645 (4.4) | 482 (4.2) |
| American Indian or Alaska Native | 113 (0.8) | 92 (0.8) |
| Native Hawaiian or Other Pacific Islander | 36 (0.2) | 22 (0.2) |
| Multiracial | 310 (2.1) | 259 (2.3) |
| Other | 288 (2.0) | 210 (1.8) |
| Not reported or unknown | 153 (1.0) | 89 (0.7) |
| Risk Factor for Severe Covid-19 at Screening§ | | |
| At Risk | 3375 (22.9) | 2744 (24.0) |
| Chronic lung disease | 702 (4.8) | 601 (5.3) |
| Significant cardiac disease | 745 (5.1) | 623 (5.5) |
| Severe obesity | 1041 (7.1) | 813 (7.1) |
| Diabetes | 1431 (9.7) | 1169 (10.2) |
| Liver disease | 102 (0.7) | 76 (0.7) |
| Human Immunodeficiency Virus Infection | 88 (0.6) | 70 (0.6) |
| Occupational Risk | 12115 (82.2) | 9142 (80.0) |
| Healthcare Workers | 3721 (25.2) | 2177 (19.0) |
| Body Mass Index, ($kg/m^2$) | | |
| n | 14659 | 11362 |
| Mean (SD) | 29.3 (6.8) | 29.4 (6.7) |

Percentages are based on the number of participants in the open label at-risk modified intent-to treat set. mRNA-1273 includes participants originally randomized to mRNA-1273 in the blinded phase (vaccinated from 27-July-2020 to 16-Dec-2020). Placebo-mRNA-1273 includes participants originally randomized to placebo in the blinded phase and who received mRNA-1273 post-EUA (vaccinated from 29-Dec-2020 to 30-Apr-2021). *Based on stratification factor from IRT, participants who were <65 years old were categorized as at risk for severe Covid-19 illness if they had at least 1 of the risk factors specified in the study protocol at screening. §Participants could be under one or more categories and were counted once at each category.

### Table S2. Geographical Breakdown of Covid-19 Cases in the COVE Trial July-August 2021

| State | mRNA-1273p | mRNA-1273e | Total Cases |
|---|---|---|---|
| **Total** | **88** | **162** | **250** |
| Texas | 17 | 40 | 57 |
| Florida | 16 | 26 | 42 |
| California | 4 | 14 | 18 |
| North Carolina | 8 | 6 | 14 |
| Kansas | 5 | 7 | 12 |
| Nevada | 3 | 9 | 12 |
| Tennessee | 1 | 10 | 11 |
| Louisiana | 6 | 4 | 10 |
| South Carolina | 4 | 6 | 10 |
| Arkansas | 1 | 8 | 9 |
| Missouri | 3 | 5 | 8 |
| Illinois | 4 | 2 | 6 |
| Maryland | 1 | 5 | 6 |
| Ohio | 2 | 3 | 5 |
| Georgia | 1 | 3 | 4 |
| Arizona | 2 | 1 | 3 |
| District of Columbia | 2 | 1 | 3 |
| Washington | 1 | 2 | 3 |
| Massachusetts | 1 | 1 | 2 |
| Mississippi | 1 | 1 | 2 |
| Nebraska | 2 | 0 | 2 |
| Oklahoma | 0 | 2 | 2 |
| Oregon | 1 | 1 | 2 |
| South Dakota | 2 | 0 | 2 |
| Colorado | 0 | 1 | 1 |
| Michigan | 0 | 1 | 1 |
| New York | 0 | 1 | 1 |
| Pennsylvania | 0 | 1 | 1 |
| Utah | 0 | 1 | 1 |
| Cases starting 14 days after second dose | | | |

**Table S3. Covid-19 Cases in Ongoing COVE Trial Blinded and Blinded + Open-label Phases**

| | Blinded Phase* | | Blinded + Open-Label Phase* |
|---|---|---|---|
| | Placebo | mRNA-1273 | mRNA-1273e |
| Median follow-up, mon | 5.3 mon (148 days)† | | 13 mon (364 days)‡ |
| Cases, n | 751 | 58 | 277 |
| Person-yr | 5604 | 5863 | 14110 |
| Rate/1000 person yr | 134 | 9.9 | 19.6 |

mon-=months; Yr=years. *Participants were randomized to placebo or mRNA-1273 and received one dose. mRNA-1273 group: participants randomized to mRNA-1273 (vaccinated from 27-July-2020 to 16-Dec-2020). †From first dose of mRNA-1273 or placebo to study discontinuation, unblinding/PDV, data-cutoff date 3/26/21, whichever occurred earlier. ‡From first dose of mRNA-1273 to study discontinuation, case date, data-cutoff date of 8/27/21, whichever occurred earlier. 1 month= 28 days. Cases starting 14 days after second dose.

**Table S4. Delta Variants in Covid-19 Cases Sequenced 14 Days After Dose 2 July and August 2021**

| n (%) | mRNA-1283e | mRNA-1273p |
|---|---|---|
| Cases sequenced total | 149 | 87 |
| Delta variant in cases sequenced | 144 (97.0) | 86 (99.0) |
| Non-delta variant in cases sequenced | 5 (3.4) | 1 (1.0) |

Covid-19 cases as of data cutoff date of 27-Aug-2021. The proportion of the observed COVID-19 cases attributed to the delta variant includes B.1.617.2, AY.1, AY.2, AY.3, AY.4, AY.5, AY.6, AY.7, AY.8, Ay.9, AY.10, AY.11, and AY lineages. CDC https://www.cdc.gov/coronavirus/2019-ncov/cases-updates/variant-surveillance/variant-info.html#Concern Sequence data as of 07-Sep-2021. Results are based on ongoing sequencing and may be subject to refinement.

### Table S5. Cox Proportional Hazards Model of Covid-19 Cases

| Covid-19 cases July – August 2021* (started ≥14 days after dose 2 of mRNA-1273) | mRNA-1273p N=11431 Cases n | mRNA-1273e N=14746 Cases n | mRNA-1273p vs. mRNA-1273e Reduction in hazard ratio % (95% CI) |
|---|---|---|---|
| Adjusting for stratification factor (<65yr and not at risk, <65yr and at risk, ≥65yr) | 88 | 162 | 36.1 (17.2-50.7) |
| Adjusting for Healthcare Worker (Yes, No) | | | 36.9 (18.1-51.6) |
| Adjusting for stratification factor and Healthcare Worker | | | 36.6 (17.3-51.1) |

A stratified Cox proportional hazards model was used to assess the difference between those who crossed-over to receive mRNA-1273 with those in terms of reduction in hazard ratio. Results from separate models adjusting for different covariate/baseline characteristics are consistent with those reported in Table 1.

**Table S6. Details of Severe COVID-19 Cases**

| Cases | mRNA-1273e | mRNA-1273p | Total (% of cases) |
|---|---|---|---|
| COVID-19 (per protocol) | 162 | 88 | 250 (100%) |
| Severe (per protocol) | 13 | 6 | 19 (7.6%) |
| Hospitalization | 3 | 0 | 3 (1.2%) |
| Death | 2 | 0 | 2 (0.8%) |
| Majority of severe cases met criteria for low SpO2 (range 88-93%). All severe cases had >5 symptoms (range of 5-13). | | | |

**Figure S1. Monthly Covid-19 Cases Accrued in the COVE Trial 14 days After Dose 2 of mRNA-1273**

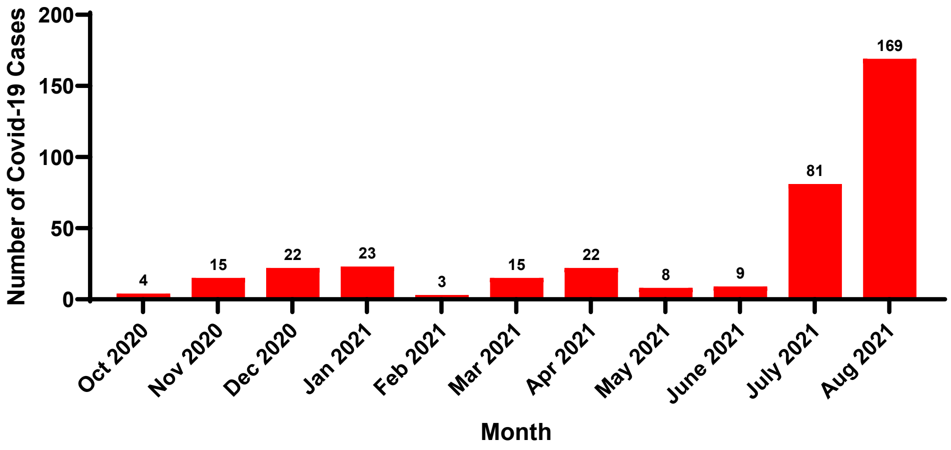


Genetically sequenced Covid Cases accrued in the COVE trial by month as of data cut-off date of 27-Aug-2021, from sequence data through Sept 7th, 2021. Cases were counted starting 14 days after dose 2.
