## Supplementary material for "Covid-19 in the Phase 3 Trial of mRNA-1273 During the Delta-variant Surge": Protocol

### CLINICAL STUDY PROTOCOL

| | |
|---|---|
| **Protocol Title:** | A Phase 3, Randomized, Stratified, Observer-Blind, Placebo-Controlled Study to Evaluate the Efficacy, Safety, and Immunogenicity of mRNA-1273 SARS-CoV-2 Vaccine in Adults Aged 18 Years and Older |
| **Protocol Number:** | mRNA-1273-P301 |
| **Sponsor Name:** | ModernaTX, Inc. |
| **Legal Registered Address:** | 200 Technology Square<br>Cambridge, MA 02139 |
| **Sponsor Contact and Medical Monitor:** | Allison August, MD<br>VP, Clinical Development<br>ModernaTX, Inc.<br>200 Technology Square, Cambridge, MA 02139<br>Telephone: 1-617-678-8937<br> |
| **Regulatory Agency Identifier Number(s):** | IND: 19745 |
| **Amendment Number:** | 8 |
| **Date of Amendment 8:** | 23 Mar 2021 |
| **Date of Amendment 7:** | 10 Feb 2021 |
| **Date of Amendment 6:** | 23 Dec 2020 |
| **Date of Amendment 5:** | 11 Nov 2020 |
| **Date of Amendment 4:** | 30 Sep 2020 |
| **Date of Amendment 3:** | 20 Aug 2020 |
| **Date of Amendment 2:** | 31 Jul 2020 |
| **Date of Amendment 1:** | 26 Jun 2020 |
| **Date of Original Protocol:** | 15 Jun 2020 |

#### CONFIDENTIAL

All financial and nonfinancial support for this study will be provided by ModernaTX, Inc. The concepts and information contained in this document or generated during the study are considered proprietary and may not be disclosed in whole or in part without the expressed written consent of ModernaTX, Inc. The study will be conducted according to the *International Council for Harmonisation (ICH) Technical Requirements for Registration of Pharmaceuticals for Human Use, E6(R2) Good Clinical Practice (GCP) Guidance.*

##### Protocol Approval – Sponsor Signatory

**Study Title:** A Phase 3, Randomized, Stratified, Observer-Blind, Placebo-Controlled Study to Evaluate the Efficacy, Safety, and Immunogenicity of mRNA-1273 SARS-CoV-2 Vaccine in Adults Aged 18 Years and Older

**Protocol Number:** mRNA-1273-P301

**Protocol Version Date:** 23 Mar 2021

Protocol accepted and approved by:

**See signature and date signed on last page of document**

Allison August, MD
VP, Clinical Development
ModernaTX, Inc.
200 Technology Square
Cambridge, MA 02139

Date

### DECLARATION OF INVESTIGATOR

I have read and understood all sections of the protocol entitled "A Phase 3, Randomized, Stratified, Observer-Blind, Placebo-Controlled Study to Evaluate the Efficacy, Safety, and Immunogenicity of mRNA-1273 SARS-CoV-2 Vaccine in Adults Aged 18 Years and Older" and the most recent version of the Investigator's Brochure.

I agree to supervise all aspects of the protocol and to conduct the clinical investigation in accordance with the current Protocol, the *International Council for Harmonisation (ICH) Technical Requirements for Registration of Pharmaceuticals for Human Use, E6(R2) Good Clinical Practice (GCP) Guidance,* and all applicable government regulations. I will not make changes to the protocol before consulting with ModernaTX, Inc. or implement protocol changes without IRB/IEC approval except to eliminate an immediate risk to participants.

I agree to administer study treatment only to participants under my personal supervision or the supervision of a sub-investigator. I will not supply study treatment to any person not authorized to receive it. I also agree that persons debarred from conducting or working on clinical studies by any court or regulatory agency will not be allowed to conduct or work on studies for the Sponsor or a partnership in which the Sponsor is involved. I will immediately disclose it in writing to the Sponsor if any person who is involved in the study is debarred, or if any proceeding for debarment is pending, or, to the best of my knowledge, threatened.

I will not disclose confidential information contained in this document including participant information, to anyone other than the recipient study staffs and members of the IRB/IEC. I agree to ensure that this information will not be used for any purpose other than the evaluation or conduct of the clinical investigation without the prior written consent from ModernaTX, Inc. I will not disclose information regarding this clinical investigation or publish results of the investigation without authorization from ModernaTX, Inc.

The signature below provides the necessary assurance that this study will be conducted according to all stipulations of the protocol, including statements regarding confidentiality, and according to local legal and regulatory requirements, US federal regulations, and ICH E6(R2) GCP guidelines.

____________________________________ ____________________

Signature of Principal Investigator Date

____________________________________

Printed Name of Principal Investigator

### Protocol Amendment Summary of Changes

| DOCUMENT HISTORY | |
|---|---|
| **Document** | **Date** |
| Amendment 8 | 23 Mar 2021 |
| Amendment 7 | 10 Feb 2021 |
| Amendment 6 | 23 Dec 2020 |
| Amendment 5 | 11 Nov 2020 |
| Amendment 4 | 30 Sep 2020 |
| Amendment 3 | 20 Aug 2020 |
| Amendment 2 | 31 Jul 2020 |
| Amendment 1 | 26 Jun 2020 |
| Original Protocol | 15 Jun 2020 |

**Amendment 8, 23 Mar 2021**

**Main Rationale for the Amendment:**

The purpose of this amendment is to update language around unblinding and open-label dosing in the context of the majority of participants already having completed their blinded follow-up. In addition, the amendment updates the reporting of serious COVID-19 cases.

The summary of changes table provided here describes the major changes made in Amendment 8 relative to Amendment 7, including the sections modified and the corresponding rationales. As applicable, the synopsis of Amendment 8 has been modified to correspond to changes in the body of the protocol.

**Summary of Major Changes from Protocol Amendment 7 to Protocol Amendment 8:**

| Section # and Name | Description of Change | Brief Rationale |
|---|---|---|
| Section 4.1.2 (Part B, the Open-label Observational Phase) | States that, due to ethical and scientific concerns, all participants will be unblinded once investigational product is no longer available. In addition, the text indicates that if participants want to receive emergency use authorization (EUA) vaccine outside of the study, participants will need to be withdrawn from the study.<br><br>The amendment also provides directions for unblinding after Biologics License Application (BLA) database lock. | At the time of this amendment, more than 95% of the enrolled participants have completed blinded follow-up, either through being unblinded or study discontinuation. Therefore, there is no scientific value for participants to remain blinded.<br><br>In addition, non-study EUA vaccines are widely available, so ethically, participants should be unblinded to ensure they can make an informed choice if they wish to receive EUA COVID-19 vaccine outside of the study. |

| Section # and Name | Description of Change | Brief Rationale |
|---|---|---|
| Section 8.3.7 (Time Period and Frequency for collecting Adverse Events and Serious Adverse Event Information) and Section 8.3.10 (Reporting Adverse Events) | Reporting of only serious COVID-19 cases within 24 hours to sponsor or it's designee. | Non-serious COVID-19 cases are an efficacy endpoint, so reporting to Pharmacovigilance is no longer warranted. |

### IRB and Regulatory Authority Approval

A copy of this amended protocol will be sent to the institutional review board (IRB) and regulatory authority.

The changes described in this amended protocol require IRB approval prior to implementation. In addition, if the changes herein affect the informed consent, sites are required to update and submit a revised informed consent for approval that incorporates the changes described in this amended protocol.

### 1. PROTOCOL SUMMARY

#### 1.1. Synopsis

| | |
|---|---|
| **Protocol Number:** | **mRNA-1273-P301** |
| **Title:** | A Phase 3, Randomized, Stratified, Observer-Blind, Placebo-Controlled Study to Evaluate the Efficacy, Safety, and Immunogenicity of mRNA-1273 SARS-CoV-2 Vaccine in Adults Aged 18 Years and Older. |
| **Study Phase:** | 3 |
| **Objectives:** | Primary: |

Primary:

- To demonstrate the efficacy of mRNA-1273 to prevent COVID-19.
- To evaluate the safety and reactogenicity of 2 injections of mRNA-1273 given 28 days apart.

Secondary:

- To evaluate the efficacy of mRNA-1273 to prevent severe COVID-19.
- To evaluate the efficacy of mRNA-1273 to prevent serologically confirmed SARS-CoV-2 infection or COVID-19 regardless of symptomatology or severity.
- To evaluate vaccine efficacy (VE) against a secondary definition of COVID-19.
- To evaluate VE to prevent death caused by COVID-19.
- To evaluate the efficacy of mRNA-1273 to prevent COVID-19 after the first dose of investigational product (IP).
- To evaluate the efficacy of mRNA-1273 to prevent COVID-19 in all study participants, regardless of evidence of prior SARS-CoV-2 infection.
- To evaluate the efficacy of mRNA-1273 to prevent asymptomatic SARS-CoV-2 infection.

Exploratory:

- To evaluate the effect of mRNA-1273 on the viral infection kinetics as measured by viral load at SARS-CoV-2 infection diagnosis by RT-PCR and number of days from the estimated date of SARS-CoV-2 infection until undetectable SARS-CoV-2 infection by RT-PCR.
- To assess VE to reduce the duration of symptoms of COVID-19.
- To evaluate VE against all-cause mortality.
- To assess VE against burden of disease (BOD) due to COVID-19.
- To evaluate the genetic and/or phenotypic relationships of isolated SARS-CoV-2 strains to the vaccine sequence.
- To evaluate immune response markers after dosing with IP as correlates of risk of COVID-19 and as correlates of risk of SARS-CoV-2 infection.
- To conduct additional analyses related to furthering the understanding of SARS-CoV-2 infection and COVID-19, including analyses related to the immunology of this or other vaccines, detection of viral infection, and clinical conduct.

**Study Design and Methodology:**

This is a two-part Phase 3 study: Part A and Part B (Figure 1). Participants in Part A, the Blinded Phase of this study are blinded to their treatment assignment. Given that the primary efficacy endpoint for mRNA-1273 against COVID-19 was met per the protocol-defined interim analysis (IA), Part B, the Open-Label Observational Phase of this study is designed to offer participants who received placebo in Part A of this study, an option to request Open-label mRNA-1273 while investigational vaccine is still available. With regards to eligibility in Part B, the CDC-EUA guidance specifications will supersede eligibility criteria in the protocol.

**Part A, the Blinded Phase**:

The Blinded Phase of this study (Figure 1) is a randomized, stratified, observer-blind, placebo-controlled evaluation of the efficacy, safety, and immunogenicity of mRNA-1273 SARS-CoV-2 vaccine compared to placebo in adults 18 years of age and older who have no known history of SARS-CoV-2 infection but whose locations or circumstances put them at appreciable risk of acquiring COVID-19 and/or SARS-CoV-2 infection.

Participants will be randomly assigned to receive injections of either 100 µg of mRNA-1273 vaccine or a placebo control in a 1:1 randomization ratio. Assignment will be stratified by age and health risk. This is a case-driven study and thus final sample size of the study will depend on the actual attack rate of COVID-19.

All participants will be assessed for efficacy and safety endpoints and provide a nasopharyngeal (NP) swab sample and blood sample before the first and second dose of IP, in addition to a series of post-dose blood samples for immunogenicity through 24 months after the second dose of IP. Efficacy assessments will include surveillance for COVID-19 with RT-PCR confirmation of SARS-CoV-2 infection after the first and second dose of IP.

As noted above, this is a case-driven study: if the prespecified criteria for early efficacy are met at the time of either IA or overall efficacy at the primary analysis, a final study report describing the efficacy and safety of mRNA-1273 will be prepared based on the data available at that time. In the event that success criteria are met either at the time of the IA or when the total number of cases toward the primary endpoint have accrued, participants will continue to be followed in a blinded fashion until Month 25 to enable assessment of long-term safety and durability of VE.

If the study concludes early, all participants will be requested to provide a final blood sample at the time of study conclusion.

Each participant will receive 2 doses of IP by 0.5 mL intramuscular (IM) injection, the first on Day 1 and the second on Day 29. An NP swab sample will be collected prior to the first and second dose of IP, for evaluation by RT-PCR. Participants will be given an electronic diary (eDiary) to report solicited adverse reactions (ARs) for 7 days after each dose of IP (Part A, Blinded Phase only) and to prompt an unscheduled clinic visit for clinical evaluation and NP swab sample if a participant experiences any symptoms of COVID-19. All participants will receive safety calls on Day 8, Day 15, Day 22, Day 36, and Day 43 that will serve both to monitor for unsolicited adverse events (AEs) and to monitor for symptoms of COVID-19.

Surveillance for COVID-19 will be performed through weekly contacts with the participant via a combination of telephone calls and completion of an eDiary starting at Day 1 through the end of the study.

Participants with symptoms of COVID-19 lasting at least 48 hours (except for fever and/or respiratory symptoms) will return to the clinic or will be visited at home by medically qualified site staff within 72 hours (an "Illness Visit") to collect an NP swab sample for RT-PCR testing for SARS-CoV-2 and other respiratory pathogens, or alternatively, if a clinic or home visit is not possible, will submit a saliva (or nasal swab) sample for SARS-CoV-2 RT-PCR testing. Any confirmed COVID-19 occurring in a participant will be captured as a medically attended adverse event (MAAE) along with relevant concomitant medications and details about severity, seriousness, and outcome. All confirmed serious COVID-19 cases will be reported to the Sponsor or designee within 24 hours.

Starting with the Illness Visit, study participants will be monitored by the study investigator (or appropriately delegated study staff) for a 14-day period after diagnosis or until symptoms resolve, whichever is later. Each participant diagnosed with COVID-19 will monitor their body temperature, oxygen saturation, and symptoms following the diagnosis of COVID-19. In addition to daily follow-up of symptoms, assessments will include the collection of saliva samples during the 28-day period following the diagnosis of SARS-CoV-2 infection. The investigator will determine if medical attention is required due to worsening of COVID-19. Finally, a convalescent visit will be scheduled approximately 28 days after the initial Illness Visit. At this visit, a saliva (or nasal swab) sample will be collected, and a blood sample will be drawn for immunologic assessment of SARS-CoV-2 infection.

The 28-day period following the Illness Visit is referred to as the Convalescent Period. If during the Convalescent Period the participant has a positive result for SARS-CoV-2 from the Illness Visit, the participant will continue the Convalescent Period. If the participant has a negative result for SARS-CoV-2 from the Illness Visit, the participant will exit the Convalescent Period, including discontinuation of daily telemedicine visits and collection of saliva (or nasal swab) samples, and will return to their respective study schedule.

At each dosing visit, participants will be instructed (Day 1) or reminded (Day 29) on how to document and report solicited ARs in the eDiary provided. Solicited ARs will be assessed for 7 days after each IP dose (Part A, the Blinded Phase only) and unsolicited AEs will be assessed for 28 days after each IP dose (Part A, the Blinded Phase only); serious adverse events (SAEs), MAAEs, AEs leading to withdrawal, will be assessed throughout the study.

**Part B, the Open-Label Observational Phase**:

Part B, the Open-Label Observational Phase of the study is prompted by the authorization of a COVID-19 vaccine under EUA. Transitioning the study to Part B permits all ongoing study participants to be informed of the availability and eligibility criteria of any COVID-19 vaccine made available under an EUA and the option to offer all ongoing study participants who request unblinding, an opportunity to schedule a Participant Decision clinic visit to know their original treatment assignment (placebo vs. mRNA-1273 vaccine).

Part B, the Open-Label Observational Phase of the study, also provides the opportunity for eligible study participants who previously received placebo to actively request to receive 2 doses of mRNA-1273 vaccine (as long as investigational vaccine is available). All study participants will receive a Notification Letter and will be asked to schedule a Participant Decision clinic visit. Principal Investigators should consider current local public health guidance for administration of COVID-19 vaccines under EUA when determining the scheduling priority of participants.

All participants will proceed to Part B, the Open-Label Observational Phase of the study, starting with a Participant Decision clinic visit (Figure 2).

At the Participant Decision clinic visit, all participants will:

- Be encouraged to remain in the ongoing study,
- Be given the option to be unblinded as to their original group assignment (placebo vs. mRNA-1273 vaccine),
- Be counselled about the importance of continuing other public health measures to limit the spread of disease including social distancing, wearing a mask, and hand-washing,
- Sign a revised informed consent form,
- Provide a NP swab for RT-PCR for SARS-CoV-2 and a blood sample for serology prior to unblinding.

The following participants will continue with the original study Schedule of Events (SoEs) presented in Table 16, Table 17, Table 18, and Table 19 (Figure 2):

- participants who request to not be unblinded,
- participants who request to be unblinded and received mRNA-1273, and
- participants who request to be unblinded and received placebo and choose to remain on placebo.

Participants who are unblinded, received placebo, are eligible, and request to receive mRNA-1273 (Figure 2) will have the following clinic visits as shown in the Supplemental SoE (Table 21):

- Open-label Dose 1 (OL-D1): To occur at the Participant Decision clinic visit or at a scheduled subsequent visit.
- Open-label Dose 2 (OL-D29): To occur 28 days after Dose 1 on OL-D1.
- Open-label Clinic Visit (OL-D57): To occur 1-month after Dose 2 on OL-D29.
- in addition to continuing to comply with the Original Study Schedule of Events (Table 16, Table 17, Table 18 and Table 19, as applicable).

Participants who are unblinded, received ONLY 1 dose of mRNA-1273, are eligible, and request to receive mRNA-1273 (Figure 2) will have the following clinic visits as shown in the Modified Supplemental Schedule of Events (Table 22):

- Their second dose on OL-D1: To occur at the Participant Decision clinic visit or at a scheduled subsequent visit
- And return on OL-D29: To occur 1-month after Dose 2 on OL-D1.
- in addition to continuing to comply with the Original Study Schedule of Events (Table 16, Table 17, Table 18, and Table 19, as applicable).

All participants remain on their original SoE to complete 24 months of follow-up after Dose 2; study participants who enter the Supplemental/Modified Supplemental SoE as described above do so in addition to their original SoE. Accordingly, all study participants will complete the full study follow-up to 24 months after the second dose of their original inoculation (mRNA-1273 or placebo).

| | |
|---|---|
| | At a point when the mRNA-1273 vaccine is no longer available for study use, any participant who schedules a Participant Decision Visit after this time-point will be unblinded (with no option to stay blinded), but will not be offered to receive mRNA-1273 study vaccine. These participants will have all of the same procedures performed outlined in the SoE table, with the exception of being offered to receive mRNA-1273 vaccine..<br><br>After the Biologics License Application (BLA) database lock, any remaining participants who have not been unblinded will be unblinded by the Moderna study team and will receive their unblinding treatment assignment (placebo vs. mRNA vaccine) via certified letter from the site..<br><br>This study will be conducted in compliance with the protocol, Good Clinical Practice (GCP), and all applicable regulatory requirements. |
| **Randomization:** | In Part A, the Blinded Phase of the study, approximately 30,000 participants will be randomly assigned in 1:1 ratio to receive either mRNA-1273 100 µg or placebo. The randomization will be in a blinded manner using a centralized Interactive Response Technology, in accordance with pre-generated randomization schedules. |
| **Stratification:** | Randomization in Part A, the Blinded Phase of the study, will be stratified based on age and, if they are < 65 years of age, based on the presence or absence of risk factors for severe illness from COVID-19 based on CDC recommendation as of March 2020. There will be 3 strata for randomization: ≥ 65 years, < 65 years and categorized to be at increased risk ("at risk") for the complications of COVID-19, and < 65 years "not at risk". Risk will be defined based on the study participants' relevant past and current medical history. At least 25% of enrolled participants, up to 50%, will be either ≥ 65 years of age or < 65 years of age and "at risk" at Screening.<br><br>Participants who are < 65 years old will be categorized as at risk for severe COVID-19 illness if they have at least 1 of the following risk factors at Screening:<br><br>• Chronic lung disease (eg, emphysema and chronic bronchitis, idiopathic pulmonary fibrosis, and cystic fibrosis) or moderate to severe asthma |

- Significant cardiac disease (eg, heart failure, coronary artery disease, congenital heart disease, cardiomyopathies, and pulmonary hypertension)
- Severe obesity (body mass index ≥ 40 kg/m$^2$)
- Diabetes (Type 1, Type 2 or gestational)
- Liver disease
- Human Immunodeficiency Virus (HIV) infection

| | |
|---|---|
| **Study Population:** | Participants (males and females 18 years of age or older at time of consent), who are at risk of SARS-CoV-2 infection with no known history of SARS-CoV-2 infection, are a subset of the planned target population. Additionally, potential study participants at increased risk of complications from COVID-19 will be included, since it is hypothesized that these participants might derive the greatest benefit from a vaccine. Participants ≥ 65 years of age will be eligible for enrollment with or without underlying medical conditions further increasing their risk of severe COVID-19.<br>Study sites may be selected based on SARS-COV-2 infection risk of the local population. Approximately 30,000 participants will be enrolled.<br>The full lists of inclusion and exclusion criteria are provided in the body of the protocol. |
| **Efficacy Assessments:** | **Primary Efficacy Assessment:**<br>To be considered as a case of COVID-19 for the evaluation of the Primary Efficacy Endpoint, the following criteria must be met:<br>• The participant must have experienced at least TWO of the following systemic symptoms: Fever (≥ 38ºC), chills, myalgia, headache, sore throat, new olfactory and taste disorder(s), OR<br>• The participant must have experienced at least ONE of the following respiratory signs/symptoms: cough, shortness of breath or difficulty breathing, OR clinical or radiographical evidence of pneumonia; AND<br>• The participant must have at least one NP swab, nasal swab, or saliva sample (or respiratory sample, if hospitalized) positive for SARS-CoV-2 by RT-PCR. |

**Secondary Efficacy Assessments:**

To be considered a severe COVID-19, the following criteria must be met: a confirmed COVID-19 as per the Primary Efficacy Endpoint case definition, plus any of the following:

- Clinical signs indicative of severe systemic illness, Respiratory Rate ≥ 30 per minute, Heart Rate ≥ 125 beats per minute, $SpO_2 \leq 93\%$ on room air at sea level or $PaO_2/FIO_2 < 300$ mm Hg, OR
- Respiratory failure or Acute Respiratory Distress Syndrome (ARDS), (defined as needing high-flow oxygen, non-invasive or mechanical ventilation, or ECMO), evidence of shock (systolic blood pressure < 90 mmHg, diastolic BP < 60 mmHg or requiring vasopressors), OR
- Significant acute renal, hepatic or neurologic dysfunction, OR
- Admission to an intensive care unit or death.

The secondary case definition of COVID-19 is defined as the following systemic symptoms: fever (temperature ≥ 38ºC) or chills, cough, shortness of breath or difficulty breathing, fatigue, muscle aches or body aches, headache, new loss of taste or smell, sore throat, nasal congestion or rhinorrhea, nausea or vomiting or diarrhea AND a positive NP swab, nasal swab, or saliva sample (or respiratory sample, if hospitalized) for SARS-CoV-2 by RT-PCR.

Death attributed to COVID-19 is defined as any participant who dies during the study with a cause directly attributed to a complication of COVID-19.

Asymptomatic SARS-CoV-2 infection is determined by seroconversion due to infection assessed by binding antibody (bAb) levels against SARS-CoV-2 as measured by a ligand-binding assay specific to the SARS-CoV-2 nucleocapsid protein and a negative NP swab sample for SARS-CoV-2 at Day 1.

**Immunogenicity Assessments:**

Immunogenicity assessments will include the following:

- Serum bAb levels against SARS-CoV-2 as measured by ligand-binding assay specific to the SARS-CoV-2 S protein.
- Serum bAb levels against SARS-CoV-2 as measured by ligand-binding assay specific to the SARS-CoV-2 nucleocapsid protein.
- Serum nAb titer against SARS-CoV-2 as measured by pseudovirus and/or live virus neutralization assays.

| | |
|---|---|
| **Safety Assessments:** | Safety assessments will include monitoring and recording of the following for each participant:<br>• Solicited local and systemic ARs that occur during the 7 days following each injection (ie, the day of injection and 6 subsequent days). Solicited ARs will be recorded daily using eDiaries. (Part A, Blinded Phase Only)<br>• Unsolicited AEs observed or reported during the 28 days following each injection (ie, the day of injection and 27 subsequent days). (Part A, Blinded Phase Only)<br>• AEs leading to discontinuation from dosing and/or study participation from Day 1 through Day 759 or withdrawal from the study.<br>• MAAEs from Day 1 through Day 759 or withdrawal from the study.<br>• SAEs from Day 1 through Day 759 or withdrawal from the study.<br>• Abnormal vital sign measurements.<br>• Physical examination findings.<br>• Pregnancy and accompanying outcomes.<br>• Concomitant medications and non-study vaccinations. |
| **Investigational Product, Dosage, and Route of Administration:** | The mRNA-1273 IP is an LNP dispersion of an mRNA encoding the prefusion stabilized S protein of SARS-CoV-2 formulated in LNPs composed of 4 lipids (1 proprietary and 3 commercially available): the proprietary ionizable lipid SM-102; cholesterol; 1,2-distearoyl-sn-glycero-3 phosphocholine (DSPC); and 1 monomethoxypolyethyleneglycol-2,3-dimyristylglycerol with polyethylene glycol of average molecular weight 2000 (PEG2000-DMG). The mRNA-1273 vaccine is provided as a sterile liquid for injection and is a white to off- white dispersion in appearance, at a concentration of 0.2 mg/mL in 20 mM Tris buffer containing 87 mg/mL sucrose and 10.7 mM sodium acetate at pH 7.5.<br><br>The placebo is 0.9% sodium chloride (normal saline) injection, which meets the criteria of the United States Pharmacopeia (USP).<br><br>Investigational product will be administered as an IM injection into the deltoid muscle on a 2-dose injection schedule on Day 1 and Day 29.<br><br>Each injection will have a volume of 0.5 mL and contain mRNA-1273 100 µg, or saline placebo. Preferably, vaccine should be |

| | |
|---|---|
| | administered into the nondominant arm. The second dose of IP should be administered in the same arm as the first dose.<br>Unblinded personnel, who will not participate in any other aspect of the study during Part A, the Blinded Phase, will perform IP accountability, dose preparation, and IP administration. Study site personnel who were blinded during the Blinded Phase, will be unblinded at the participant level at the Participant Decision clinic visit. |
| **Sample Size:** | The sample size is driven by the total number of cases to demonstrate VE (mRNA-1273 vs. placebo) to prevent COVID-19. Under the assumption of proportional hazards over time and with 1:1 randomization of mRNA-1273 and placebo, a total of 151 COVID-19 cases will provide 90% power to detect a 60% reduction in hazard rate (60% VE), rejecting the null hypothesis H0: VE ≤ 30%, with 2 IAs at 35% and 70% of the target total number of cases using a 1-sided O'Brien-Fleming boundary for efficacy and a log-rank test statistic with a 1-sided false positive error rate of 0.025. The total number of cases pertains to the Per-protocol (PP) Set accruing at least 14 days after the second dose. There are 2 planned IAs in this study, which will be performed when approximately 35% and 70% of the target total number of cases have been observed. Approximately 30,000 participants will be randomized with the following assumptions:<br>• The target VE against COVID-19 is 60% (with 95% confidence interval (CI) lower bound ruling out 30%, rejecting the null hypothesis H0: VE ≤ 30%).<br>• A 6-month COVID-19 incidence rate of 0.75% in the placebo arm.<br>• An annual dropout rate of 2% (loss of evaluable participants).<br>• Two IAs at 35% and 70% of total target cases across the 2 treatment groups with O'Brien-Fleming boundaries for efficacy monitoring.<br>• 3-month uniform accrual.<br>• Approximately 15% of participants will be excluded from the PP population, and participants are at risk for COVID-19 starting 14 days after the second dose.<br>**Power for Selected Secondary Efficacy Endpoints:**<br>For the secondary objective on VE against virologically confirmed SARS-CoV-2 infection or COVID-19 regardless of symptomology or |

| | |
|---|---|
| | severity (COV-INF), the study will have ≥ 90% power to demonstrate the VE is above 30% (to reject null hypothesis VE ≤ 30%) at 1-sided alpha of 2.5% if the true VE to prevent COV-INF is 60%, because every COVID-19 endpoint is necessarily a COV-INF endpoint.<br><br>For the secondary objective on VE against severe COVID-19, the power of demonstrating VE based on a total of 20 and 30 events under different scenarios of true VE and VE criteria has been calculated. |
| **Statistical Methods:** | **Statistical Hypotheses:** For the primary efficacy objective, the null hypothesis of this study is that the VE of mRNA-1273 to prevent first occurrence of COVID-19 is ≤ 30% (ie, $H_0^{efficacy}$: VE ≤ 0.3).<br><br>The study will be considered to meet the primary efficacy objective if the corresponding CI of VE rules out 30% at either one of the IA or at the primary analysis. In the primary analysis of VE of COVID-19, cases will be counted starting 14 days after the second dose of IP.<br><br>Vaccine efficacy is defined as the percent reduction in the hazard of the primary endpoint (mRNA-1273 vs placebo). Equivalently, the null hypothesis is:<br><br>• $H_0^{efficacy}$: hazard ratio (HR) ≥ 0.7 (equivalently, proportional hazards VE ≤ 0.3).<br><br>A stratified Cox proportional hazard model will be used to assess the magnitude of the treatment group difference (ie, HR) between mRNA-1273 and placebo at a 1-sided 0.025 significance level.<br><br>**Analysis Populations:** Analysis populations for statistical analyses are Randomization Set, Full Analysis Set (FAS), Modified Intent-to-Treat (mITT) Set, PP Set, Immunogenicity Subset, Solicited Safety Set, and Safety Set, as shown below:<br><br>• Randomization Set: All participants who are randomized, regardless of the participants' treatment status in the study.<br>• FAS: All randomized participants who received at least one dose of IP. Participants will be analyzed according to the group to which they were randomized. |

- mITT Set: All participants in the FAS who had no immunologic or virologic evidence of prior COVID-19 (ie, negative NP swab test at Day 1 and/or bAb against SARS-CoV-2 nucleocapsid below limit of detection or lower limit of quantification [LLOQ]) at Day 1 before the first dose of IP. Participants will be analyzed according to the group to which they were randomized.
- Immunogenicity Subset: All participants in the FAS who were sampled into subset for characterizing mRNA-1273 immunogenicity and had a valid immunogenicity test result prior to the first dose of IP and at least 1 valid result after the first dose of IP. Participants in the subset who had major protocol deviations that impact critical or key immunogenicity or study data may be excluded from the Immunogenicity Subset. The details of the Immunogenicity Subset will be documented prior to analysis of immunogenicity data.
- Solicited Safety Set: The Solicited Safety Set consists of all randomized participants who received at least one dose of IP and contributed any solicited AR data. The Solicited Safety Set will be used for the analyses of solicited ARs and participants will be included in the treatment group corresponding to the IP that they actually received.
- Safety SetAll randomized participants who received at least one dose of IP. The Safety Set will be used for all analyses of safety except for the solicited ARs. Participants will be included in the treatment group corresponding to the IP that they actually received.

**Efficacy Analyses:** Efficacy analyses will be performed using the FAS, mITT and PP populations, and participants will be included in the treatment group to which they are randomized. The primary analysis population for efficacy will be the PP Set.

The table below summarizes the analysis approach for primary and secondary efficacy endpoints. Sensitivity analysis methods are described for each endpoint as applicable.

| Endpoint | Statistical Analysis Methods |
|---|---|
| **Primary endpoint:**<br>VE of mRNA-1273 to prevent COVID-19 | • Primary analysis: VE will be estimated with 1 - HR (mRNA-1273 vs placebo) using a Cox proportional hazard regression model with treatment group as a fixed effect and adjust for |

| | |
|---|---|
| | stratification factor based on the PP Set, with cases counted starting 14 days after the second dose of IP.<br>• Analysis using the same model based on the mITT Set.<br>• Sensitivity analysis using the same model based on the PP Set, with cases counted starting either immediately after the second dose of IP or immediately after the first dose of IP.<br>• Subgroup analysis of the primary efficacy endpoint will be performed to assess consistency of VE, such as in the age groups ≥ 18 and < 65 years and ≥ 65 years.<br>• Supportive analysis of VE to be estimated with 1 - ratio of incidence rates with 95% CI using the exact method conditional upon the total number of cases.<br>• Supportive analysis of cumulative incidence VE. |

| | |
|---|---|
| **Secondary endpoints:**<br>• Vaccine efficacy of mRNA-1273 to prevent severe COVID-19<br>• Vaccine efficacy of mRNA-1273 to prevent serologically confirmed SARS-CoV-2 infection or COVID-19 regardless of symptomatology or severity.<br>• Vaccine efficacy of mRNA-1273 to prevent COVID-19 using a secondary definition of symptoms<br>• Vaccine efficacy of mRNA-1273 to prevent death caused by COVID-19<br>• Vaccine efficacy of mRNA-1273 to prevent COVID-19 after the first dose of IP<br>• Vaccine efficacy of mRNA-1273 to prevent asymptomatic SARS-CoV-2 infection | Similar analysis method as for the primary endpoint analysis.<br>For each of the secondary endpoints:<br>• Primary analysis: VE will be estimated with 1 - HR (mRNA-1273 vs placebo) using a Cox proportional hazard regression model with treatment group as a fixed effect and adjusting for stratification factor based on the PP Set, with cases counted starting 14 days after the second dose of IP.<br>• Analysis using the same model based on the mITT Set.<br>• Sensitivity analyses with cases counted starting immediately after the second dose of IP, 14 days after the first dose of IP, immediately after the first dose of IP, and immediately after randomization.<br>• Vaccine efficacy and 95% CI based on the case incidence will be estimated with 1 - ratio of incidence rates using the exact method conditional upon the total number of cases. |

| | |
|---|---|
| Vaccine efficacy of mRNA-1273 to prevent COVID-19 in all study participants, regardless of evidence of prior SARS-CoV-2 infection | The FAS population will be used for this secondary objective, using similar analysis methods as for the primary endpoint analysis.<br>• Primary analysis: VE will be estimated with 1 - HR (mRNA-1273 vs placebo) using a Cox proportional hazard regression model with treatment group as a fixed effect and adjusting for stratification factor based on the FAS, with cases counted starting 14 days after the second dose of IP.<br>• Sensitivity analyses with cases counted starting immediately after the second dose of IP, 14 days after the first dose of IP, immediately after the first dose of IP, and immediately after randomization. |

| | |
|---|---|
| **Long-term endpoints:**<br>Cases after the second injection of IP for participants in the mRNA-1273 Cohort and the Placebo Cohort, or after the second injection of mRNA-1273 for participants in the Placebo-mRNA-1273 Cohort<br>• COVID-19<br>• Severe COVID-19<br>• Either COVID-19 or SARS-CoV-2 infection<br>• Secondary definition of COVID-19<br>• Death caused by COVID-19<br>• COVID-19 regardless of evidence of prior SARS-CoV-2 infection determined by serologic titer against SARS-CoV-2 nucleocapsid<br>Cases after the first injection of IP for participants in the mRNA-1273 Cohort and the Placebo Cohort, or after the first injection of mRNA-1273 for participants in the Placebo-mRNA-1273 Cohort<br>• COVID-19 | • Long-term efficacy will be evaluated after the primary analysis by including data collected in Open-label Phase.<br>• Long-term efficacy data will be summarized descriptively by treatment cohort without cohort comparison.<br>• In the primary approach, cases will be counted starting 14 days after the second dose of IP for participants in treatment cohorts of mRNA-1273 and Placebo or starting 14 days after the second dose of mRNA-1273 for participants in the Placebo-mRNA-1273 Cohort.<br>• Sensitivity analyses with cases starting from the second dose, 14 days after the first dose, or first dose of IP for participants in the cohorts of mRNA-1273 and placebo, or mRNA-1273 for participants in the Placebo-mRNA-1273 Cohort will be provided.<br>• Incidences of cases assessed by numbers, rates, and 2-sided 95% CI based on the exact method adjusting for person-time will be summarized by treatment cohort.<br>• The Kaplan-Meier analysis will be used to estimate cumulative incidences of time to first cases by treatment cohort.<br>• Long-term efficacy analysis will be performed using FAS for the case of COVID-19 regardless of prior SARS- |

| | |
|---|---|
| | CoV-2 infection, and long-term efficacy analysis of other cases will be conducted using the PP Set and mITT Set.<br>• Efficacy analyses based on data collected in the Open-Label Observational Phase only will be provided in the SAP. |

**Safety:** Safety and reactogenicity will be assessed by clinical review of all relevant parameters including solicited ARs (local and systemic events), unsolicited AEs, SAEs, MAAEs, AEs leading to discontinuation, abnormal vital signs, and physical examination findings.

All safety analyses will be based on the Safety Set, except summaries of solicited ARs, which will be based on the Solicited Safety Set. All safety analyses will be provided by treatment group, and by treatment cohort as applicable, unless otherwise specified.

The number and percentage of participants with any solicited local AR, with any solicited systemic AR, and with any solicited AR during the 7-day follow-up period after each injection will be provided (Part A, the Blinded Phase only). A 2-sided 95% exact CI using the Clopper-Pearson method will be also provided for the percentage of participants with any solicited AR for each treatment group.

The number and percentage of participants with solicited ARs, unsolicited AEs, SAEs, MAAEs, Grade 3 or higher ARs and AEs, and AEs leading to discontinuation from study vaccine or participation in the study will be summarized. Unsolicited AE will be presented by MedDRA preferred term and system organ class.

For all other safety parameters, descriptive summary statistics will be provided.

Further details will be described in the statistical analysis plan (SAP).

**Immunogenicity:** The secondary immunogenicity endpoints will be analyzed using the Immunogenicity Subset by treatment group, by treatment cohort as applicable, and by baseline SARS-CoV-2 serostatus, unless otherwise specified.

The SAP will describe the complete set of immunogenicity analyses, including the approach to sample individuals into an Immunogenicity Subset for characterizing vaccine immunogenicity and assessing immunological correlates of risk and protection.

Data from quantitative immunogenicity assays will be summarized for each treatment group using positive response rates and geometric means with 95% CIs, for each timepoint for which an assessment is performed. Data from qualitative (ie, yielding a positive or negative result) assays will be summarized by tabulating the frequency of positive responses for each assay by group at each timepoint that an assessment is performed. Analyses will focus on the 2 key immunogenicity time points and the change in marker response between them: Day 1 before the first dose of IP and Day 57 (28 days after the second dose of IP). The SAP will describe the complete set of immunogenicity analyses.

Quantitative levels or geometric mean titer (GMT) of specific bAb with corresponding 95% CI at each timepoint and geometric mean fold rise (GMFR) of specific bAb with corresponding 95% CI at each post-baseline timepoint over pre-dose baseline at Day 1 will be provided by study arm. Descriptive summary statistics including median, minimum, and maximum will also be provided.

GMT of specific nAb with corresponding 95% CI at each timepoint and GMFR of specific nAb with corresponding 95% CI at each post-baseline timepoint over pre-dose baseline at Day 1 will be provided by study arm. Descriptive summary statistics including median, minimum, and maximum will also be provided. For summarizations of group variables values, antibody values reported as below the LLOQ will be replaced by $0.5 \times$ LLOQ. Values that are greater than the upper limit of quantification (ULOQ) will be converted to the ULOQ.

The number and percentage of participants with a fold rise $\geq 2$, $\geq 3$, and $\geq 4$ of serum SARS-CoV-2-specific nAb titers and participants with seroconversion due to vaccination from baseline will be provided with 2-sided 95% CI using the Clopper-Pearson method at each post-baseline timepoint. Seroconversion due to vaccination at a participant level may be defined as a change from below the LLOQ to equal or above LLOQ, or at least a 4-fold rise in terms of neutralizing antibody or vaccine antigen-specific bAb in participants with pre-existing bAb or nAb. The definition of seroconversion will be finalized and documented in the SAP prior to analyzing the immunogenicity data.

The GMT of specific nAb for each group and the geometric mean ratio (GMR) of mRNA-1273 versus placebo with corresponding 2-sided 95% CI will be estimated at each study timepoint using an analysis of covariance (ANCOVA) model with the treatment group and baseline values, if applicable, as explanatory variables, the analysis may adjust for the stratification factor.

#### 1.2. Schema

**Figure 1: Study Flow Diagram: Part A, the Blinded Phase followed by Part B, the Open-Label Observational Phase**

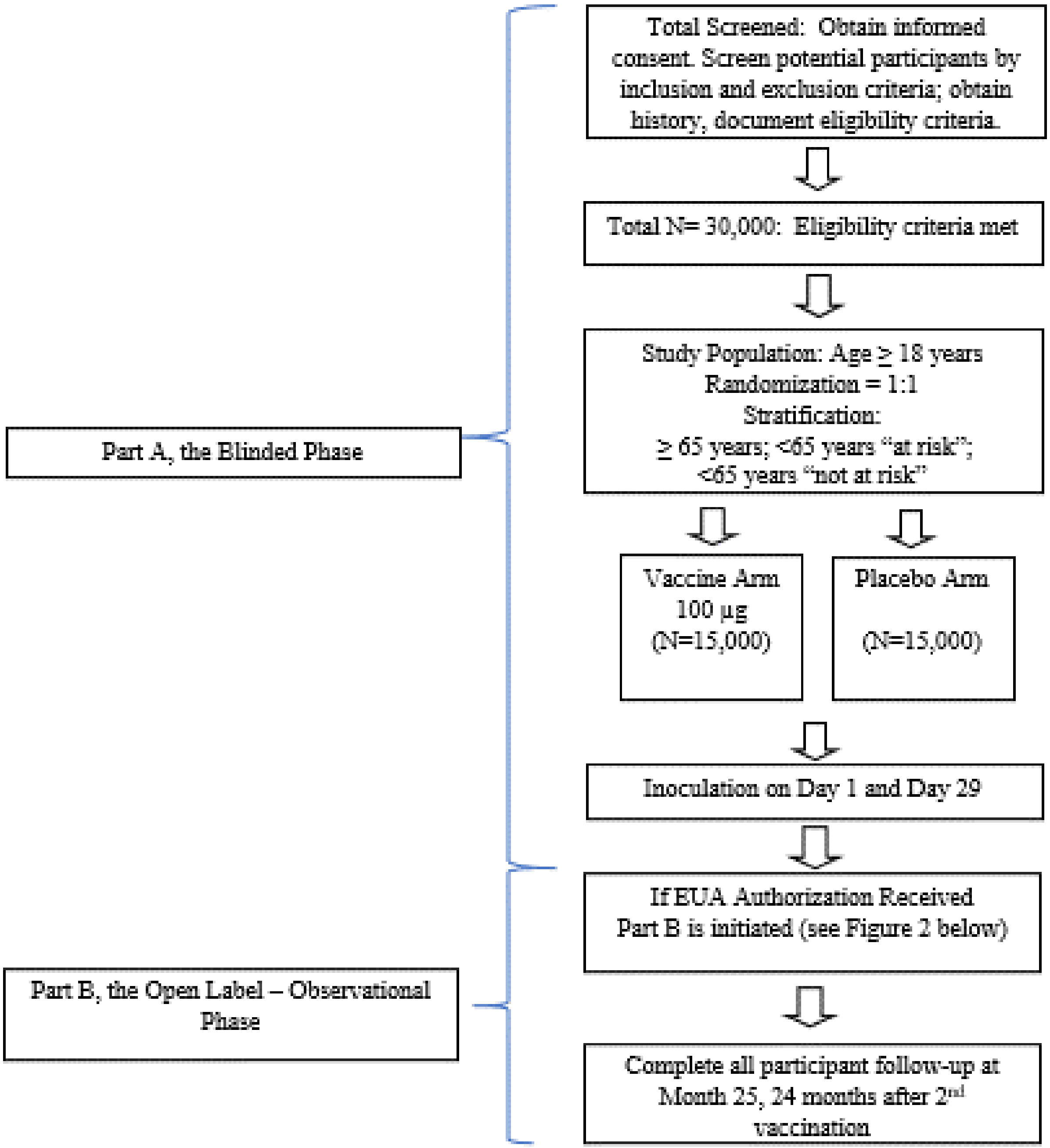

##### Figure 2: Part B, the Open-Label Observational Phase

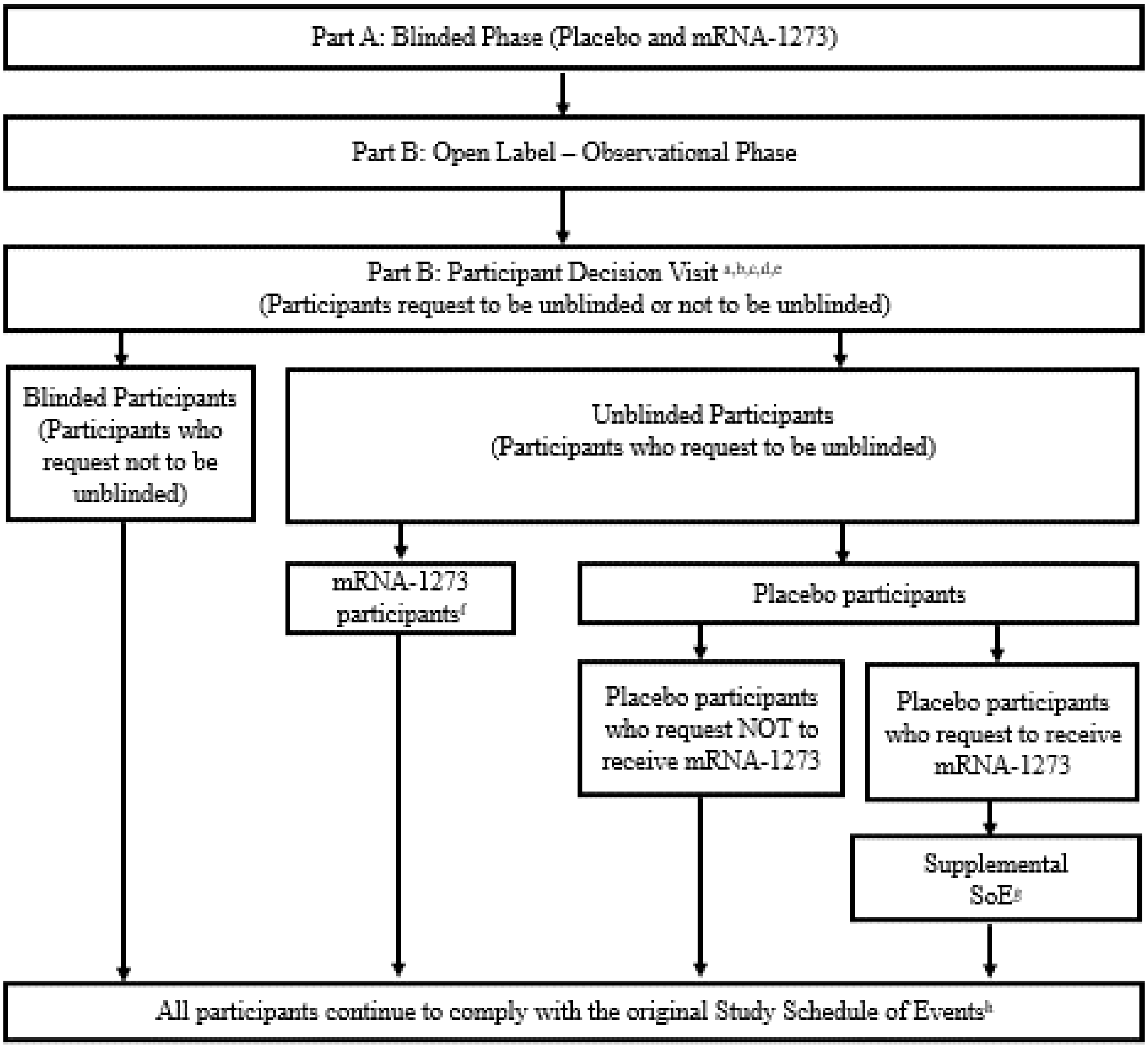

[a] All participants are encouraged to remain in the study.
[b] All participants are given the option to be unblinded to treatment received in Part A: Blinded Phase.
[c] All participants are counselled about the importance of continuing other public health measures to limit the spread of disease including physical-social distancing, wearing a mask, and hand-washing.
[d] All participants sign a revised ICF.
[e] All participants consent to provide a nasopharyngeal swab and a blood sample for immunologic analysis.
[f] mRNA-1273 recipients who only received one dose of the mRNA-1273 vaccine will receive the second dose of mRNA-1273 vaccine. Additional details provided in the Supplemental Schedule of Events (Table 22).
[g] Placebo recipients who request to receive mRNA-1273 and meet eligibility, will comply with a Supplemental Schedule of Events (Table 21) in addition to the Original Study Schedule of Events.
[h] Original Study Schedule of Events (Table 16, Table 17, Table 18, and Table 19).

### LIST OF ABBREVIATIONS

The following abbreviations and terms are used in this study protocol.

| Abbreviation | Definition |
|---|---|
| AC | Adjudication Committee |
| AE | adverse event |
| AR | adverse reaction |
| ARDS | acute respiratory distress syndrome |
| bAb | binding antibody |
| BARDA | Biomedical Advance Research and Development Authority |
| BOD | burden of disease |
| CDC | US Centers for Disease Control and Prevention |
| CFR | Code of Federal Regulations |
| CI | confidence interval |
| CLIA | Clinical Laboratory Improvement Amendments |
| CMV | cytomegalovirus |
| CONSORT | Consolidated Standards of Reporting Trials |
| CoV | coronavirus |
| COV-INF | virologically confirmed SARS-CoV-2 infection regardless of symptomology or severity |
| CRO | contract research organization |
| CSR | clinical study report |
| DHHS | Department of Health and Human Services |
| DMID | Division of Microbiology and Infectious Diseases |
| DSMB | Data and Safety Monitoring Board |
| DSPC | 1,2-distearoyl-sn-glycero-3-phosphocholine |
| eCRF | electronic case report form |
| eDiary | electronic diary |
| EUA | Emergency Use Authorization |
| ART | antiretroviral therapy |
| FAS | full analysis set |
| FDA | Food and Drug Administration |
| FSH | follicle-stimulating hormone |
| GCP | Good Clinical Practice |
| GMFR | geometric mean fold rise |
| GMP | Good Manufacturing Practice |
| GMT | geometric mean titer |

| Abbreviation | Definition |
|---|---|
| HCP | healthcare practitioner |
| HIV | Human Immunodeficiency Virus |
| hMPV | human metapneumovirus |
| HR | hazard ratio |
| HRT | hormonal replacement therapy |
| IA | interim analysis |
| IB | investigator's brochure |
| ICF | informed consent form |
| ICH | International Council for Harmonisation |
| IM | intramuscular |
| IP | investigational product |
| IRB | institutional review board |
| IRT | interactive response technology |
| LB | lower boundary |
| LLOQ | lower limit of quantification |
| LNP | lipid nanoparticle |
| LOD | limit of detection |
| LTFU | lost to follow-up |
| MAAE | medically attended adverse event |
| MedDRA | Medical Dictionary for Regulatory Activities |
| MERS-CoV | Middle East Respiratory Syndrome coronavirus |
| mRNA | messenger RNA |
| nAb | neutralizing antibody |
| NIAID | National Institute of Allergy and Infectious Diseases |
| NP | nasopharyngeal |
| PCR | polymerase chain reaction |
| PEG2000-DMG | 1-monomethoxypolyethyleneglycol-2,3-dimyristylglycerol with polyethylene glycol of average molecular weight 2000 |
| PIV3 | parainfluenza virus type 3 |
| PP | per-protocol |
| RT-PCR | reverse transcriptase polymerase chain reaction |
| S | spike |
| S-2P | spike protein with 2 proline residues introduced for stability in a prefusion conformation |
| SAE | serious adverse event |

| Abbreviation | Definition |
|---|---|
| SAP | statistical analysis plan |
| SARS-CoV | Severe Acute Respiratory Syndrome coronavirus |
| SM-102 | heptadecan-9-yl 8-((2-hydroxyethyl) (6-oxo-6-(undecyloxy) hexyl) amino) octanoate |
| SoE | Schedule of Events |
| TEAE | treatment-emergent adverse event |
| ULOQ | upper limit of quantification |
| USP | United States Pharmacopeia |
| VE | vaccine efficacy |
| WHO | World Health Organization |

### 2. INTRODUCTION

#### 2.1. Study Rationale

Coronaviruses (CoVs) are a large family of viruses that cause illness ranging from the common cold to more severe diseases, such as Middle East Respiratory Syndrome (MERS-CoV) and Severe Acute Respiratory Syndrome (SARS-CoV).

An outbreak of the CoV disease (COVID-19) caused by SARS-CoV-2 began in Wuhan, Hubei Province, China in December 2019 and has spread throughout China and to over 216 other countries and territories, including the United States (WHO 2020). On 11 March 2020, the World Health Organization (WHO) officially declared COVID-19 a pandemic. As of 28 May 2020, the WHO reported more than 5,593,631 confirmed cases and 353,334 deaths globally and the US Centers for Disease Control and Prevention (CDC) reported 1,698,523 confirmed and probable cases of COVID-19, with 100,446 deaths in the United States (CDC 2020a). The CDC have reported that the highest risk of disease burden is in older adults (≥ 65 years old) and people of any age who have serious underlying medical conditions, such as chronic lung disease or moderate to severe asthma; serious heart conditions; severe obesity; diabetes; chronic kidney disease requiring dialysis; liver disease; and those who are immunocompromised (CDC 2020b).

There is currently no vaccine against SARS-CoV-2. Global efforts to evaluate novel antivirals and therapeutic strategies to treat severe SARS-CoV-2 infections have intensified, but no proven therapeutic currently exists. Therefore, there is an urgent public health need for rapid development of novel interventions to prevent the spread of this disease. The primary goal of this Phase 3 study is to evaluate the vaccine efficacy (VE) of mRNA-1273 to prevent COVID-19, compared to placebo.

#### 2.2. Background and Overview

ModernaTX, Inc. (the Sponsor) has developed a rapid-response, proprietary vaccine platform based on a messenger RNA (mRNA) delivery system. The platform is based on the principle and observations that cells in vivo can take up mRNA, translate it, and then express protein viral antigen(s) on the cell surface. The delivered mRNA does not enter the cellular nucleus or interact with the genome, is nonreplicating, and is expressed transiently. mRNA vaccines have been used to induce immune responses against infectious pathogens such as cytomegalovirus (CMV) (NCT03382405), human metapneumovirus (hMPV) and parainfluenza virus type 3 (PIV3) (NCT03392389), Zika virus (NCT03325075), and influenza virus (NCT03076385 and NCT03345043).

The Sponsor is using its mRNA-based platform to develop a novel lipid nanoparticle (LNP)-encapsulated mRNA-based vaccine against SARS-CoV-2 (mRNA-1273). mRNA-1273 encodes for the full-length spike (S) protein of SARS-CoV-2, modified to introduce 2 proline residues to stabilize the S protein (S2P) in a prefusion conformation. The CoV S protein mediates attachment and entry of the virus into host cells (by fusion), making it a primary target for neutralizing antibodies that prevent infection (Johnson et al 2016; Wang et al 2015; Wang et al 2018; Chen et al 2017; Corti et al 2015; Yu et al 2015; Kim et al 2019; Widjaja et al 2019). It has been confirmed that the stabilized SARS-CoV-2 S2P antigen presents in the correct prefusion conformation (Wrapp et al 2020).

The development of the mRNA-1273 vaccine is being accelerated to address the current SARS-CoV-2 outbreak as a result of the uniquely rapid and scalable manufacturing process for mRNA-1273. The primary goal of this Phase 3 study is to evaluate the VE of mRNA-1273 to prevent COVID-19, compared to placebo.

##### 2.2.1. Nonclinical Studies

Nonclinical studies have demonstrated that CoV S proteins are immunogenic and S protein-based vaccines, including those based on mRNA delivery platforms, are protective in animals. Prior clinical studies of vaccines targeting related CoVs and other viruses have demonstrated that mRNA-based vaccines are safe and immunogenic. It is therefore anticipated that mRNA-1273 will generate robust immune responses to the SARS-CoV-2 S protein and will be well tolerated. In addition, mRNA-1273 has shown preliminary evidence of protection against SARS-CoV-2 in a murine model of infection (data on file).

In support of development of mRNA-1273 for prophylaxis against SARS-CoV-2 infection, nonclinical immunogenicity, biodistribution, and safety studies have been completed with similar mRNA-based vaccines formulated in LNPs containing SM-102 (heptadecan-9-yl 8 ((2 hydroxyethyl)(6 oxo 6-(undecyloxy)hexyl)amino)octanoate), the novel proprietary lipid used in the mRNA-1273 LNP formulation.

A detailed review of nonclinical experience with mRNA-1273 vaccine is provided in the Investigator's Brochure (IB).

##### 2.2.2. Clinical Studies

The mRNA-1273 vaccine is currently being evaluated for safety and immunogenicity in a dose-ranging Phase 1 study (NCT04283461) sponsored and conducted by the Division of Microbiology and Infectious Diseases (DMID) of the National Institute of Allergy and Infectious Diseases (NIAID). The Phase 1 DMID study is an open-label dose-ranging study of mRNA-1273 in healthy adult male and non-pregnant female participants in 3 age groups: age 18 to 55 years, inclusive (45 participants); age 56 to 70 years, inclusive (30 participants); and ≥ 71 years (30 participants). Each participant will receive an IM injection (0.5 mL) of mRNA-1273 on Days 1 and 29 in the deltoid muscle and will be followed for 13 months after the second injection.

As of 26 October 2020, 120 participants were enrolled. All participants received the first injection of mRNA-1273, and 116 participants received the second injection. Three mRNA-1273 dose levels (25, 100, and 50 µg) administered 28 days apart have been assessed in participants 18 to 55 years, 56 to 70 years, and ≥ 71 years of age. The mRNA-1273 250-µg dose was not evaluated in participants 56 to 70 years and ≥ 71 years of age due to reactogenicity observed in 4 participants in the 250 µg (18 to 55 years) dose cohort. Across all age groups, solicited adverse reactions (ARs) were predominantly mild or moderate in severity and most commonly included fatigue, chills, headache, myalgia, and pain at the injection site. The incidence and severity of solicited ARs were generally dose dependent and increases in incidence and severity were generally observed after the second injection. The incidence of systemic reactions increased after the second injection, particularly at the highest (250 µg) dose. Four (27%) participants in the 250 µg dose cohort reported at least 1 severe solicited AR after the second injection, including feverishness, fatigue, fever, headache, myalgia, nausea, and erythema/redness. No serious adverse events (SAEs) have been reported through Day 119, and no pause rules were triggered during the study.

Additionally, a dose-finding Phase 2a study (mRNA-1273-P201) conducted by the Sponsor under IND 19745 will expand the safety and immunogenicity database by testing 2 two dose levels (50 µg and 100 µg) in 400 adults. As of 05 November 2020, 200 (100%) participants each in the mRNA-1273 50 µg group, mRNA-1273 100 µg group, and placebo group received the first dose, and 195 (97.5%) participants in the mRNA-1273 50 µg group, 198 participants (99.0%) in the mRNA-1273 100 µg group, and 194 (97.0%) participants in the placebo group received the second injection (mRNA-1273-P201). In this study, 2 mRNA-1273 dose levels (50 and 100 µg) or placebo administered 28 days apart were assessed in participants ≥ 18 to < 55 years and ≥ 55 years of age. No protocol-specified pause rules were met during the study. In addition, the study initiated with a sentinel group of 50 participants ≥ 55 years of age; after review of the safety data by the independent Safety Monitoring Committee, the cohort was expanded to include the remaining 250 participants ≥ 55 years of age.

In the Phase 3 study (mRNA-1273-P301), as of 25 October 2020, a total of 30,350 participants have been dosed in the Blinded Phase of the Study (15,184 received mRNA-1273 and 15,165 received placebo). The protocol-defined interim analysis (IA) was conducted in November 2020, which indicated that the primary efficacy endpoint in Study mRNA-1273-P301 was met: mRNA-1273 prevented COVID-19 starting 14 days after the second dose of the mRNA-1273 vaccine, based on a total of 95 adjudicated cases accrued (5 cases in the mRNA-1273 group and 90 cases in the placebo group). The VE was 94.5% (95% CI: 86.5%, 97.8%; 1-sided p-value < 0.0001), rejecting the null hypothesis of VE ≤ 30% and achieving the prespecified efficacy boundary based on the 1-sided nominal alpha of 0.0047 using the Lan-DeMets O'Brien-Fleming spending function. The mRNA-1273 vaccine was effective in preventing severe COVID-19, with 11 cases in the placebo group and 0 cases in the mRNA-1273 group. In addition, the vaccine was efficacious in preventing COVID-19 regardless of prior SARS-CoV-2 infection for cases starting 14 days after the second injection (VE of 93.5% based on hazard ratio [HR]).

In this study, the safety and reactogenicity of mRNA-1273 100 µg compared with placebo administered 28 days apart were assessed in participants 18 years of age and older at increased risk for acquiring COVID-19 based on occupation or location and living circumstances. Reactogenicity (solicited local and/or systemic ARs) was observed in the majority of participants in the mRNA-1273 group and generally increased after the second injection. The rates of local and systemic ARs were higher in the mRNA-1273 group than in the placebo group after each injection. The majority of solicited ARs in the mRNA-1273 group were grade 1 to grade 2 in severity and generally resolved within 3 days or less. The incidence of unsolicited treatment-emergent adverse events (TEAEs), severe TEAEs, and medically attended adverse events (MAAEs) during the 28 days after injection was also generally similar in participants who received mRNA-1273 and those who received placebo.

Deaths and SAEs were generally reported at a similar incidence in the mRNA 1273 and placebo groups. There was no evidence of enhanced disease, as fewer cases of severe COVID-19 and COVID-19 were observed in participants who received mRNA-1273 than in those who received placebo.

#### 2.3. Benefit/Risk Assessment

##### 2.3.1. Potential Benefits of Study Participation

The target study population for this study is adults with no known history of SARS-CoV-2 infection but whose locations or circumstances put them at high risk of COVID-19. The following benefits may accrue to participants:

- The mRNA-1273 vaccine may be an effective vaccine against COVID-19.
- A baseline (Day 1) evaluation for SARS-CoV-2 infection and ongoing surveillance for COVID-19 throughout the study.
- Contributing to the development of a vaccine against COVID-19, a current pandemic disease.

##### 2.3.2. Risks from Study Participation

Immediate systemic allergic reactions (eg, anaphylaxis) can occur following any vaccination. These reactions are very rare and are estimated to occur once per 450,000 vaccinations for vaccines that do not contain allergens such as gelatin or egg protein (Zent et al 2002). As a precaution, all participants will remain under observation at the study site for at least 30 minutes after vaccination.

Vasovagal syncope (fainting) can occur before or after any vaccination, is usually triggered by the pain or anxiety caused by the injection and is not related to the substance injected. Therefore, it is important that standard precautions and procedures be followed to avoid injury from fainting.

Intramuscular injection with other mRNA vaccines manufactured by the Sponsor containing the SM-102 lipid formulation commonly results in a transient and self-limiting local inflammatory reaction. This typically includes pain, erythema (redness), or swelling (hardness) at the injection site, which are mostly mild-to-moderate in severity and usually occur within 24 hours of vaccination. More severe, but self-limited, local reactions, erythema and induration, have been observed at dose of mRNA-1273 exceeding the dose proposed in this study.

Most systemic adverse events (AEs) observed after vaccination do not exceed mild-to-moderate severity. The most commonly reported systemic ARs are anticipated to be fever, fatigue, chills, headache, myalgias and arthralgias. More severe reactions, including erythema, induration, fever, headache and nausea, were reported after receiving doses of mRNA-1273 that were greater than the dose proposed for use in this study. In all cases, the reactions resolved spontaneously.

Laboratory abnormalities (including increases in liver functional tests and serum lipase levels) following vaccination were observed in clinical studies with similar mRNA-based vaccines. These abnormalities were without clinical symptoms or signs and returned toward baseline (Day 1) values over time. The clinical significance of these observations is unknown. Further details are provided in the current IB.

There is a theoretical risk that active vaccination to prevent the novel viral infection caused by SARS-CoV-2 may cause a paradoxical increase in the risk of disease. This possibility is based on the rare phenomenon of vaccine-associated disease enhancement which was first seen in the 1960s with 2 vaccines made in the same way (formalin-inactivated whole virus) and designed to protect children against infection with RSV (Chin et al 1969) or measles (Fulginiti et al 1967). Disease enhancement has also been proposed as a possible explanation for cases of more serious disease associated with dengue vaccination (Thomas and Yoon 2019; WHO 2018). It is not known if mRNA-1273 will increase the risk of enhanced disease.

To monitor the risk of enhanced disease in this study, an independent Data and Safety Monitoring Board (DSMB) will review unblinded cases of COVID-19 to assess for inefficacy and also for numerical imbalance in cases of both COVID-19 and severe COVID-19 with the purpose of providing a non-binding recommendation to the Sponsor (Section 8.4.2).

##### 2.3.3. Overall Benefit/Risk Conclusion

All participants will be included based on their increased risk of SARS-CoV-2 infection. Accordingly, all will benefit from baseline and ongoing evaluations for SARS-CoV-2 infection.

Since this is a placebo-controlled study (Section 4), half the participants will have the potential to receive mRNA-1273 vaccine, the efficacy of which is unknown at present. Vaccination with mRNA-1273 may not prevent COVID-19 in all vaccines.

Participants who receive placebo as part of this study may have an opportunity cost of not being treated with another investigational vaccine against COVID-19.

The placebo for this study is a saline solution, without any LNP. Thus, participants receiving saline may be at lower risk of AEs related to injection than participants receiving mRNA-1273.

Safety findings will be monitored and periodically reviewed by the DSMB to evaluate the safety and treatment status of all participants. The DSMB will review and assess the safety data as described in Section 8.4.2.

Considering the lack of approved vaccines for COVID-19, the participants' risk of COVID-19 outside the study, and the nonclinical and clinical data to date, the Sponsor considers the potential benefits of participation to exceed the risks.

Based on the interim results from the pivotal Phase 3 study, mRNA-1273 prevents COVID-19 and severe COVID-19. The demonstrated clinical benefit of mRNA-1273 is supported by evidence of a robust immune response both in terms of binding antibodies (bAbs) and neutralizing antibodies (nAbs) as well as the induction of CD4+ T-cells with a Th-1 dominant phenotype. Based on administration of mRNA-1273 to approximately 15,693 adults across all 3 clinical studies to date, there have been no emergent safety concerns and the AE profile is manifested largely by mild to moderate reactogenicity lasting 2 to 3 days.

### 3. OBJECTIVES AND ENDPOINTS

#### Table 1: Objectives and Endpoints

| **Objectives and Endpoints** | |
|---|---|
| **Primary Objective** | **Primary Endpoints** |
| **Efficacy Objective (Primary):**<br>To demonstrate the efficacy of mRNA-1273 to prevent COVID-19. | **Efficacy Endpoints (Primary):** Vaccine efficacy of mRNA-1273 to prevent the first occurrence of COVID-19 starting 14 days after the second dose of investigational product (IP), where COVID-19 is defined as symptomatic disease based on the following criteria:<br>• The participant must have experienced at least TWO of the following systemic symptoms: Fever (≥ 38ºC), chills, myalgia, headache, sore throat, new olfactory and taste disorder(s), OR<br>• The participant must have experienced at least ONE of the following respiratory signs/symptoms: cough, shortness of breath or difficulty breathing, OR clinical or radiographical evidence of pneumonia; AND<br>• The participant must have at least one NP swab, nasal swab, or saliva sample (or respiratory sample, if hospitalized) positive for SARS-CoV-2 by RT-PCR |
| **Safety Objective (Primary):**<br>To evaluate the safety and reactogenicity of 2 injections of the mRNA-1273 vaccine given 28 days apart. | **Safety Endpoint (Primary):**<br>• Solicited local and systemic ARs through 7 days after each dose of IP (Part A: Blinded Phase only).<br>• Unsolicited AEs through 28 days after each dose of IP (Part A: Blinded Phase only).<br>• MAAEs or AEs leading to withdrawal through the entire study period.<br>• SAEs throughout the entire study period. |

| **Objectives and Endpoints** | |
|---|---|
| **Efficacy Objectives (Secondary)** | **Efficacy Endpoints (Secondary)** |
| To evaluate the efficacy of mRNA-1273 to prevent severe COVID-19 | Vaccine efficacy of mRNA-1273 to prevent severe COVID-19, defined as first occurrence of COVID-19 starting 14 days after the second dose of IP, (as per the primary endpoint) AND any of the following:<br>• Clinical signs indicative of severe systemic illness, Respiratory Rate ≥ 30 per minute, Heart Rate ≥ 125 beats per minute, $SpO_2 \leq 93\%$ on room air at sea level or $PaO_2/FIO_2 < 300$ mm Hg, OR<br>• Respiratory failure or Acute Respiratory Distress Syndrome (ARDS), (defined as needing high-flow oxygen, non-invasive or mechanical ventilation, or ECMO), evidence of shock (systolic blood pressure < 90 mmHg, diastolic BP < 60 mmHg or requiring vasopressors), OR<br>• Significant acute renal, hepatic or neurologic dysfunction, OR<br>• Admission to an intensive care unit or death |
| To evaluate the efficacy of mRNA-1273 to prevent serologically confirmed SARS-CoV-2 infection or COVID-19 regardless of symptomatology or severity. | Vaccine efficacy of mRNA-1273 to prevent the first occurrence of either COVID-19 or SARS-CoV-2 infection starting 14 days after the second IP dose.<br>This endpoint is a combination of COVID-19, defined as for the primary endpoint, and asymptomatic SARS-CoV-2 infection, determined by seroconversion assessed by bAb levels against SARS-CoV-2 as measured by a ligand-binding assay specific to the SARS-CoV-2 nucleocapsid protein and with a negative nasopharyngeal (NP) swab sample for SARS-CoV-2 at Day 1 (Section 8.1.1). |

| **Objectives and Endpoints** | |
|---|---|
| To evaluate VE against a secondary definition of COVID-19. | Vaccine efficacy of mRNA-1273 to prevent the secondary case definition of COVID-19 starting 14 days after the second IP dose.<br>The secondary case definition of COVID-19 is defined as the following systemic symptoms: fever (temperature ≥ 38ºC), or chills, cough, shortness of breath or difficulty breathing, fatigue, muscle aches or body aches, headache, new loss of taste or smell, sore throat, nasal congestion or rhinorrhea, nausea or vomiting, or diarrhea AND a positive NP swab, nasal swab, or saliva sample (or respiratory sample, if hospitalized) for SARS-CoV-2 by RT-PCR |
| To evaluate VE to prevent death caused by COVID-19. | Vaccine efficacy of mRNA-1273 to prevent death due to a cause directly attributed to a complication of COVID-19, starting 14 days after the second IP dose. |
| To evaluate the efficacy of mRNA-1273 to prevent COVID-19 after the first dose of IP. | Vaccine efficacy of mRNA-1273 to prevent the first occurrence of COVID-19 starting 14 days after the first dose of IP. |
| To evaluate the efficacy of mRNA-1273 to prevent COVID-19 in all study participants, regardless of evidence of prior SARS-CoV-2 infection. | Vaccine efficacy of mRNA-1273 to prevent the first occurrence of COVID-19 starting 14 days after the second dose of IP regardless of evidence of prior SARS-CoV-2 infection determined by serologic titer against SARS-CoV-2 nucleocapsid (FAS analysis population, see Section 9.4). |
| To evaluate the efficacy of mRNA-1273 to prevent asymptomatic SARS-CoV-2 infection. | Vaccine efficacy to prevent the first occurrence of SARS-CoV-2 infection in the absence of symptoms defining COVID-19 starting 14 days after the second IP dose.<br>SARS-CoV-2 infection determined by seroconversion assessed by bAb levels against SARS-CoV-2 as measured by a ligand-binding assay specific to the SARS-CoV-2 nucleocapsid protein and with a negative NP swab sample for SARS-CoV-2 at Day 1 (Section 8.1.1). |

| Objectives and Endpoints | |
|---|---|
| **Immunogenicity Objective (Secondary):** | **Immunogenicity Endpoints (Secondary):** |
| To evaluate the immunogenicity of 2 doses of mRNA-1273 given 28 days apart. | • Geometric mean titer (GMT) of SARS-CoV-2 -specific neutralizing antibody (nAb) on Day 1, Day 29, Day 57, Day 209, Day 394, and Day 759.<br>• Geometric mean fold rise (GMFR) of SARS-CoV-2-specific nAb relative to Day 1 on Day 29, Day 57, Day 209, Day 394, and Day 759.<br>• Quantified levels or GMT of S protein-specific binding antibody (bAb) on Day 1, Day 29, Day 57, Day 209, Day 394, and Day 759.<br>• GMFR of S protein -specific bAb relative to Day 1 on Day 29, Day 57, Day 209, Day 394, and Day 759. |
| **Exploratory Objectives** | |
| To evaluate the effect of mRNA-1273 on the viral infection kinetics as measured by viral load at SARS-CoV-2 infection diagnosis by RT-PCR and number of days from the estimated date of SARS-CoV-2 infection until undetectable SARS-CoV-2 infection by RT-PCR. | |
| To assess VE to reduce the duration of symptoms of COVID-19. | |
| To evaluate VE against all-cause mortality. | |
| To assess VE against burden of disease (BOD) due to COVID-19. | |
| To evaluate the genetic and/or phenotypic relationships of isolated SARS-CoV-2 strains to the vaccine sequence. | |
| To evaluate immune response markers after dosing with IP as correlates of risk of COVID-19 and as correlates of risk of SARS-CoV-2 infection. | |
| To conduct additional analyses related to furthering the understanding of SARS-CoV-2 infection and COVID-19, including analyses related to the immunology of this or other vaccines, detection of viral infection, and clinical conduct. | |

Abbreviations: AE = adverse event; AR = adverse reaction; bAb = binding antibody; FAS = full analysis set; GMFR = geometric mean fold rise; GMT = geometric mean titer; ICU = intensive care unit; MAAE = medically attended adverse event; nAb = neutralizing antibody; NP = nasopharyngeal; SAE = serious adverse event; VE = vaccine efficacy.

### 4. STUDY DESIGN

#### 4.1. General Design

This is a two-part Phase 3 study: Part A and Part B. Participants in Part A, the Blinded Phase of this study are blinded to their treatment assignment. Given that the primary efficacy endpoint for mRNA-1273 against COVID-19 was met per the protocol-defined IA, Part B, the Open-Label Observational Phase of this study is designed to offer eligible participants who received placebo in Part A of this study, an option to request open-label mRNA-1273 (as long as investigational vaccine is available) (Figure 1).

All participants may have up to 8 scheduled clinic visits, including Screening, Day 1, Day 29, Day 57, Day 209, Day 394, Day 759, and the Participant Decision Visit as specified in Schedule of Events (SoEs) (Table 16, Table 17, Table 18, Table 19, and Table 20). Participants eligible for Part B OL dosing may have up to 3 additional scheduled clinic visits as specified in SoEs (Table 21 and Table 22).

This study will be conducted in compliance with the protocol, Good Clinical Practice (GCP), and all applicable regulatory requirements.

##### 4.1.1. Part A, the Blinded Phase

The Blinded Phase of this study is a randomized, stratified, observer-blind, placebo-controlled evaluation of the efficacy, safety, and immunogenicity of mRNA-1273 SARS-CoV-2 vaccine compared to placebo in adults 18 years of age and older who have no known history of SARS-CoV-2 infection but whose locations or circumstances put them at appreciable risk of acquiring COVID-19 and/or SARS-CoV-2 infection. Figure 1 shows the study flow and Appendix 1 (Table 16, Table 17, Table 18, and Table 19) displays the planned Schedules of Events.

Approximately 30,000 participants will be randomly assigned to receive doses of either 100 µg of mRNA-1273 vaccine or a placebo control in a 1:1 randomization ratio. Assignment will be stratified by age and health risk (Section 6.2.1.1). This is a case-driven study and thus final sample size of the study will depend on the actual attack rate of COVID-19.

All participants will be assessed for efficacy and safety endpoints and provide a NP swab sample and blood sample before the first and second dose of IP, in addition to a series of post-dose blood samples for immunogenicity through 24 months after the second dose of IP. Efficacy assessments will include surveillance for COVID-19 with RT-PCR confirmation of SARS-CoV-2 infection after the first and second dose of IP. As noted above, this is a case-driven study: if the prespecified criteria for early efficacy are met at the time of either IA or overall efficacy at the primary analysis, a final study report describing the efficacy and safety of mRNA-1273 will be prepared based on the data available at that time. In the event that success criteria are met either at the time of the IA or when the total number of cases toward the primary endpoint have accrued, participants will continue to be followed in a blinded fashion until Month 25 to enable assessment of long-term safety and durability of VE (Figure 3). If the study concludes early, all participants will be requested to provide a final blood sample at the time of study conclusion.

Each participant will receive 2 doses of IP by 0.5 mL intramuscular (IM) injection, the first on Day 1 and the second on Day 29. An NP swab sample will be collected prior to the first and second dose of IP, for evaluation by RT-PCR. To preserve observer blinding, only delegated unblinded study personnel responsible for study vaccine preparation, administration and/or accountability will have knowledge of study treatment assignment (Section 6.2.8.1).

Participants will be given an electronic diary (eDiary) to report solicited ARs for 7 days after each dose of IP (Part A, Blinded Phase only) and to prompt an unscheduled clinic visit for clinical evaluation and NP swab sample if a participant experiences any symptoms of COVID-19. Participants will use the eDiary to report solicited ARs for 7 days after each dose of IP (Part A, Blinded Phase only) and weekly eDiary prompts (every 7 days) to elicit an unscheduled Illness Visit if the participant is experiencing COVID-19 symptoms. All participants will receive safety calls on Day 8, Day 15, Day 22, Day 36, and Day 43 that will serve both to monitor for unsolicited AEs and to monitor for symptoms of COVID-19.

Safety telephone calls and eDiary safety prompts will be performed in conjunction with surveillance for COVID-19 according to the SoEs (Table 16, Table 17, Table 18, and Table 19) and are intended to capture SAEs, MAAEs, AEs leading to withdrawal, concomitant medications associated with these events, receipt of non-study vaccinations, and pregnancy (Section 8.2.1). If an eDiary prompt results in identification of a relevant safety event, a follow-up safety call will be triggered.

Surveillance for COVID-19 will be performed through weekly contacts with the participant via a combination of telephone calls and completion of an eDiary starting at Day 1 through the end of the study (Section 8.1.2). Participants with symptoms of COVID-19 lasting at least 48 hours (except for fever and/or respiratory symptoms) will return to the clinic or will be visited at home by medically qualified site staff within 72 hours to collect an NP swab sample for RT-PCR testing for SARS-CoV-2 and other respiratory pathogens, or alternatively, if a clinic or home visit is not possible, will submit a saliva (or nasal swab) sample for SARS-CoV-2 RT-PCR testing (Section 8.1.1).

All study participants who experience COVID-19 symptoms and subsequently present for an Illness Visit (in-clinic or at home) will be given an instruction card listing symptoms and severity grading system along with a thermometer, an oxygen saturation monitor, and saliva collection tubes. The list of symptoms is presented in Section 8.1.2 and the severity scoring system is presented in Section 8.1.3. Study participants will be contacted by the investigator (or appropriately delegated study staff) daily with telemedicine visits through Day 14 or until symptoms have resolved, whichever is later. During the telemedicine visit (preferably done in the evening), the participant will be asked to verbally report the severity of each symptom, their highest body temperature and lowest oxygen saturation for that day, and the investigator will determine if medical attention is required due to worsening of COVID-19 symptoms (Table 19). Study participants will collect their own saliva (or nasal swab) sample on 3, 5, 7, 9, 14, and 21 days after the initial Illness Visit meeting criteria for COVID-19 (defined as the date of onset of symptoms and positive virologic test). Finally, a convalescent visit will be scheduled approximately 28 days after the initial Illness Visit. At this visit, a saliva (or nasal swab) sample will be collected and a blood sample will be drawn for immunologic assessment of SARS-CoV-2 infection.

At each dosing visit, participants will be instructed (Day 1) or reminded (Day 29) on how to document and report solicited ARs in the eDiary provided. Solicited ARs will be assessed for 7 days after each IP dose (Part A, Blinded Phase only) and unsolicited AEs will be assessed for 28 days after each IP dose (Part A, Blinded Phase only); SAEs, MAAEs, AEs leading to withdrawal, will be assessed throughout the study.

Participants may experience AEs that necessitate an unscheduled visit. There may also be situations in which the investigator asks a participant to report for an unscheduled visit following the report of an AE. Additional examinations may be conducted at these visits as necessary to ensure the safety and well-being of participants during the study. Electronic case report forms (eCRFs) should be completed for each unscheduled visit.

##### 4.1.2. Part B, the Open-label Observational Phase

The Part B, the Open-Label Observational Phase of the study is prompted by the authorization of a COVID-19 vaccine under EUA. Transitioning the study to Part B permits all ongoing study participants to be informed of the availability and eligibility criteria of any COVID-19 vaccine made available under an EUA and the option to offer all ongoing study participants who request unblinding, an opportunity to schedule a Participant Decision clinic visit to know their original treatment assignment (placebo vs. mRNA-1273 vaccine).

Part B, the Open-Label Observational Phase of the study, also provides the opportunity for eligible study participants who previously received placebo, to actively request to receive 2 doses of mRNA-1273 vaccine (as long as investigational vaccine is available). All study participants will receive a Notification Letter and will be asked to schedule a Participant Decision clinic visit. Principal Investigators should consider current local public health guidance for administration of COVID-19 vaccines under EUA when determining the scheduling priority of participants.

With regards to eligibility in Part B, the CDC-EUA guidance specifications will supersede eligibility criteria in the protocol.

All participants will proceed to Part B, the Open-Label Observational Phase of the study, starting with a Participant Decision clinic visit (Figure 2).

At the Participant Decision clinic visit, all participants will:

- Be encouraged to remain in the ongoing study,
- Be given the option to be unblinded as to their original group assignment (placebo vs. mRNA-1273 vaccine),
- Be counselled about the importance of continuing other public health measures to limit the spread of disease including social distancing, wearing a mask, and hand-washing,
- Sign a revised informed consent form (ICF),
- Provide a NP swab for RT-PCR for SARS-CoV-2 and a blood sample for serology prior to unblinding.

Figure 2 shows the Part B Study Flow schematic.

The following participants will continue with the original study SoEs presented in Table 16, Table 17, Table 18, and Table 19 (Figure 2):

- participants who request to not be unblinded,
- participants who request to be unblinded and received mRNA-1273, and
- participants who request to be unblinded and received placebo and choose to remain on placebo.

Participants who are unblinded, received placebo, are eligible, and request to receive mRNA-1273 (as long as investigational vaccine is available) (Figure 2), will have the following clinic visits as shown in the Supplemental SoE (Table 21):

- Open-label Dose 1 (OL-D1): To occur at the Participant Decision clinic visit or at a scheduled subsequent visit.
- Open-label Dose 2 (OL-D29): To occur 28 days after Dose 1 on OL-D1.
- Open-label Clinic Visit (OL-D57): To occur 1-month after Dose 2 on OL-D29.
- And in addition, continue to comply with the Original Study Schedule of Events (Table 16, Table 17, Table 18, and Table 19) as applicable.

Participants who are unblinded, received ONLY 1 dose of mRNA-1273:

- They had a dosing error in Part A of the study that resulted in 1 dose of mRNA-1273 and 1 dose of placebo being administered.
- They did not have an AE that contraindicated the second dose of mRNA-1273 in Part A of the study.
- They did not withdraw consent in Part A of the study.
- They did not complete their second dose in Part A of the study for reasons other than the above.

If participants are eligible, and request to receive mRNA-1273 (Figure 2) will proceed to have the following clinic visits as shown in the Modified Supplemental Schedule of Events (Table 22):

- Their second dose on OL-D1: To occur at the Participant Decision clinic visit or at a scheduled subsequent visit.
- Open-label Clinic Visit (OL-D29): To occur 1-month after Dose 2 on OL-D1.
- And in addition, continue to comply with the Original Study Schedule of Events (Table 16, Table 17, Table 18, and Table 19) as applicable.

All participants remain on their original SoE to complete 24 months of follow-up after Dose 2; study participants who enter the Supplemental/Modified Supplemental SoE as described above do so in addition to their original SoE. Accordingly, all study participants will complete the full study follow-up to 24 months after the second dose of their original inoculation (mRNA-1273 or placebo).

At the point when the mRNA-1273 vaccine is no longer available for study use, any participant who schedules a Participant Decision Visit after this time-point will be unblinded (with no option to stay blinded), but will not be offered to receive mRNA-1273 study vaccine. These participants will have all of the same procedures performed outlined in Table 20 with the exception of being offered to receive mRNA-1273 vaccine There will be no scientific metric or ethical advantage for participants to remain blinded, when the majority of the study has already been unblinded. If the participant wants to receive EUA vaccine outside of the study after being unblinded, they will need to be withdrawn from the study.

After the Biologics License Application (BLA) database lock, any remaining participants who have not been unblinded will be unblinded by the Moderna study team and will receive their unblinding treatment assignment (placebo vs. mRNA vaccine) via certified letter from the site. These participants will not be required to return to the site for a NP swab or blood draw.

**Figure 3: Event/Case-Driven Study with 24 Months of Planned Follow-Up**

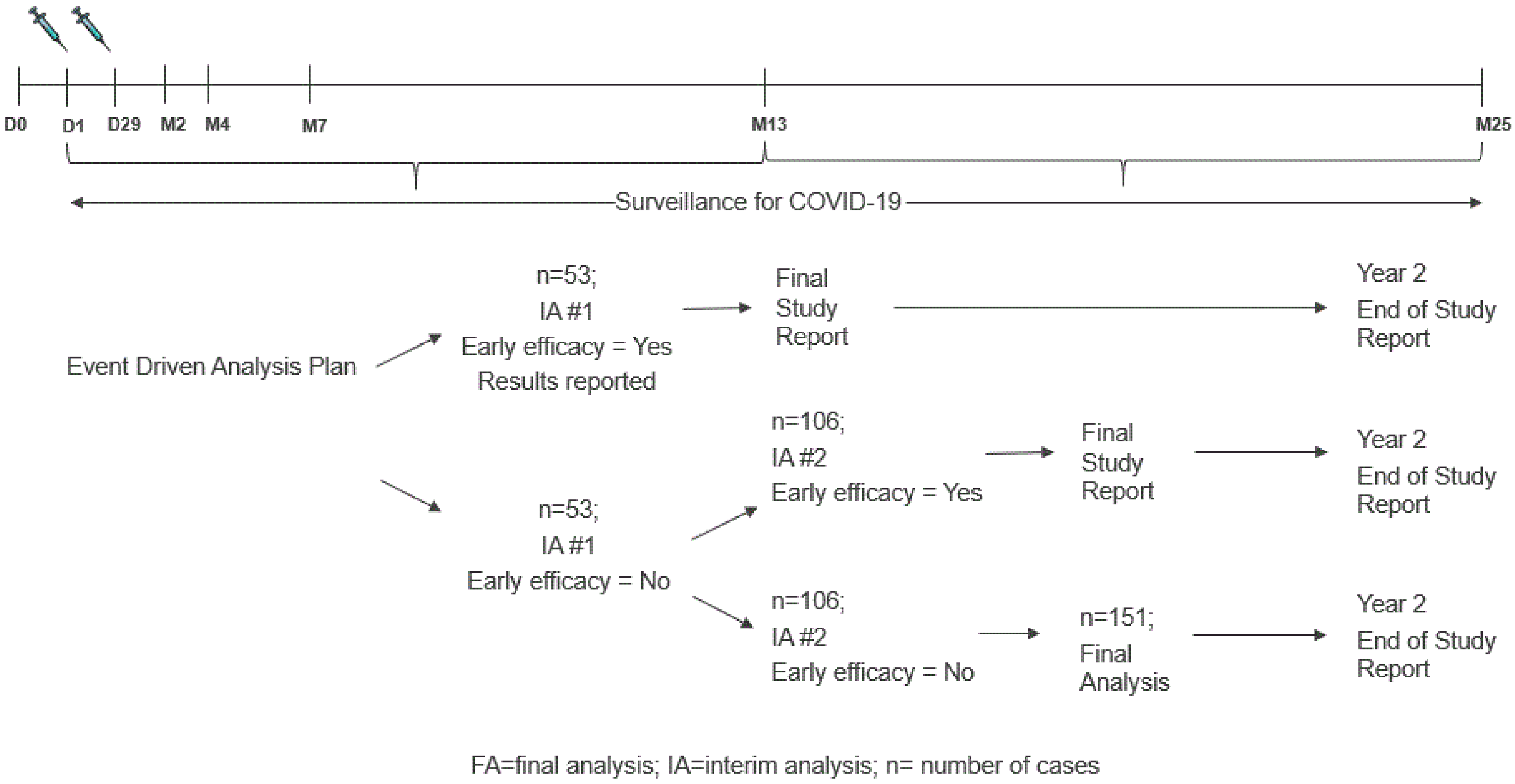

##### 4.2. Scientific Rationale for Study Design

The mRNA-1273 vaccine is being developed to prevent COVID-19, the disease resulting from SARS-CoV-2 infection. The study is designed to primarily evaluate the clinical efficacy and safety of mRNA-1273 to prevent COVID-19 for up to 2 years after the second dose of mRNA-1273. The immunogenicity endpoints and detection of SARS-CoV-2 infection are secondary objectives.

The design and focus of the study are dependent on the current COVID-19 Pandemic, requiring identification of participant candidates at high risk of SARS-CoV-2 infection. The Sponsor may adjust the size of the study or duration of follow-up based on the blinded review of the total number of cases of COVID-19 accrued during the study, in addition to estimated percentages of study participants with immunologic evidence of SARS-CoV-2 infection at baseline. If achieving 151 cases of COVID-19 is manifestly unattainable based on plausible expansions of sample size or increased follow-up, an analysis of blinded data may be performed and change of the study design such as changing lower bound or the endpoint may be proposed.

This Phase 3 study was designed to be a randomized, stratified, observer-blind, placebo controlled-study to evaluate the efficacy, safety, and immunogenicity of mRNA-1273 SARS-CoV-2 vaccine compared to placebo in adults 18 years of age and older who have no known history of SARS-CoV-2 infection but whose locations or circumstances put them at appreciable risk of acquiring COVID-19 and/or SARS-CoV-2 infection.

Following authorization of a COVID-19 vaccine under EUA, the study design was previously amended to include a transition to Part B, the Open-Label Observational Phase (Figure 2). The demonstration of efficacy against COVID-19, and satisfactory safety data, at a time when the COVID-19 pandemic remains critical, warranted allowing those study participants who actively request to know their original assignment (placebo or mRNA-1273 vaccine) on study. Transitioning the study to Part B, the Open-Label Observational Phase, permits all ongoing study participants to (a) be informed of the availability and eligibility criteria of any COVID-19 vaccine made available under an EUA and (b) the option to offer all ongoing study participants who request unblinding an opportunity to schedule a study visit to know their original group assignment (placebo vs. mRNA-1273 vaccine). Part B, the Open-Label Observational Phase of the study, also provides the opportunity for eligible study participants who previously received placebo to actively request to receive 2 doses of mRNA-1273 vaccine (as long as investigational vaccine is available).

#### 4.3. Choice of Dose and Control Product

The dose selected for this study (100 µg of mRNA-1273) is based on assessment of available safety and immunogenicity data from Phase 1 studies of mRNA-1647 (NCT03382405), the DMID study 20-0003 entitled "Phase I, Open-Label, Dose-Ranging Study of the Safety and Immunogenicity of 2019-nCoV Vaccine (mRNA-1273) in Healthy Adults" (NCT04283461) and the mRNA-1273-P201 study (Section 2.2.2). The schedule (2 doses administered 28 days apart) is being used in these studies.

There is no licensed SARS-CoV-2 vaccine currently available to serve as a reference control; accordingly, 0.9% sodium chloride (normal saline) injection (USP) will be used as a placebo control.

#### 4.4. End of Study Definition

Participants are considered to have completed the study if they complete the final visit at Day 759 (Month 25), 24 months following their receipt of the original second dose of IP (where original refers to the IP received following randomization).

The study duration will be approximately 26 months for each participant. This includes a screening period of up to 1 month and a study period of 25 months that includes the first dose of IP on Day 1 and the second dose on Day 29. The participant's final scheduled visit will be on Day 759 (Month 25), 24 months after the second dose of IP on Day 29 (Month 1).

The end of study will be the final participant's final scheduled visit at Day 759 (Month 25).

### 5. STUDY POPULATION

The study population, adults at risk of SARS-CoV-2 infection who have no known history of SARS-CoV-2 infection, is a subset of the planned target population. Additionally, potential study participants at increased risk of complications from COVID-19 will be included since it is hypothesized that these participants might derive the greatest benefit from a vaccine. Participants ≥ 65 years of age will be eligible for enrollment with or without underlying medical conditions further increasing their risk of severe COVID-19.

Given the disproportionate disease burden of COVID-19 in racial and ethnic minorities, the study will also aim to enroll a representative sample of participants from these minority population and adjust site selection and enrollment accordingly, per FDA Draft Guidance "Enhancing the Diversity of Clinical Trial Populations - Eligibility Criteria, Enrollment Practices, and Trial Designs" (June, 2019). Study sites may be selected on the basis of SARS-COV-2 infection risk of the local population as well. Approximately 30,000 participants will be enrolled.

There will be no prospective approval of protocol deviations to recruitment and enrollment criteria (also known as protocol waivers or exemptions).

#### 5.1. Inclusion and Exclusion Criteria (Part A: Blinded Phase and Part B: Open-Label Phase)

##### 5.1.1. Inclusion Criteria

Participants are eligible to be included in the study only if all the following criteria apply:

1. Adults, ≥ 18 years of age at time of consent, who are at high risk of SARS-CoV-2 infection, defined as adults whose locations or circumstances put them at appreciable risk of exposure to SARS-CoV-2 and COVID-19.
2. Understands and agrees to comply with the study procedures and provides written informed consent.
3. Able to comply with study procedures based on the assessment of the investigator.
4. Female participants of nonchildbearing potential may be enrolled in the study. Nonchildbearing potential is defined as surgically sterile (history of bilateral tubal ligation, bilateral oophorectomy, hysterectomy) or postmenopausal (defined as amenorrhea for ≥ 12 consecutive months prior to Screening without an alternative medical cause). A follicle-stimulating hormone (FSH) level may be measured at the discretion of the investigator to confirm postmenopausal status (see additional information in Appendix 11.3).

5. Female participants of childbearing potential may be enrolled in the study if the participant fulfills all the following criteria:

   - Has a negative pregnancy test at Screening and on the day of the first dose (Day 1).
   - Has practiced adequate contraception or has abstained from all activities that could result in pregnancy for at least 28 days prior to the first dose (Day 1).
   - Has agreed to continue adequate contraception through 3 months following the second dose (Day 29).
   - Is not currently breastfeeding.

   Adequate female contraception is defined as consistent and correct use of a Food and Drug Administration (FDA) approved contraceptive method in accordance with the product label. For example:

   - Barrier method (such as condoms, diaphragm, or cervical cap) used in conjunction with spermicide
   - Intrauterine device
   - Prescription hormonal contraceptive taken or administered via oral (pill), transdermal (patch), subdermal, or IM route
   - Sterilization of a female participant's monogamous male partner prior to entry into the study

   Note: periodic abstinence (eg, calendar, ovulation, symptothermal, post-ovulation methods) and withdrawal are not acceptable methods of contraception.

6. *<Inclusion criterion regarding male contraception has been removed by Amendment 2>*
7. Healthy adults or adults with pre-existing medical conditions who are in stable condition. A stable medical condition is defined as disease not requiring significant change in therapy or hospitalization for worsening disease during the 3 months before enrollment.

##### 5.1.2. Exclusion Criteria

Participants are excluded from the study if any of the following criteria apply:

1. Is acutely ill or febrile 72 hours prior to or at Screening. Fever is defined as a body temperature ≥ 38.0°C/100.4°F. Participants meeting this criterion may be rescheduled within the relevant window periods. Afebrile participants with minor illnesses can be enrolled at the discretion of the investigator.
2. Is pregnant or breastfeeding.

3. (Part A Only) Known history of SARS-CoV-2 infection.
4. Prior administration of an investigational coronavirus (SARS-CoV, MERS-CoV) vaccine or current/planned simultaneous participation in another interventional study to prevent or treat COVID-19.
5. (Part A Only) Demonstrated inability to comply with the study procedures.
6. (Part A Only) An immediate family member or household member of this study's personnel.
7. Known or suspected allergy or history of anaphylaxis, urticaria, or other significant AR to the vaccine or its excipients.
8. Bleeding disorder considered a contraindication to intramuscular injection or phlebotomy.
9. Has received or plans to receive a non-study vaccine within 28 days prior to or after any dose of IP (except for seasonal influenza vaccine which is not permitted within 14 days before or after any dose of IP, see Section 6.4.3).
10. Has participated in an interventional clinical study within 28 days prior to the day of enrollment.
11. Immunosuppressive or immunodeficient state, asplenia, recurrent severe infections (HIV-positive participants with CD4 count ≥350 cells/mm$^3$ and an undetectable HIV viral load within the past year [low level variations from 50-500 viral copies which do not lead to changes in antiretroviral therapy [ART] are permitted]).
12. Has received systemic immunosuppressants or immune-modifying drugs for >14 days in total within 6 months prior to Screening (for corticosteroids ≥ 20 mg/day of prednisone equivalent).
13. Has received systemic immunoglobulins or blood products within 3 months prior to the day of screening.
14. Has donated ≥ 450 mL of blood products within 28 days prior to Screening.

#### 5.2. Participant Restrictions

Participants must not eat or drink anything hot or cold within 10 minutes before oral temperature is taken.

#### 5.3. Screen Failures (Part A: Blinded Phase Only)

Screen failures are defined as participants who consent to participate in the clinical study but are not subsequently randomly assigned to treatment. A minimum set of screen failure information is required to ensure transparent reporting of screen failures to meet the Consolidated Standards of Reporting Trials (CONSORT) publishing requirements and to respond to queries from regulatory authorities. Minimum information includes date of informed consent, demography, screen failure details, eligibility criteria, and information on any SAE that may have occurred from the time informed consent was obtained to the time of withdrawal.

Participants meeting the exclusion criterion #1, acutely ill or febrile prior to or at the Screening Visit (exclusion criterion #1, Section 5.1.2), may be rescheduled within the relevant window periods and will retain their initially assigned participant number.

### 6. STUDY TREATMENT

#### 6.1. Investigational Product

The term "investigational product" refers to both mRNA-1273 vaccine (blinded and open label) and placebo administered in this study.

The mRNA-1273 IP is an LNP dispersion of an mRNA encoding the prefusion stabilized S protein of SARS-CoV-2 formulated in LNPs composed of 4 lipids (1 proprietary and 3 commercially available): the proprietary ionizable lipid SM-102; cholesterol; 1,2-distearoyl-sn-glycero-3-phosphocholine (DSPC); and 1-monomethoxypolyethyleneglycol-2,3-dimyristylglycerol with polyethylene glycol of average molecular weight 2000 (PEG2000-DMG). The mRNA-1273 vaccine is provided as a sterile liquid for injection and is a white to off white dispersion in appearance, at a concentration of 0.2 mg/mL in 20 mM Tris buffer containing 87 mg/mL sucrose and 10.7 mM sodium acetate at pH 7.5.

The placebo is 0.9% sodium chloride (normal saline) injection, which meets the criteria of the United States Pharmacopeia (USP).

#### 6.2. Dosing and Management of Investigational Product

##### 6.2.1. Method of Randomly Assigning Participants to Treatment Groups (Part A, Blinded Phase Only)

Approximately 30,000 participants will be randomly assigned in 1:1 ratio to receive either mRNA-1273 100 µg or placebo. The randomization will be in a blinded manner using a centralized Interactive Response Technology (IRT), in accordance with pre-generated randomization schedules. Only the unblinded personnel (Section 6.2.8.1) will have controlled access to which arm the participant is randomly assigned.

Dose group assignment in is summarized in Table 2.

**Table 2: Summary of Treatment Groups**

| Treatment Groups | Investigational Product | Age (years) | Estimated Total Participants |
|---|---|---|---|
| mRNA-1273 | mRNA-1273 100 µg | ≥ 18 | 15,000 |
| Placebo | Placebo | ≥ 18 | 15,000 |
| **Total** | | | **30,000** |

###### 6.2.1.1. Stratification

Randomization in Part A, Blinded Phase of the study will be stratified based on age and, if they are < 65 years of age, based on the presence or absence of risk factors for severe illness from COVID-19 based on CDC recommendation as of Mar 2020 (CDC 2020b). There will be 3 strata for randomization: ≥ 65 years, < 65 years and categorized to be at increased risk ("at risk") for the complications of COVID-19, and < 65 years "not at risk." Risk will be defined based on the study participants' relevant past and current medical history. At least 25% of enrolled participants, up to 50%, will be either ≥ 65 years of age or < 65 years of age and at risk at Screening.

Participants who are < 65 years old will be categorized as at risk for severe COVID-19 illness if they have at least 1 of the following risk factors at Screening:

- Chronic lung disease (eg, emphysema and chronic bronchitis), idiopathic pulmonary fibrosis and cystic fibrosis) or moderate to severe asthma
- Significant cardiac disease (eg, heart failure, coronary artery disease, congenital heart disease, cardiomyopathies, and pulmonary hypertension)
- Severe obesity (body mass index ≥ 40 $kg/m^2$)
- Diabetes (Type 1, Type 2 or gestational)
- Liver disease
- HIV infection

##### 6.2.2. Administration of Investigational Product

Investigational product will be administered as an IM injection into the deltoid muscle on a 2-dose injection schedule on Day 1 and Day 29. Each injection will have a volume of 0.5 mL and contain mRNA-1273 100 µg or saline placebo. Preferably, vaccine should be administered into the nondominant arm. The second dose of IP should be administered in the same arm as the first dose.

The IP will be prepared for injection as a single 0.5 mL dose for each participant based on the randomization assignment, as detailed in the Pharmacy Manual. Unblinded personnel who will not participate in any other aspect of the study during Part A, the Blinded Phase, will perform IP accountability, dose preparation, and IP administration. The investigator will designate unblinded medically qualified personnel (not involved in assessments of study endpoints) to administer the IP according to the procedures stipulated in this study protocol (Part A, the Blinded Phase), and Pharmacy Manual. Study-specific training will be provided. Study site personnel who were blinded during the Blinded Phase, will be unblinded at the participant level at the Participant Decision clinic visit.

At each visit when IP is administered, participants will be monitored for a minimum of 30 minutes after administration. Assessments will include vital sign measurements and monitoring for local or systemic reactions (Table 16, Table 21, and Table 22).

Eligibility for a subsequent dose of IP is determined by following the criteria outlined in Section 7.

The study site will be appropriately staffed with individuals with basic CPR training/certification. Either on site resuscitation equipment and personnel or appropriate protocols for the rapid transport of participant to a resuscitation area/facility are required.

###### 6.2.3. Delivery and Receipt

The Sponsor or designee is responsible for the following:

- Supplying the IP
- Confirming the appropriate labeling of mRNA-1273 IP, so that it complies with the legal requirements of the US

The investigator is responsible for acknowledging the receipt of the IP by a designated staff member at the site, including the following:

- Confirming that the IP was received in good condition
- Confirmation that the temperature during shipment from the Sponsor to the investigator's designated storage location was appropriate
- Confirming whether the Sponsor has authorized the IP for use
- Ensuring the appropriate dose level of mRNA-1273 is properly prepared using aseptic technique

Further description of the IP and instructions for the receipt, storage, preparation, administration, accountability, and destruction of the IP are described in the mRNA-1273-P201 Pharmacy Manual.

###### 6.2.4. Packaging and Labeling

The Sponsor will provide the investigator and study site with adequate quantities of mRNA-1273. The sterile vaccine product is packaged in a 10R glass vial with a 6-dose, 5.0-mL or 10-dose, 6.3-mL fill volume. mRNA-1273 vaccine will have all required labeling per regulations and will be supplied to the pharmacy in an unblinded manner. Each vial will be individually labeled for future participant identification purposes.

mRNA-1273 will be packaged and labeled in accordance with the standard operating procedures of the Sponsor or of its designee, Code of Federal Regulations Title 21 (CFR), Good Manufacturing Practice (GMP) guidelines, International Council for Harmonisation (ICH) GCP guidelines, guidelines for Quality System Regulations, and applicable regulations.

The Sponsor or Sponsor's designee will supply the 0.9% sodium chloride injection for use as both a placebo and a diluent to mRNA-1273. The 0.9% sodium chloride bears a commercial label and does not contain study-specific identification.

##### 6.2.5. Storage

The 6-dose mRNA-1273 vaccine must be stored at 2°C to 8°C in a secure area with limited access (unblinded personnel only) and protected from moisture and light until it is prepared for administration (Section 6.2.2). The 10-dose mRNA-1273 vaccine must be stored at -20°C in a secure area with limited access (unblinded personnel only) and protected from moisture and light until it is prepared for administration. The 10-dose mRNA-1273 vaccine can be stored at 2°C to 8°C for up to 30 days, but site personnel must account for this manually as it will not be managed in the IRT system. The freezer or refrigerator should have automated temperature recording and a 24-hour alert system in place that allows for rapid response in case of freezer or refrigerator malfunction. There must be an available backup freezer or refrigerator. The freezers or refrigerators must be connected to a backup generator. In addition, vaccine accountability study staff (eg, the unblinded personnel) are required to keep a temperature log to establish a record of compliance with these storage conditions. The site is responsible for reporting any mRNA-1273 vaccine that was not temperature controlled during shipment or during storage to the unblinded site monitor. Such mRNA-1273 will be retained for inspection by the unblinded monitor and disposed of according to approved methods.

The 0.9% sodium chloride injection (USP) should be stored at 20ºC to 25ºC (68ºF to 77ºF) in a restricted access area.

##### 6.2.6. Investigational Product Accountability

It is the investigator's responsibility that the unblinded personnel maintain accurate records in an IP accountability log of receipt of all IP, inventory at the site, dispensing of mRNA-1273 and placebo, IP injections, and return to the Sponsor or alternative disposition of used/unused products.

An unblinded site monitor will review the inventory and accountability log during site visits and at the completion of the study. Additional details are found in the mRNA-1273-P301 Pharmacy Manual.

##### 6.2.7. Handling and Disposal

An unblinded site monitor will reconcile the IP during the conduct and at the end of the study for compliance. Once fully reconciled at the site at the end of the study, the IP can be destroyed at the investigational site or at a Sponsor-selected third party, as appropriate.

Investigational product may be destroyed at the study site only if permitted by local regulations and authorized by the Sponsor. A Certificate of Destruction must be completed and sent to the Sponsor or designee.

##### 6.2.8. Unblinding

###### 6.2.8.1. Planned Unblinding

See Section 4.1.2 regarding the Participant Decision clinic visit.

- Part A, the Blinded Phase of this study is observer-blind. The investigator, study staff, study participants, site monitors, and Sponsor personnel (or its designees) will be blinded to the IP administered until study end, with the following exceptions:
- Unblinded personnel (of limited number) will be assigned to vaccine accountability procedures and will prepare IP for all participants. These personnel will have no study functions other than study vaccine management, documentation, accountability, preparation, and administration. They will not be involved in participant evaluations and will not reveal the identity of IP to either the participant or the blinded study site personnel involved in the conduct of the study unless this information is necessary in the case of an emergency.
- Unblinded medically qualified study site personnel will administer the IP. They will not be involved in assessments of any study endpoints.
- The dosing assignment will be concealed by having the unblinded personnel prepare the IP in a secure location that is not accessible or visible to other study staff. An opaque sleeve over the syringe used for injection will maintain the blind at the time of injection, as the doses containing mRNA-1273 will look different than placebo. Only delegated unblinded site staff will conduct the injection procedure. Once the injection is completed, only the blinded study staff will perform further assessments and interact with the participants. Access to the randomization code will be strictly controlled at the pharmacy.
- Unblinded site monitors, not involved in other aspects of monitoring, will be assigned as the IP accountability monitors. They will have responsibilities to ensure that sites are following all proper IP accountability, preparation, and administration procedures.
- An unblinded statistical and programming team will perform the pre-planned IAs (Section 9.6).
- An independent DSMB will review the interim data to safeguard the interests of clinical study participants and to help ensure the integrity of the study. The DSMB will review unblinded statistical outputs and IA results, provided by the independent unblinded statistician, and make recommendations to the Sponsor (Section 8.4.2).

- At the initiation of Part B, the Open-Label Observational Phase of this study (Section 4.1.3), study site personnel who were blinded during the Blinded Phase will be unblinded at the participant level at the Participant Decision clinic visit.

If prespecified criteria for early efficacy are met by an IA or if the primary efficacy analysis is completed based on accrual of prespecified COVID-19 cases, pre-identified Sponsor and CRO team members responsible for the analysis and reporting will be unblinded to treatment assignments in order to prepare a final study report. In order to maintain an observer-blind design, investigators, site staff, participants, and Sponsor and CRO staff with oversight of study conduct will remain blinded to treatment allocation for the study duration. All study participants will be followed for efficacy and safety endpoints through the remainder of planned study period and results will be summarized in an end of study report (Sections 4.1 and 9.1).

###### 6.2.8.2. Unplanned Unblinding

A participant or participants may be unblinded in the event of an SAE or other severe event, or if there is a medical emergency requiring the identity of the product to be known to properly treat a participant. If a participant becomes seriously ill or pregnant during the study, the study investigator may request that the blind will be broken if knowledge of the administered vaccine will affect that participant's dosing options. In this situation or in the event of a medical emergency requiring identification of the IP administered to an individual participant, the investigator will make every attempt to contact the Sponsor medical lead to explain the need for opening the code within 24 hours of opening the code. The investigator will be responsible for documenting the time, date, reason for the code break, and the names of the personnel involved.

In addition to the situations described above, where the blind may be broken, the data will also be unblinded to a statistical team at specified time points for IAs as outlined in Section 9.6.

In December 2020, COVID-19 vaccines have started to become available under EUA as an alternative option to some participants based on evolving recommended populations by the CDC and local supply chain distribution. While all participants should be encouraged to stay blinded in the study for as long as possible, their participation should not otherwise deny them the opportunity to receive a COVID-19 vaccine under EUA. Clinical investigators can exercise discretion as to whether individual participants should be unblinded upon request to allow them to make an informed decision regarding receipt of a COVID-19 vaccine outside of this study. Investigator judgment should consider a participant's risk status under CDC recommendations, any current local public health guidance, and their access to imminently receive a COVID-19 vaccine under an EUA.

If a decision is made to unblind a participant to support an informed decision to receive a COVID-19 vaccine under an EUA outside of this study, investigators are asked to take the following steps:

- Obtain a final assessment of safety from the participant to collect and resolve any outstanding safety experience. Verbal interview is sufficient.
- Unblind the participant in the IRT and inform the participant of their treatment assignment.
- Inform the Sponsor Medical Lead within 24 hours to confirm rationale for unblinding.

Participants who are confirmed to have received placebo upon unblinding and intend to receive a COVID-19 vaccine under an EUA outside of this study will be withdrawn from the study at the point of unblinding (Section 7.3.1).

Note: Participants who requested unblinding because of a stated imminent access to a COVID-19 vaccine under an EUA outside of this study, were withdrawn from study, and then were unable to in fact receive a COVID-19 vaccine, will be permitted to re-enter this study upon re-consent to receive the mRNA-1273 vaccine, once they are eligible (as long as investigational vaccine is available).

#### 6.3. Study Treatment Compliance

All doses of IP will be administered at the study site under direct observation of unblinded medically qualified study personnel and appropriately recorded (date and time) in the eCRF. Unblinded personnel will confirm that the participant has received the entire dose of vaccine. If a participant does not receive vaccine or does not receive all of the planned doses, the reason for the missed dose will be recorded. Data will be reconciled with site accountability records to assess compliance.

Participants who miss the second dose of IP due to noncompliance with the visit schedule and not due to a safety pause will still be required to follow the original visit and testing schedule as described in the protocol. Unless consent is withdrawn, a participant who withdraws or is withheld from receiving the second dose of study vaccine will remain in the study and complete all efficacy, safety and immunogenicity assessments required through the participant's scheduled end of study.

The study site is responsible for ensuring that participants comply with the study windows allowed. If a participant misses a visit, every effort should be made to contact the participant and complete a visit within the defined visit window (SoE Tables, Section 11.1). If a participant does not complete a visit within the time window, that visit will be classified as a missed visit (with the exception of Dose 2 visits in Part A and Part B, as applicable) and the participant will continue with subsequent scheduled study visits. All safety requirements of the missed visit will be captured and included in the subsequent visit (eg, clinical laboratory testing, eDiary review for reactogenicity, immunologic testing, as applicable).

#### 6.4. Prior and Concomitant Therapy

##### 6.4.1. Prior Medications and Therapies

Information about prior medications (including any prescription or over-the-counter medications, vaccines, or blood products) taken by the participant within the 28 days before providing informed consent (or as designated in the inclusion/exclusion requirements) will be recorded in the participant's eCRF.

##### 6.4.2. Concomitant Medications and Therapies

Study site staff must question the participant regarding any medications taken and vaccinations received by the participant and record the following information in the eCRF:

- All non-study vaccinations administered within the period starting 28 days before the first dose of IP.
- Seasonal influenza vaccine administered for the current influenza season (typically October through April in the Northern Hemisphere).
- Part A only: All concomitant medications and non-study vaccinations taken through 28 days after each dose of IP. Antipyretics and analgesics taken prophylactically (ie, taken in the absence of any symptoms in anticipation of an injection reaction) will be recorded as such.
- Any concomitant medications used to prevent or treat COVID-19.
- Any concomitant medications relevant to or for the treatment of an SAE or a MAAE.
- Part A only: Participant will be asked in the eDiary if they have taken any antipyretic or analgesic to treat or prevent fever or pain within 7 days after each IP injection, including the day of dosing. Reported antipyretic or analgesic medications should be recorded in the source document by the site staff during the post-injection study visits or via other participant interactions (eg, phone calls).

##### 6.4.3. Concomitant Medications and Vaccines that May Lead to the Elimination of a Participant from Per-protocol Analyses

The use of the following concomitant medications and/or vaccines will not require withdrawal of the participant from the study but may determine a participant's eligibility to receive a second dose or evaluability in the per-protocol (PP) analysis (Analysis Sets are described in Section 9.4):

- Any investigational or nonregistered product (drug or vaccine) other than the study vaccine used during the study period.
- A non-study vaccine administered during the period from 28 days before through 28 days after each dose of IP or any seasonal influenza vaccine that was administered within 14 days before or after any dose of IP.
- Immunoglobulins and/or any blood products administered during the study period (except for treatment of COVID-19).
- Medications that suppress the immune system (except for treatment of COVID-19).

If a participant takes a prohibited drug therapy, the investigator and the contract research organization's (CRO's) medical monitor will make a joint decision about continuing or withholding further dosing from the participant based on the time the medication was administered, the drug's pharmacology and pharmacokinetics, and whether use of the medication will compromise the participant's safety or the interpretation of data. It is the investigator's responsibility to ensure that details regarding the concomitant medications are adequately recorded in the eCRF.

All medication and interventions necessary for the appropriate care for the study participant, particularly to treat COVID-19, should be administered and appropriately documented along with the AE.

### 7. DELAYING OR DISCONTINUING STUDY TREATMENT AND PARTICIPANT WITHDRAWAL FROM THE STUDY

#### 7.1. Criteria for Delay of Study Treatment

Body temperature must be measured at the Day 1 and Day 29 visits prior to any study treatment administration. The following events constitute criteria for delay of study treatment, and if either of these events occur at the time scheduled for dosing, the participant may be injected at a later date within the time window specified in Table 16, or the participant may be discontinued from dosing at the discretion of the investigator (Section 7.2):

- Acute moderate or severe infection with or without fever at the time of dosing
- Fever, defined as body temperature ≥ 38.0°C (100.4°F) at the time of dosing

Participants with a minor illness without fever, as assessed by the investigator, can be administered IP. Participants with a fever of 38.0°C (100.4°F) or higher will be contacted within the time window acceptable for participation and reevaluated for eligibility. If the investigator determines that the participant's health on the day of administration temporarily precludes dosing with IP, the visit should be rescheduled within the allowed interval for that visit.

#### 7.2. Discontinuation of Study Treatment

Every reasonable attempt will be made to follow up with participants for safety throughout the entire study period, even if further dosing is discontinued or the participant misses one or more visits. Unless consent is withdrawn, a participant who withdraws or is withheld from receiving the second dose of study vaccine will remain in the study and complete all scheduled visits and assessments (Section 11.1).

The investigator, in consultation with the Sponsor's medical monitor, may withhold a participant from further dosing if the participant experiences any of the following:

- Becomes pregnant (Section 8.3.5)
- Develops, during the course of the study, symptoms or conditions listed in the exclusion criteria
- Experiences an AE (other than reactogenicity) after dosing that is considered by the investigator to be related to IP (Section 8.3.4) and is of Grade 3 (severe) or greater intensity (Section 8.3.8)
- Experiences an AE or SAE that, in the judgment of the investigator, requires study IP withdrawal due to its nature, severity, or required treatment, regardless of the causal relationship to vaccine
- Experiences a clinically abnormal vital sign measurement or finding on physical examination, or general condition that, in the judgment of the investigator, requires IP withdrawal

The reason(s) for discontinuation from further dosing will be recorded in the eCRF.

Prior to receiving a second dose of IP, participants will be reassessed to ensure that they continue to meet eligibility requirements as outlined below.

The following events in a participant constitute absolute contraindications to any further dosing of the IP to that participant. If any of these events occur during the study, the participant must not receive additional doses of vaccine but will be encouraged to continue study participation for safety through 24 months following last dose.

- Part A only: Diagnosed COVID-19 by detection of SARS-CoV-2 in Day 1 NP swab or COVID-19 diagnosed prior to Day 29. If COVID-19 is suspected on or prior to Day 29, further administration of IP must be withheld until COVID-19 test results are available.
- Anaphylaxis or systemic hypersensitivity reaction following the administration of vaccine.
- Any SAE judged by investigator or Sponsor to be related to study vaccine.
- Any clinically significant medical condition that, in the opinion of the investigator, poses an additional risk to the participant if he/she continues to participate in the study.

The following events constitute contraindications to administration of study vaccine at certain points in time, and if any of these events occur at the time scheduled for dosing, the participant may be injected at a later date, within the time window specified in the SoE (Table 16), or the participant may be withdrawn from dosing at the discretion of the investigator (Section 7.3):

- Acute moderate or severe infection with or without fever at the time of dosing
- Fever, defined as body temperature ≥ 38.0°C (100.4°F) at the time of dosing
- Part B only: Diagnosed COVID-19 by detection of SARS-CoV-2 by RT-PCR and accompanying symptoms. Participant must be asymptomatic at the time of dosing although may still be RT-PCR positive, with the exception of loss of sense of smell or taste and fatigue, if by the investigator's judgement these are deemed to be mild chronic sequelae.

Participants with a minor illness without fever, as assessed by the investigator, can be administered IP. Participants with a fever of 38.0°C (100.4°F) or higher will be contacted within the time window acceptable for participation and reevaluated for eligibility.

#### 7.3. Participant Withdrawal from the Study

##### 7.3.1. Participant Withdrawal

Participants who withdraw from the study will not be replaced. A "withdrawal" from the study refers to a situation wherein a participant does not return for the final visit planned in the protocol. The statistical management of participant withdrawals is discussed in Section 9.

Participants can withdraw consent and withdraw from the study at any time, for any reason, without prejudice to further treatment the participant may need to receive. The investigator will request that the study participant complete all study procedures pending at the time of withdrawal.

If participant desires to withdraw from the study because of an AE, the investigator will try to obtain agreement to follow up with the participant until the event is considered resolved or stable and will then complete the end of study eCRF.

Information related to the withdrawal will be documented in the eCRF. The investigator will document whether the decision to withdraw a participant from the study was made by the participant or by the investigator, as well as which of the following possible reasons was responsible for withdrawal:

- AE (specify)
- SAE (specify)
- Death
- Lost to follow-up
- Physician decision (specify)
- Pregnancy
- Protocol deviation
- Study terminated by Sponsor
- Withdrawal of consent by participant (specify); this includes participants who, at the Participant Decision clinic visit, withdraw consent from continuing in the study.
- Other (specify)

Participants who are withdrawn from the study because of AEs (including SAEs) must be clearly distinguished from participants who are withdrawn for other reasons. Investigators will follow up with participants who are withdrawn from the study as result of an SAE or AE until resolution of the event.

Participants who are confirmed to have received placebo upon unblinding and intend to receive a COVID-19 vaccine under an EUA outside of this study will be withdrawn from the study.

A participant withdrawing from the study may request destruction of any samples taken and not tested, and the investigator must document this in the site study records.

If the participant withdraws consent for disclosure of future information, the Sponsor may retain and continue to use any data collected before such a withdrawal of consent (see Section 11.2.10).

The Sponsor will continue to retain and use all research results that have already been collected for the study evaluation, unless the participant has requested destruction of these samples. All biological samples that have already been collected may be retained and analyzed at a later date (or as permitted by local regulations).

#### 7.4. Lost to Follow-up

Participants will be considered lost to follow-up (LTFU) if they repeatedly fail to return for scheduled visits without stating an intention to withdraw consent and they cannot be contacted by the study site. The following actions must be taken if a participant fails to return to the clinic for a required study visit:

- The site must attempt to contact the participant and reschedule the missed visit as soon as possible, counsel the participant on the importance of maintaining the assigned visit schedule and ascertain whether the participant wishes to and/or should continue in the study.
- Before a participant is deemed lost to follow-up, the investigator or designee must make every effort to regain contact with the participant (where possible, 3 telephone calls and, if necessary, a certified letter to the participant's last known mailing address or local equivalent methods). These contact attempts (eg, dates of telephone calls and registered letters) should be documented in the participant's medical record. A participant should not be considered LTFU until these efforts have been made.
- Should the participant continue to be unreachable, he/she will be considered to have withdrawn from the study.

### 8. STUDY ASSESSMENTS AND PROCEDURES

Before performing any study procedures, all potential participants will sign an ICF (as detailed in Section 11.2.6). Participants will undergo study procedures at the time points specified in the SoEs (Section 11.1).

A participant also can be seen for an unscheduled visit at any time during the study. An unscheduled visit may be prompted by reactogenicity issues, Illness Visit criteria for COVID-19, or new or ongoing AEs. The site also has the discretion to make reminder telephone calls or send text messages to inform the participant about visits, review eDiary requirements, or follow-up on ongoing or outstanding issues.

In accordance with "FDA Guidance on Conduct of Clinical Trials of Medical Products during the COVID-19 Pandemic" (DHHS 2020), investigators may convert study site visits to telemedicine visits with the approval of the Sponsor. Such action should be taken to protect the safety and well-being of study participants and study site staff or to comply with state or municipal mandates.

General considerations for study assessments and procedures include the following:

- Protocol waivers or exemptions are not allowed. The study procedures and their timing must be followed as presented in Section 11.1. Adherence to the study design requirements is essential and required for study conduct.
- Immediate safety concerns should be discussed with the Sponsor immediately upon occurrence or awareness to determine if the participant should continue study treatment or participation in the study.
- All screening evaluations must be completed and reviewed to confirm that potential participants meet all eligibility criteria. The investigator will maintain a screening log to record details of all participants screened and to confirm eligibility or record reasons for screening failure, as applicable.
- Procedures conducted as part of the participant's routine clinical management (eg, blood count) and obtained before signing of the ICF may be utilized for screening or baseline purposes provided the procedures met the protocol-specified criteria and were performed within the time frame defined in the SoE.

#### 8.1. Efficacy and Immunogenicity Assessments and Procedures

##### 8.1.1. Efficacy Assessments Related to COVID-19 and SARS-CoV-2 Infection

Each study participant will have an NP swab sample collected for SARS-CoV-2 testing by RT-PCR on Day 1 and Day 29, prior to receiving a dose of the IP as specified in the SoE (Section 11.1).

**COVID-19:**

To be considered as a case of COVID-19 for the evaluation of the Primary Efficacy Endpoint, the following case definition must be met:

- The participant must have experienced at least TWO of the following systemic symptoms: Fever (≥ 38ºC), chills, myalgia, headache, sore throat, new olfactory and taste disorder(s), OR
- The participant must have experienced at least ONE of the following respiratory signs/symptoms: cough, shortness of breath or difficulty breathing, OR clinical or radiographical evidence of pneumonia; AND
- The participant must have at least one NP swab, nasal swab, or saliva sample (or respiratory sample, if hospitalized) positive for SARS-CoV-2 by RT-PCR.

**Severe COVID-19:**

To be considered severe COVID-19, the following criteria must be met:

- Confirmed COVID-19 as per the Primary Efficacy Endpoint case definition, plus any of the following:
  - Clinical signs indicative of severe systemic illness, Respiratory Rate ≥ 30 per minute, Heart Rate ≥ 125 beats per minute, $SpO_2$ ≤ 93% on room air at sea level or $PaO_2/FIO_2$ < 300 mm Hg, OR
  - Respiratory failure or Acute Respiratory Distress Syndrome (ARDS), (defined as needing high-flow oxygen, non-invasive or mechanical ventilation, or ECMO), evidence of shock (systolic blood pressure < 90 mmHg, diastolic BP < 60 mmHg or requiring vasopressors), OR
  - Significant acute renal, hepatic or neurologic dysfunction, OR
  - Admission to an intensive care unit or death.

The secondary case definition of COVID-19 is defined as the following systemic symptoms: fever (temperature ≥ 38ºC), or chills, cough, shortness of breath or difficulty breathing, fatigue, muscle aches, or body aches, headache, new loss of taste or smell, sore throat, nasal congestion or rhinorrhea, nausea, or vomiting or diarrhea AND a positive NP swab, nasal swab, or saliva sample (or respiratory sample, if hospitalized) for SARS-CoV-2 by RT-PCR.

Death attributed to COVID-19 is defined as any participant who dies during the study with a cause directly attributed to a complication of COVID-19.

**SARS-CoV-2 Infection:**

- SARS-CoV-2 infection is defined by seroconversion due to infection measured by bAb against SARS-CoV-2 nucleocapsid. Seroconversion is defined differently for participants seronegative at Baseline and seropositive at Baseline:
  - Participants seronegative at Baseline: bAb levels against SARS-CoV-2 nucleocapsid either from below the limit of detection (LOD) or lower limit of quantification (LLOQ) at Study Day 1 that increase to above or equal to LOD or LLOQ starting at Study Day 57 or later.
  - Participants seropositive at Baseline: bAb levels against SARS-CoV-2 nucleocapsid above the LOD or LLOQ at study Day 1 that increase by 4-fold or more in participants starting at Study Day 57 or later.

##### 8.1.2. Surveillance for COVID-19 Symptoms

Surveillance for COVID-19 symptoms will be conducted via weekly telephone calls in Part A, Blinded Phase and Part B, Open-Label Observational Phase of the study and/or eDiary prompts as specified in Section 8.2.2; Figure 4 starting after enrollment and throughout the study. If there is no response to an eDiary prompt for 2 days, the site staff will contact the study participant by phone.

According to the CDC as of 10 June 2020 (CDC 2020c), patients with COVID-19 have reported a wide range of symptoms ranging from mild symptoms to severe illness. Throughout the study, to surveil for COVID-19, the following prespecified symptoms that meet the criteria for suspicion of COVID-19 will be elicited weekly from the participant and the presence of any one of these symptoms lasting at least 48 hours (except for fever and/or respiratory symptoms) will result in the site arranging an Illness Visit to collect an NP swab within 72 hours:

- Fever (temperature ≥ 38ºC) or chills (of any duration, including ≤ 48 hours)
- Shortness of breath or difficulty breathing (of any duration, including ≤ 48 hours)
- Cough (of any duration, including ≤ 48 hours)
- Fatigue
- Muscle or body aches

- Headache
- New loss of taste or smell
- Sore throat
- Congestion or runny nose
- Nausea or vomiting
- Diarrhea

It is important to note that some of the symptoms of COVID-19 overlap with solicited systemic ARs that are expected after vaccination with mRNA-1273 (eg, myalgia, headache, fever, and chills). During the first 7 days after vaccination, when these solicited ARs are common, investigators should use their clinical judgement to decide if an NP swab should be collected (Part A, the Blinded Phase only). The collection of an NP swab prior to the Day 1 and Day 29 vaccination can help ensure that cases of COVID-19 are not overlooked. Any study participant reporting respiratory symptoms during the 7-day period after vaccination should be evaluated for COVID-19.

**Figure 4: Surveillance for COVID-19 Symptoms and the Corresponding Clinical Data Pathways**

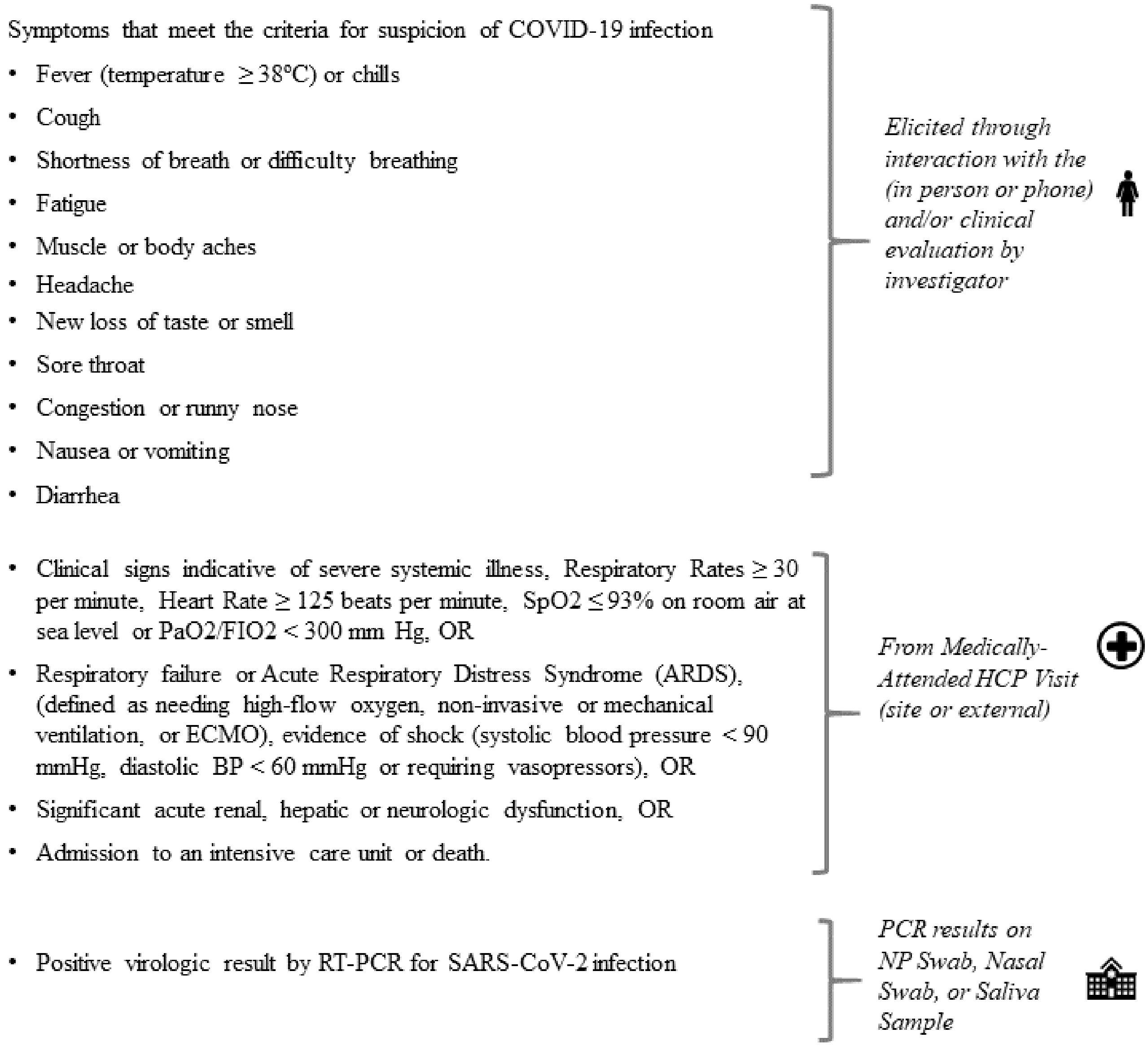

During the course of the study, participants with symptoms of COVID-19 will be asked to return within 72 hours or as soon as possible to the study site or medically qualified staff from the study site will conduct a home visit as soon as possible to collect an NP swab sample (for RT-PCR), collect a blood sample for immunologic analysis of SARS-CoV-2 infection, and to evaluate for COVID-19. Both study site visits and home visits are referred to as Illness Visits. The NP swab sample will also be tested for the presence of other respiratory infections.

In addition, the study site may collect an additional respiratory sample for SARS-CoV-2 testing to be able to render appropriate medical care for the study participant as determined by local standards of care. If neither a study site visit or home visit is possible, participants will be sent a saliva kit via courier or other Sponsor-approved method. The study site will arrange to retrieve a saliva (or nasal swab) sample by local courier or other Sponsor-approved method, and the sample will be tested by RT-PCR for SARS-CoV-2. If participants are confirmed to have SARS-CoV-2 infection, the investigator will notify the participant, and the participant's primary care physician, of the diagnosis. If the study participant does not have a primary care physician, the investigator will assist them to obtain one. The participant will also be instructed on infection prevention measures consistent with local public health guidance.

If scheduled, a study site Illness Visit may include assessments such as medical history, physical examination, blood sampling for clinical laboratory testing, and NP swab sampling for viral PCR (including multiplex PCR for respiratory viruses including SARS-CoV-2) to evaluate the severity of the clinical case. Radiologic imaging studies may be conducted. Blood samples will be collected for potential future immunologic assessment of SARS-CoV-2 infection.

Cases are defined as participants meeting clinical criteria based both on symptoms for COVID-19 and on RT-PCR detection of SARS-CoV-2 from samples collected within 72 hours of the study participant reporting symptoms meeting the definition of COVID-19. Participants who are hospitalized for COVID-19 without the opportunity for a clinic or home visit will also be considered cases, assuming that the symptomology criteria for COVID-19 are met and a respiratory sample is positive for SARS-CoV-2 by PCR at a Clinical Laboratory Improvement Amendments (CLIA)-certified or CLIA-certified waiver laboratory. Investigators are encouraged to try to obtain a respiratory sample during the course of hospitalization for submission to the study central laboratory, if feasible. The investigator should determine if the criteria for severe COVID-19 has been met.

Evidence of severe COVID-19 is defined as in Section 8.1.1.

All clinical findings will be recorded in the eCRF. All confirmed cases of COVID-19 will be captured as MAAEs, along with relevant concomitant medications and details about severity, seriousness, and outcome.

##### 8.1.3. Convalescent Period Starting with the Illness Visit

All study participants who experience COVID-19 symptoms and subsequently present for an Illness Visit (in-clinic or at home) will be given an instruction card listing symptoms and severity grading system, a thermometer, an oxygen saturation monitor, and saliva collection tubes. Participants will be trained on the use of the oxygen saturation monitor and how to take saliva specimens. The list of symptoms is presented in Section 8.1.2 and the severity scoring system is presented in Table 3.

**Table 3: Grading of COVID-19 Symptoms**

| Grading | All Symptoms | For Nausea/Vomiting ONLY | For Sense of Smell/Taste ONLY |
|---|---|---|---|
| None | No symptom | | |
| Mild | I had the symptom, but I could still do my normal activities | I was able to eat and drink normally | I had the symptom, but I retained some taste/smell |
| Moderate | The symptom really bothered me. It was hard to do my normal activities | It bothered me enough that I did not eat or drink normally | My taste/smell was significantly affected |
| Severe | The symptom was very bad. I was not able to do activities that I usually do | I could not eat or drink | I have no taste or smell |

The initial Illness Visit is considered Day 1 for the Convalescent Period. Starting on Day 2 of the Convalescent Period, the investigator (or medically qualified staff appropriately delegated by the investigator) will contact participants daily with telemedicine visits through Day 14 or until symptoms have resolved, whichever is later. Telemedicine visits may be conducted by videoconference or by audio only (telephone). During the telemedicine visit (preferably done in the evening), the participant will be asked to verbally report the severity of each symptom, their highest body temperature and lowest oxygen saturation for that day, and the investigator will determine if medical attention is required due to worsening of COVID-19 symptoms. The presence and severity of each symptom reported by the participant will be noted in the appropriate source document (Table 19).

During the telemedicine visits, participants will be reminded both to collect their own saliva (or nasal swab) sample on 3, 5, 7, 9, 14, and 21 days after the initial Illness Visit and to return the sample to the study site. Immediately upon receipt of a saliva (or nasal swab) sample, the study site will send it for testing to the study central virology laboratory.

During the telemedicine visits, if the participant has a positive result for SARS-CoV-2 from the Day 1 Illness Visit, the participant will continue the Convalescent Period. If the participant has a negative result for SARS-CoV-2 from the Day 1 Illness Visit, the participant will exit the Convalescent Period, including discontinuation of daily telemedicine visits and collection of saliva (or nasal swab) samples, and will return to their respective study schedule (Table 16, Table 17, and Table 18). Participants who are pending a central laboratory PCR may exit from the convalescent period based on a negative PCR result from a CLIA-certified or CLIA-certified waiver local laboratory, at the investigator's discretion.

All participants confirmed to be COVID-19 cases will be scheduled for a convalescent visit (study site or home visit) 28 days after the initial Illness Visit. At this visit, a saliva (or nasal swab) sample will be collected and a blood sample will be drawn for immunologic assessment of SARS-CoV-2 infection (Table 19).

If the participant is hospitalized, medically qualified site personnel will try to obtain medical records and SARS-CoV-2 diagnostic results and document if the criteria for COVID-19 or severe COVID-19 have been met. If the participant is later discharged from the hospital during the 28-day period following diagnosis of COVID-19, the study site personnel will arrange for a resumption of a schedule for telemedicine visits and sampling for the remainder of the Convalescent Period, followed by a return to their respective study schedule (Table 16, Table 17, and Table 18).

##### 8.1.4. Ancillary Supplies for Participant Use

Study sites will distribute Sponsor-provided oral thermometers and rulers for use by participants in assessing body temperature and injection site reactions for recording solicited ARs in eDiaries (Section 8.2.2). Based on availability, smartphone devices may be provided to those participants who do not have their own device to use for eDiary activities.

Participants will also receive the following Sponsor-provided supplies at Illness Visits where COVID-19 is suspected:

- An instruction card listing symptoms and severity grading system
- A pulse oximeter for measuring oxygen saturation
- Saliva collection tubes and instructions/means for returning saliva samples collected at home to the study site
- Additional oral thermometer, if required.

##### 8.1.5. Immunogenicity Assessments

Blood samples for immunogenicity assessments will be collected at the time points indicated in the SoEs (Section 11.1). On Day 1 and Day 29, as specified in Table 16, on OL-D1 and OL-D57 as specified in Table 21, and on OL-D1 and OL-D29 as specified in Table 22, blood samples for immunogenicity assessment will be collected before administration of study treatment. The following analytes will be measured:

- Serum bAb levels against SARS-CoV-2 as measured by ligand-binding assay specific to the SARS-CoV-2 S protein
- Serum bAb levels against SARS-CoV-2 as measured by ligand-binding assay specific to the SARS-CoV-2 nucleocapsid protein
- Serum nAb titer against SARS-CoV-2 as measured by pseudovirus and/or live virus neutralization assays

Serum will be tested using the ligand-binding assay specific to the SARS-CoV-2 nucleocapsid to determine the immunologic status of study participants at baseline and assess for seroconversion due to infection during the course of the study. Serum from a subset of participants will be tested in the other assays. The selection of the subset and timepoints to be tested will be described in the statistical analysis plan (SAP). Sample aliquots will be designed to ensure that backup samples are available and that adequate vial volumes may allow for further testing. The actual time and date of each sample collected will be recorded in the eCRF, and unique sample identification will be used to maintain the blind at the laboratory at all times and to allow for automated sample tracking and storage. Handling and preparation of the samples for analysis, as well as shipping and storage requirements, will be provided in a separate study manual.

The ligand-binding assay and measurement of nAb titers will be performed in laboratories designated by the Sponsor.

#### 8.2. Safety Assessments

Safety assessments will include monitoring and recording of the following for each participant:

- Solicited local and systemic ARs (Section 8.3.4) that occur during the 7 days following each injection (ie, the day of dosing and 6 subsequent days). Solicited ARs will be recorded daily using eDiaries (Section 8.2.2); (Part A, the Blinded Phase only).
- Unsolicited AEs observed or reported during the 28 days following each dose of IP (ie, the day of dosing and 27 subsequent days, (Part A, the Blinded Phase only). Unsolicited AEs are AEs that are not included in the protocol-defined solicited ARs (Section 8.3.4).
- AEs leading to discontinuation from dosing and/or study participation from Day 1 through Day 759 or withdrawal from the study.
- MAAEs from Day 1 through Day 759 or withdrawal from the study.
- SAEs from Day 1 through Day 759 or withdrawal from the study.
- Abnormal vital sign measurements.
- Physical examination findings.
- Pregnancy and accompanying outcomes.
- Concomitant medications and non-study vaccinations.

##### 8.2.1. Safety Phone Calls

A safety phone call is a telephone call made to the participant by medically qualified study staff. Medically qualified staff are those appropriately delegated individuals who are permitted to elicit verbal medical history from participants based on local regulations and local licensing requirements.

This call will follow a script, which will facilitate the collection of relevant safety information. The participant will be interviewed according to the script about occurrence of unsolicited AEs, MAAEs, SAEs, or AEs leading to study withdrawal and concomitant medications associated with those events, receipt of any non-study vaccinations, and pregnancy. Occurrence of AEs will only be collected by safety phone call during the Vaccination Phase (Table 16 and Table 21).

The timing of the safety phone calls and the relevant safety information collected is provided in the SoEs (Section 11.1).

All safety information described by the participant must be documented in source documents and not documented on the script used for the safety telephone contact. All AEs, MAAEs, SAEs, and AEs leading to study withdrawal must be recorded in the eCRF as specified in Section 8.3.6.

##### 8.2.2. Use of Electronic Diaries

In Part A, the Blinded Phase of the Study, at the time of consent, the participants must confirm they will be willing to complete an eDiary (for 7-day reactogenicity and to surveil weekly for COVID-19 symptoms) using either an application downloaded to their smartphone or using a device that is provided at the time of enrollment. This study will utilize the Medidata Patient Cloud Application as the eDiary for both collection of 7-day reactogenicity and weekly eDiary prompts to elicit an unscheduled Illness Visit if the participant is experiencing COVID-19 symptoms. This application allows for real-time data collection on a 21 CFR Part 11 compliant system directly from participants.

In Part A, the Blinded Phase of the Study, before enrollment on Day 1, the participant will be instructed to download the eDiary application on their personal smartphone or will be provided a Sponsor-provisioned device to record solicited ARs (Section 8.3.4) and also to be utilized for eDiary prompts through the COVID-19 surveillance period.

In Part A, the Blinded Phase of the Study, at each dosing visit, participants will record data into the eDiary starting approximately 30 minutes after dosing under supervision of the study site staff to ensure successful entry of assessments. The 30-minute assessment is an opportunity for site staff to train the participant. The site staff will perform any retraining as necessary. Study participants will continue to record data in an eDiary after they leave the study site, preferably in the evening and at the same time each day, on the day of dosing and for 6 days following dosing.

In Part A, the Blinded Phase of the Study, the following local ARs will be solicited by the eDiary: pain at injection site, erythema (redness) at injection site, swelling/induration (hardness) at injection site, and localized axillary swelling or tenderness ipsilateral to the injection arm.

The following systemic ARs will be solicited by the eDiary: headache, fatigue, myalgia (muscle aches all over the body), arthralgia (aching in several joints), nausea/vomiting, body temperature (potentially fever), and chills.

In Part A, the Blinded Phase of the Study, solicited local and systemic reactogenicity ARs, as defined in Section 8.3.4, will be collected on the day of each IP injection and during the 7 days after IP injection (ie, the day of dosing and 6 subsequent days). Any solicited AR that is ongoing beyond Day 7 will be reported in an eDiary until resolution. Adverse reactions recorded in diaries beyond Day 7 should be reviewed by study site staff either during the next scheduled phone call or at the next study site visit (Table 16).

If eDiary prompts result in identification of relevant safety events according to the study period-or symptoms of COVID-19, a follow-up safety call will be triggered.

In Part A, the Blinded Phase of the Study, at each dosing visit, participants will be instructed or reminded on thermometer usage to measure body temperature, ruler usage to measure injection site erythema and swelling/induration (hardness), and self-assessment for localized axillary swelling or tenderness on the same side as the injection arm.

In Part A, the Blinded Phase of the Study, daily oral body temperature measurement should be performed at approximately the same time each day using the thermometer provided by the study site.

Surveillance for signs and symptoms of COVID-19 will start at Part A Day 1 and continue through the entire Surveillance Phase of the study (Table 17 and Table 18). After participants complete Part A Vaccination Phase of the Study (Table 16 and Table 21), the weekly eDiary Safety Follow-up Prompts will be triggered on Day 61 (Table 17). The weekly eDiary prompts will utilize the same Medidata Patient Cloud Application. Each week (ie, every 7 days) the participant will receive a prompt on their smartphone device to regularly surveil for signs and symptoms of COVID-19 and any other changes in health status. The participant will be trained on how to complete the weekly eDiary prompts at the Part A Day 57 clinic visit and also reminded to call the site immediately if they experience any COVID-19 symptoms. The weekly eDiary prompt will inquire about the following:

- Changes in health since the last time completing the weekly eDiary prompt or the last contact with the study staff
- Any known exposure to someone with SARS-CoV-2 infection or COVID-19 since the last time completing the weekly eDiary prompt or the last contact with the study staff
- Capture of any COVID-19 symptoms currently being experienced or experienced since the last time completing the weekly check-in as defined in Section 8.1.2
- Any contact with a healthcare provider that may indicate a MAAE.

A positive response by the participant to these prompts will result in a notification to both the participant and study staff to arrange a call. A follow-up safety call will be performed to the participant to determine if an unscheduled Illness Visit for the participant should be arranged as defined in Section 8.1.2. The results of the safety call should be recorded in the appropriate source documentation.

If a participant does not respond to the weekly eDiary within a 2-day window around the scheduled timepoint, study staff will follow-up directly with the participant via phone call or text to confirm their health status and to remind the participant of the importance of maintaining weekly contact via the eDiary prompt.

In addition, there will be an eDiary prompt to solicit the collection of information regarding participant's history of facial injections or dermal fillers, for cosmetic or medical indications such as migraine headaches.

##### 8.2.3. Demographics/Medical History

Demographic information relating to the participant's sex, age, and race will be recorded at Screening on the appropriate eCRF page.

Additionally, information regarding participant occupational circumstances (eg, essential worker status) will be collected at Screening.

Medical history of each participant, including risk factors for severe COVID-19 as defined in Section 6.2.1.1, will be collected and recorded on the Medical History eCRF page. Significant findings that were present prior to the signature of the informed consent will also be included in the Medical History eCRF page.

Study participants will also be asked to report history of receipt of seasonal influenza vaccine during the current influenza season (typically October through April in the Northern Hemisphere) as a concomitant medication.

##### 8.2.4. Physical Examination

A full physical examination, including vital signs, height, and weight, will be performed at Screening and on Day 1, and symptom-directed physical examinations at other scheduled time points as indicated in the SoEs (Section 11.1). The full examination will include assessment of skin, head, ears, eyes, nose, throat, neck, thyroid, lungs, heart, cardiovascular, abdomen, lymph nodes, and musculoskeletal system/extremities. Symptom directed- physical examinations may be performed at other timepoints at the discretion of the investigator.

On each dosing day before injection, the arm receiving the injection should be examined and the associated lymph nodes should be evaluated.

Body mass index will be calculated at the Screening Visit (Day 0).

Any clinically significant finding identified during a study visit after the first dose should be reported as a MAAE.

Significant new findings that begin or worsen after informed consent must be recorded on the AE eCRF page.

##### 8.2.5. Vital Sign Measurements

Vital signs will be measured at the time points indicated in the SoEs (Section 11.1). The participant will be seated for at least 5 minutes before all measurements are taken. On Day 1 and Day 29, vital sign measurements will be collected once before IP injection and at least 30 minutes after IP injection (before participants are discharged from the study site). When applicable, vital sign measurements should be performed before blood collection.

Febrile participants at Day 1 and Day 29 visits (fever is defined as a body temperature ≥ 38.0°C/100.4°F) may be rescheduled within the relevant window periods. Criteria for delay of study treatment are provided in Section 7.1. Afebrile participants with minor illnesses may be injected at the discretion of the investigator.

If any of the vital sign measurements meet the toxicity grading criteria for clinical abnormalities of Grade 3 or greater, the abnormal value and Grade will be documented on the AE page of the eCRF (unless there is another known cause of the abnormality that would result in an AE classification). The investigator will continue to monitor the participant with additional assessments until the vital sign value has reached the reference range, returns to the vital sign value at baseline, is considered stable, or until the investigator determines that follow-up is no longer medically necessary.

##### 8.2.6. Blood Sampling Volumes

The maximum planned volumes of blood sampled per participant are approximately 50 mL for 1 day, 100 mL for 28 days, and 300 to 340 mL for the complete study (Table 4).

**Table 4: Maximum Planned Blood Sampling Volumes per Participant by Visit**

| Study Visit Day | Part A D1 | Part A D29 | Part A D57 | Part A D209 | Part A D394 | Part A D759 | Part A Total |
|---|---|---|---|---|---|---|---|
| Immunogenicity blood samples [a,b] | 50 mL | 50 mL | 50 mL | 50 mL | 50 mL | 50 mL | 300 mL |
| | **Open-Label Day 1** | **Open-Label Day 29[b]** | **Open-Label Day 57[b]** | NA | NA | NA | NA |
| | 20 mL | 20 mL | 20 mL | NA | NA | NA | 40 mL |

Abbreviations: D = Day; NA = not applicable; RT-PCR = reverse transcriptase polymerase chain reaction.

[a] If a Part A and Part B (Open-Label) visit overlap, only the Part B sample will be taken.

[b] For participants who received placebo during the Blinded Phase and receive mRNA-1273 during the Open-Label Observational Phase (Table 21). Note that immunogenicity blood samples are collected only on Open-Label Days 1 and 29 or Day 57 during the Open-Label Observational Phase.

[c] Additional blood samples (16 mL) will be taken at Illness Day 1 and Convalescent Visit Day 28 for those participants who report symptoms of COVID-19 and who have RT-PCR confirmed symptomatic COVID-19, respectively.

#### 8.3. Safety Definitions and Procedures

##### 8.3.1. Adverse Event

An AE is defined as any untoward medical occurrence associated with the use of a drug in humans, whether or not considered drug related.

A TEAE is defined as any event not present before exposure to IP or any event already present that worsens in intensity or frequency after exposure.

**Events Meeting the Adverse Event Definition**

- Exacerbation of a chronic or intermittent pre-existing condition including either an increase in frequency and/or intensity of the condition.
- New conditions detected or diagnosed after the first dose of IP even though they may have been present before the start of the study.

**Events NOT Meeting the Adverse Event Definition**

- Medical or surgical procedure (eg, endoscopy, appendectomy): the condition that leads to the procedure should be the AE.
- Situations in which an untoward medical occurrence did not occur (social and/or convenience admission to a hospital).

An AR is any AE for which there is a reasonable possibility that the IP caused the AE (Section 8.3.4). For the purposes of investigational new drug safety reporting, "reasonable possibility" means that there is evidence to suggest a causal relationship between the IP and the AE.

An unsolicited AE is any AE reported by the participant that is not specified as a solicited AR in the protocol; or is specified as a solicited AR in the protocol, but starts outside the protocol-defined period for reporting solicited ARs (ie, for the 7 days after each dose of IP, Part A, Blinded Phase only).

##### 8.3.2. Medically Attended Adverse Events

An MAAE is an AE that leads to an unscheduled visit (including a telemedicine visit) to a healthcare practitioner (HCP). This would include visits to a study site for unscheduled assessments (eg, rash assessment, abnormal laboratory follow-up, COVID-19 [Section 8.1.1]) and visits to HCPs external to the study site (eg, urgent care, primary care physician). Investigators will review unsolicited AEs for the occurrence of any MAAEs. All MAAEs must be fully reported on the MAAE page of the eCRF.

###### 8.3.2.1. Anaphylaxis

All suspected cases of anaphylaxis should be recorded as MAAEs and reported as an SAE, based on criteria for a medically important event, unless the event meets other serious criteria. As an SAE, the event should be reported to the Sponsor or designee immediately and in all circumstances within 24 hours as per Section 8.3.11 (Reporting SAEs). The investigator will submit any updated anaphylaxis case data to the Sponsor within 24 hours of it being available. For reporting purposes, a participant who displays signs/symptoms consistent with anaphylaxis as shown below should be reported as a potential case of anaphylaxis. This is provided as general guidance for investigators and is based on the Brighton Collaboration case definition (Rüggeberg et al 2007).

Anaphylaxis is an acute hypersensitivity reaction with multi-organ system involvement that can present as, or rapidly progress to, a severe life-threatening reaction. It may occur following exposure to allergens from a variety of sources. Anaphylaxis is a clinical syndrome characterized by:

- Sudden onset AND
- Rapid progression of signs and symptoms AND
- Involving 2 or more organ systems, as follows:
    - **Skin/mucosal**: urticaria (hives), generalized erythema, angioedema, generalized pruritus with skin rash, generalized prickle sensation, red and itchy eyes

- **Cardiovascular**: measured hypotension, clinical diagnosis of uncompensated shock, loss of consciousness or decreased level of consciousness, evidence of reduced peripheral circulation
- **Respiratory**: bilateral wheeze (bronchospasm), difficulty breathing, stridor, upper airway swelling (lip, tongue, throat, uvula, or larynx), respiratory distress, persistent dry cough, hoarse voice, sensation of throat closure, sneezing, rhinorrhea
- **Gastrointestinal**: diarrhea, abdominal pain, nausea, vomiting

##### 8.3.3. Serious Adverse Events

An AE (including an AR) is considered an SAE if, in the view of either the investigator or Sponsor, it results in any of the following outcomes:

- **Death**
  A death that occurs during the study or that comes to the attention of the investigator during the protocol-defined follow-up period must be reported to the Sponsor, whether or not it is considered related to IP.
- **Is life-threatening**
  An AE is considered life-threatening if, in the view of either the investigator or the Sponsor, its occurrence places the participant at immediate risk of death. It does not include an AE that, had it occurred in a more severe form, might have caused death.
- **Inpatient hospitalization or prolongation of existing hospitalization**
  In general, inpatient hospitalization indicates the participant was admitted to the hospital or emergency ward for at least one overnight stay for observation and/or treatment that would not have been appropriate in the physician's office or outpatient setting. The hospital or emergency ward admission should be considered an SAE regardless of whether opinions differ as to the necessity of the admission. Complications that occur during inpatient hospitalization will be recorded as an AE; however, if a complication/AE prolongs hospitalization or otherwise fulfills SAE criteria, the complication/AE will be recorded as a separate SAE.
- **Persistent or significant incapacity or substantial disruption of the ability to conduct normal life functions**
  This definition is not intended to include experiences of relatively minor medical significance such as uncomplicated headache, nausea/vomiting, diarrhea, influenza, and accidental trauma (eg, sprained ankle) which may interfere with or prevent everyday life functions but do not constitute a substantial disruption.
- **Congenital anomaly or birth defect**

- **Medically important event**
  Medical judgment should be exercised in deciding whether SAE reporting is appropriate in other situations, such as important medical events that may not be immediately life-threatening or result in death or hospitalization but may jeopardize the participant or require medical or surgical intervention to prevent one of the other outcomes listed in the above definition. These events should usually be considered serious. Examples of such medical events include allergic bronchospasm requiring intensive treatment in an emergency room or at home, blood dyscrasias or convulsions that do not result in inpatient hospitalization, or the development of drug dependency or drug abuse.

##### 8.3.4. Solicited Adverse Reactions (Part A, Blinded Phase Only)

The term “reactogenicity” refers to the occurrence and intensity of selected signs and symptoms (ARs) occurring after IP injection. The eDiary will solicit daily participant reporting of ARs using a structured checklist (Section 8.2.2). Participants will record such occurrences in an eDiary on the day of each IP injection and for the 6 days after the day of dosing.

Severity grading of reactogenicity will occur automatically based on participant entry into the eDiary according to the grading scales presented in Table 5 modified from the Toxicity Grading Scale for Healthy Adult and Adolescent Volunteers Enrolled in Preventative Vaccine Clinical Trials (DHHS 2007).

If a solicited local or systemic AR continues beyond 7 days after dosing, the participant will be prompted daily to capture solicited local or systemic AR in the eDiary until resolution. Adverse reactions recorded in eDiaries beyond Day 7 should be reviewed by the investigator either via phone call or at the following study visit. All solicited ARs (local and systemic) will be considered causally related to dosing.

**Table 5: Solicited Adverse Reactions and Grades**

| Reaction | Grade 0 | Grade 1 | Grade 2 | Grade 3 | Grade 4 |
|---|---|---|---|---|---|
| Injection site pain | None | Does not interfere with activity | Repeated use of over-the-counter pain reliever > 24 hours or interferes with activity | Any use of prescription pain reliever or prevents daily activity | Requires emergency room visit or hospitalization |
| Injection site erythema (redness) | < 25 mm/<br>< 2.5 cm | 25 - 50 mm/<br>2.5 - 5 cm | 51 - 100 mm/<br>5.1 - 10 cm | > 100 mm/<br>> 10 cm | Necrosis or exfoliative dermatitis |
| Injection site swelling/induration (hardness) | < 25 mm/<br>< 2.5 cm | 25 - 50 mm/<br>2.5 - 5 cm | 51 - 100 mm/<br>5.1 - 10 cm | > 100 mm/<br>> 10 cm | Necrosis |
| Axillary (underarm) swelling or tenderness ipsilateral to the side of injection | None | No interference with activity | Repeated use of over-the-counter (non-narcotic) pain reliever > 24 hours or some interference with activity | Any use of prescription (narcotic) pain reliever or prevents daily activity | Emergency room visit or hospitalization |
| Headache | None | No interference with activity | Repeated use of over-the-counter pain reliever > 24 hours or some interference with activity | Significant; any use of prescription pain reliever or prevents daily activity | Requires emergency room visit or hospitalization |
| Fatigue | None | No interference with activity | Some interference with activity | Significant; prevents daily activity | Requires emergency room visit or hospitalization |
| Myalgia (muscle aches all over body) | None | No interference with activity | Some interference with activity | Significant; prevents daily activity | Requires emergency room visit or hospitalization |
| Arthralgia (joint aches in several joints) | None | No interference with activity | Some interference with activity | Significant; prevents daily activity | Requires emergency room visit or hospitalization |

| Reaction | Grade 0 | Grade 1 | Grade 2 | Grade 3 | Grade 4 |
|---|---|---|---|---|---|
| Nausea/vomiting | None | No interference with activity or 1-2 episodes/ 24 hours | Some interference with activity or<br>> 2 episodes/24 hours | Prevents daily activity, requires outpatient intravenous hydration | Requires emergency room visit or hospitalization for hypotensive shock |
| Chills | None | No interference with activity | Some interference with activity not requiring medical intervention | Prevents daily activity and requires medical intervention | Requires emergency room visit or hospitalization |
| Fever (oral) | < 38.0°C<br>< 100.4°F | 38.0 – 38.4°C<br>100.4 – 101.1°F | 38.5 – 38.9°C<br>101.2 – 102.0°F | 39.0 – 40.0°C<br>102.1 – 104.0°F | > 40.0°C<br>> 104.0°F |

Any solicited AR that meets any of the following criteria must be entered into the participant's source document and must also be recorded as an AE in the participant's Adverse Event eCRF:

- Solicited local or systemic AR that results in a visit to an HCP (MAAE)
- Solicited local or systemic AR leading to the participant withdrawing from the study or the participant being withdrawn from the study by the investigator (AE leading to withdrawal)
- Solicited local or systemic AR lasting beyond 7 days post injection
- Solicited local or systemic AR that leads to participant withdrawal from IP
- Solicited local or systemic AR that otherwise meets the definition of an SAE

##### 8.3.5. Recording and Follow-up of Pregnancy

Female participants who have a positive pregnancy test at Screening should not be enrolled; participants who have a positive pregnancy test any time during the study should receive no further dosing with IP but should be asked to remain in the study and be followed-up for safety. Pregnancy testing is scheduled to occur at Screening, Blinded and Open-label Day 1, and Blinded and Open-label Day 29 (Table 16, Table 21, and Table 22).

Details of all pregnancies in female participants will be collected after the start of study treatment and until Day 759.

- If a pregnancy is reported, the investigator should inform the Sponsor within 24 hours of learning of the pregnancy and should follow the procedures outlined in this section.
- Abnormal pregnancy outcomes (eg, spontaneous abortion, fetal death, stillbirth, congenital anomalies, ectopic pregnancy) are considered SAEs.

Pregnancies occurring in participants after enrollment must be reported to Sponsor or designee within 24 hours of the site learning of its occurrence. If the participant agrees to submit this information, the pregnancy must be followed to determine the outcome, including spontaneous or voluntary termination, details of the birth, and the presence or absence of any birth defects, congenital abnormalities, or maternal and/or newborn complications. This follow-up should occur even if intended duration of the safety follow-up for the study has ended. Pregnancy report forms will be distributed to the study site to be used for this purpose. The investigator must immediately (within 24 hours of awareness) report to the Sponsor any pregnancy resulting in an abnormal outcome according to the procedures described for SAEs.

##### 8.3.6. Recording and Follow-up of an AE and/or SAE

The investigator is responsible for ensuring that all AEs and SAEs are recorded in the eCRF and reported to the Sponsor.

Solicited ARs will be collected from Day 1 through 7 days after each dose (Part A: Blinded Phase only). Other (unsolicited) AEs will be collected from Day 1 through 28 days after each dose up to Day 57 (Part A: Blinded Phase only).

Both MAAEs and SAEs will be collected from Blinded and Open-label Day 1 throughout entire study duration (Day 759 for all participants), as specified in the SoEs (Table 16, Table 17, Table 18, Table 19, Table 21, and Table 22). Any AEs occurring before receipt of IP will be analyzed separately from TEAEs.

At every study site visit or telephone contact, participants will be asked a standard question to elicit any medically related changes, including surveillance for COVID-19 symptoms, in their well-being according to the scripts provided. Participants will also be asked if they have been hospitalized, had any accidents, used any new medications, changed concomitant medication regimens (both prescription and over-the-counter medications), or had any non-study vaccinations.

In addition to participant observations, physical examination findings or other documents relevant to participant safety classified as an AE will be documented on the AE page of the eCRF.

After the initial AE/SAE report, the investigator is required to proactively follow each participant at subsequent visits/contacts. All AEs and SAEs will be treated as medically appropriate and followed until resolution, stabilization, the event is otherwise explained, or the participant is LTFU (as defined in Section 7.4).

##### 8.3.7. Time Period and Frequency for Collecting AE and SAE Information

All confirmed serious COVID-19 cases and SAEs will be recorded and reported to the Sponsor or designee immediately and under no circumstance should this exceed 24 hours, as indicated in Section 8.3.10. The investigator will submit any updated serious COVID-19 cases and SAE data to the Sponsor within 24 hours of it being available. COVID-19 cases are defined in Section 8.1.1.

Investigators are not obligated to actively seek AEs or SAEs after conclusion of the study participation (Day 759). However, if an investigator learns of any SAE, including a death, at any time after a participant has been discharged from the study and considers the event to be reasonably related to the study IP or study participation, the investigator must promptly notify the Sponsor.

##### 8.3.8. Assessment of Intensity

An event is defined as “serious” when it meets at least one of the predefined outcomes as described in the definition of an SAE (Section 8.3.3), NOT when it is rated as severe.

The severity (or intensity) of an AR or AE refers to the extent to which it affects the participant’s daily activities. The Toxicity Grading Scale for Healthy Adult and Adolescent Volunteers Enrolled in Preventative Vaccine Clinical Trials (DHHS 2007) will be used to categorize local and systemic reactogenicity events (solicited ARs), clinical laboratory test results, and vital sign measurements observed during this study. Specific criteria for local and systemic reactogenicity events are presented in Section 8.3.4.

The determination of severity for all unsolicited AEs should be made by the investigator based upon medical judgment and the definitions of severity as follows:

- Mild: These events do not interfere with the participant’s daily activities.
- Moderate: These events cause some interference with the participant’s daily activities and require limited or no medical intervention.
- Severe: These events prevent the participant’s daily activity and require intensive therapeutic intervention.

Study staff should elicit from the participant the impact of AEs on the participant’s activities of daily living to assess severity and document appropriately in the participant’s source documentation. Changes in the severity of an AE should be documented in the participant’s source documentation to allow an assessment of the duration of the event at each level of intensity to be performed. An AE characterized as intermittent requires documentation of onset and duration of each episode. An AE that fluctuates in severity during the course of the event is reported once in the eCRF at the highest severity observed.

##### 8.3.9. Assessment of Causality

The investigator's assessment of an AE's relationship to IP is part of the documentation process but is not a factor in determining what is or is not reported in the study.

The investigator will assess causality (ie, whether there is a reasonable possibility that the IP caused the event) for all AEs and SAEs. The relationship will be characterized using the following classification:

**Not related:** There is not a reasonable possibility of a relationship to the IP. Participant did not receive the IP OR temporal sequence of the AE onset relative to administration of the IP is not reasonable OR the AE is more likely explained by another cause than the IP.

**Related:** There is a reasonable possibility of a relationship to the IP. There is evidence of exposure to the IP. The temporal sequence of the AE onset relative to the administration of the IP is reasonable. The AE is more likely explained by the IP than by another cause.

##### 8.3.10. Reporting Adverse Events

The investigator is responsible for reporting all AEs that are observed or reported during the study, regardless of their relationship to IP or their clinical significance. If there is any doubt as to whether a clinical observation is an AE, the event should be reported.

All unsolicited AEs reported or observed during the study will be recorded on the AE page of the eCRF. Information to be collected includes, type of event, time of onset, investigator -specified assessment of severity (impact on activities of daily living) and relationship to IP, time of resolution of the event, seriousness, as well as any required treatment or evaluations, and outcome. The unsolicited AEs resulting from concurrent illnesses, reactions to concurrent illnesses, reactions to concurrent medications, or progression of disease states must also be reported. All AEs will be followed until they are resolved or stable or judged by the investigator to be not clinically significant. The Medical Dictionary for Regulatory Activities (MedDRA) will be used to code all unsolicited AEs.

Any medical condition that is present at the time that the participant is screened but does not deteriorate should not be reported as an unsolicited AE. However, if it deteriorates at any time during the study, it should be recorded as an unsolicited AE.

Any AE or COVID-19 case considered serious by the investigator or that meets SAE criteria (Section 8.3.3) must be reported to the Sponsor immediately (within 24 hours of becoming aware of the SAE or COVID-19 case). The investigator will assess whether there is a reasonable possibility that the IP caused the SAE. The Sponsor will be responsible for notifying the relevant regulatory authorities of any SAE as outlined in the 21 US CFR Parts 312 and 320. The investigator is responsible for notifying the institutional review board (IRB) directly.

If the eCRF is unavailable at the time of the SAE, the following contact information is to be used for SAE reporting:

- SAE Mailbox:
- SAE Hotline (USA and Canada): +1-866-599-1341
- SAE Fax line (USA and Canada): +1-866-599-1342

##### 8.3.11. Regulatory Reporting Requirements for SAEs

Prompt notification by the investigator to the Sponsor of a SAE is essential so that legal obligations and ethical responsibilities towards the safety of participants and the safety of a study intervention under clinical investigation are met.

The Sponsor has a legal responsibility to notify both the local regulatory authority and other regulatory agencies about the safety of a study intervention under clinical investigation. The Sponsor will comply with country-specific regulatory requirements relating to safety reporting to the regulatory authority, IRBs, and investigators.

Investigator safety reports must be prepared for suspected unexpected serious ARs according to local regulatory requirements and Sponsor policy and forwarded to investigators as necessary.

An investigator who receives an investigator safety report describing a SAE or other specific safety information (eg, summary or listing of SAEs) from the Sponsor will review and then file it along with the IB and will notify the IRB/IEC, if appropriate according to local requirements.

#### 8.4. Monitoring Committees

##### 8.4.1. Protocol Safety Review Team

A Protocol Safety Review Team will be formed to review interim and cumulative blinded safety data on a regular basis with a remit to escalate concerns to the DSMB.

##### 8.4.2. Data and Safety Monitoring Board

An independent DSMB will periodically review blinded and unblinded data, including both safety and cases of COVID-19 at scheduled data review meetings and at 2 planned IAs.

- In addition to blinded and unblinded review of safety data, at each data review meeting the DSMB will review the numbers and rate of COVID-19 cases, including rate of severe COVID disease with prespecified thresholds for imbalance in the treatment groups which would trigger halting rules.
- At the IA, the DSMB will review the IA results and make recommendations to an Oversight Group in terms of study results reporting and unblinding based on the boundaries of early efficacy as described in Section 9.6 of the protocol. The Oversight Group will be comprised of a voting member each from the Sponsor, Biomedical Advance Research and Development Authority (BARDA), and NIAID.
- The DSMB will monitor the study for non-efficacy at the IA. The boundary for non-efficacy is non-binding and will be provided in the DSMB analysis plan.

- The DSMB will also monitor the study for vaccine harm based on severe COVID-19. Continuous harm monitoring will be provided for COVID-19 and severe COVID-19 separately. For harm monitoring, cases will be counted starting after the first dose of study vaccination. Boundaries will be provided based on the exact 1-sided binomial tests conditional on the total number of cases under the assumption of VE=0%. If the prespecified stopping boundary is reached for either COVID-19 or severe COVID-19, then the unblinded statisticians will immediately inform the DSMB that the harm rules have been met. Details will be provided in the DSMB analysis plan.
- The boundaries are considered guidelines, ie, a recommendation to modify the study would not be based solely on statistical rules, as many other factors (ie, totality of the data from the study including additional efficacy, safety and immunogenicity endpoints as well as data external to the study) may be part of the decision process. In the case of a recommendation to continue the trial regardless of crossing the boundaries for efficacy or inefficacy, the reason for disregarding the boundary must be documented in the meeting minutes and communicated to the Sponsor.

After each data review meeting or IA, the DSMB will make a recommendation to the Sponsor through an Oversight Group to take one of the following courses of action:

- Stop further enrollment due to meeting criteria for early efficacy or due to a safety concern.
- Pause enrollment and consider a change in study design.
- Continue enrollment and/or study conduct as planned.

The Sponsor may also request that the DSMB conduct ad hoc reviews of safety events from this study or other data, including new nonclinical or clinical information related to mRNA-1273 external to this study. The DSMB will review all available study data to adjudicate such events in accordance with the DSMB charter.

The DSMB composition, its remit, and frequency of data review will be further defined in the DSMB charter and analysis plan.

##### 8.4.3. Adjudication Committee

An Adjudication Committee (AC) will be assembled for the purpose of reviewing potential cases to determine if the criteria for the primary and secondary endpoints have been met. The AC will remain blinded to treatment assignment. The AC composition, its remit, and frequency of data review will be further defined in a Charter.

#### 8.5. Management of Overdose

As the study treatment is to be administered by a healthcare professional, it is unlikely that an overdose will occur. Dose deviations will be tracked as protocol deviations (Section 11.2.8).

#### 8.6. Pharmacokinetics

Pharmacokinetic parameters are not evaluated in this study.

#### 8.7. Pharmacodynamics

Pharmacodynamic parameters are not evaluated in this study.

#### 8.8. Exploratory Assessments and Biomarkers

Exploratory assessments may include assessment of serologic markers of disease severity, immune response to SARS-CoV-2, RT-PCR of NP swab, or nasal swab, or saliva samples, and genetic sequences of SARS-CoV-2 strains isolated from participants' samples.

#### 8.9. Medical Resource Utilization and Health Economics

Medical resource utilization and health economics parameters are not evaluated in this study.

### 9. STATISTICAL CONSIDERATIONS

This section summarizes the planned statistical analysis strategy and procedures for the study. The details of statistical analysis will be provided in the SAP, which will be finalized before the clinical database lock for the study and treatment unblinding. If, after the study has begun, but prior to any unblinding, changes are made to primary and/or key secondary objectives/hypotheses, or the statistical methods related to those hypotheses, then the protocol will be amended (consistent with ICH Guideline E9). Changes to other secondary or exploratory analyses made after the protocol has been finalized, along with an explanation as to when and why they occurred, will be listed in the SAP or Clinical Study Report (CSR) for the study. Ad hoc exploratory analyses, if any, will be clearly identified in the CSR. Statistical analysis strategy incorporating the Open-Label Phase will be provided in a SAP.

#### 9.1. Blinding and Responsibility for Analyses

Blinding during the Part A, the Blinded Phase of the study will be conducted as described in Section 6.2.8. The Sponsor Biostatistics department or designee will generate the randomized allocation schedule(s) for study treatment assignment. Randomization will be implemented via an IRT.

Planned interim and primary analyses are described in Section 9.6. Participant-level unblinding will be restricted to an independent unblinded statistician and, as needed, a statistical programmer performing the IAs, who will have no other responsibilities associated with the study.

In addition to the routine study monitoring outlined in this protocol, an external DSMB will review interim data to safeguard the interests of clinical study participants and to enhancing the integrity of the study. The DSMB will review treatment-level results of the IAs, provided by the independent unblinded statistician. Limited additional Sponsor personnel may be unblinded to the treatment-level results of the IAs, if required, in order to act on the recommendations of the DSMB. The extent to which individuals are unblinded with respect to results of IAs will be documented. Depending on the recommendation of the DSMB, the Sponsor may prepare a regulatory submission after an IA. In this case, pre-identified Sponsor members including the analysis and reporting team will be unblinded to treatment assignments and remain unblinded for the remainder of the study. Participants and investigators will remain blinded.

#### 9.2. Statistical Hypotheses

For the primary efficacy objective, the null hypothesis of this study is that the VE of mRNA-1273 to prevent first occurrence of COVID-19 is ≤ 30% (ie, $H_0^{efficacy}$: VE ≤ 0.3). The study will be considered to meet the primary efficacy objective if the corresponding CI of VE rules out 30% at either one of the IAs or at the primary analysis.

Vaccine efficacy is defined as the percent reduction in the hazard of the primary endpoint (mRNA-1273 vs. placebo). Equivalently, the null hypothesis is:

$H_0^{efficacy}$: HR ≥ 0.7 (equivalently, proportional hazards VE ≤ 0.3).

A stratified Cox proportional hazard model will be used to assess the magnitude of the treatment group difference (ie, HR) between mRNA-1273 and placebo at a 1-sided 0.025 significance level.

The primary analysis population for efficacy will be the PP Set, defined in Section 9.4. In the primary analysis of efficacy, cases will be counted starting 14 days after the second dose of IP.

#### 9.3. Sample Size Determination

The sample size is driven by the total number of cases to demonstrate VE (mRNA-1273 vs. placebo) to prevent COVID-19. Under the assumption of proportional hazards over time and with 1:1 randomization of mRNA-1273 and placebo, a total of 151 COVID-19 cases will provide 90% power to detect a 60% reduction in hazard rate (60% VE), rejecting the null hypothesis $H_0$: VE ≤ 30%, with 2 IAs at 35% and 70% of the target total number of cases using a 1-sided O'Brien-Fleming boundary for efficacy and a log-rank test statistic with a 1-sided false positive error rate of 0.025. The total number of cases pertains to the PP Set accruing at least 14 days after the second dose. There are 2 planned IAs in this study, which will be performed when approximately 35% and 70% of the target total number of cases have been observed. Approximately 30,000 participants will be randomized with the following assumptions:

- The target VE against COVID-19 is 60% (with 95% CI lower bound ruling out 30%, rejecting the null hypothesis $H_0$: VE ≤ 30%)
- A 6-month COVID-19 incidence rate of 0.75% in the placebo arm
- An annual dropout rate of 2% (loss of evaluable participants)
- Two IAs at 35% and 70% of total target cases across the 2 treatment groups with O'Brien-Fleming boundaries for efficacy monitoring
- 3-month uniform accrual
- Approximately 15% of participants will be excluded from the PP population, and participants are at risk for COVID-19 starting 14 days after the second dose

Table 6 provides sample size with 90% power to demonstrate VE on COVID-19.

**Table 6: Conditions and Sample Size to Demonstrate Vaccine Efficacy**

| Target VE | Lower Bound | Randomization Ratio | Total # of Cases | 6-Month Incidence Rate | | Total Sample Size[a] |
|---|---|---|---|---|---|---|
| -- | -- | -- | -- | Placebo | mRNA-1273 | -- |
| 60% | 30% | 1:1 | 151 | 0.75% | 0.30% | 30,000 |

[a] Sample size to account for 15% participants to be excluded from the PP Set (eg, seropositive at baseline, have not received planned IP).

The sample size is calculated using R package gsDesign (Anderson 2020).

Under these above assumptions including 6-month incidence rate of 0.75% on placebo, with 30,000 participants, it will take approximately 5, 8, and 10 months from study start (first subject first dose), respectively, to accrue 35% (approximately 53), 70% (approximately 106) and 100% (151) of the target number of cases in the PP Set.

Figure 5 shows the power of the primary efficacy endpoint under true VE at the 2 planned IAs and the primary efficacy analysis assuming a total of 151 events.

**Figure 5: Boundary Crossing Probabilities by Effect Size**

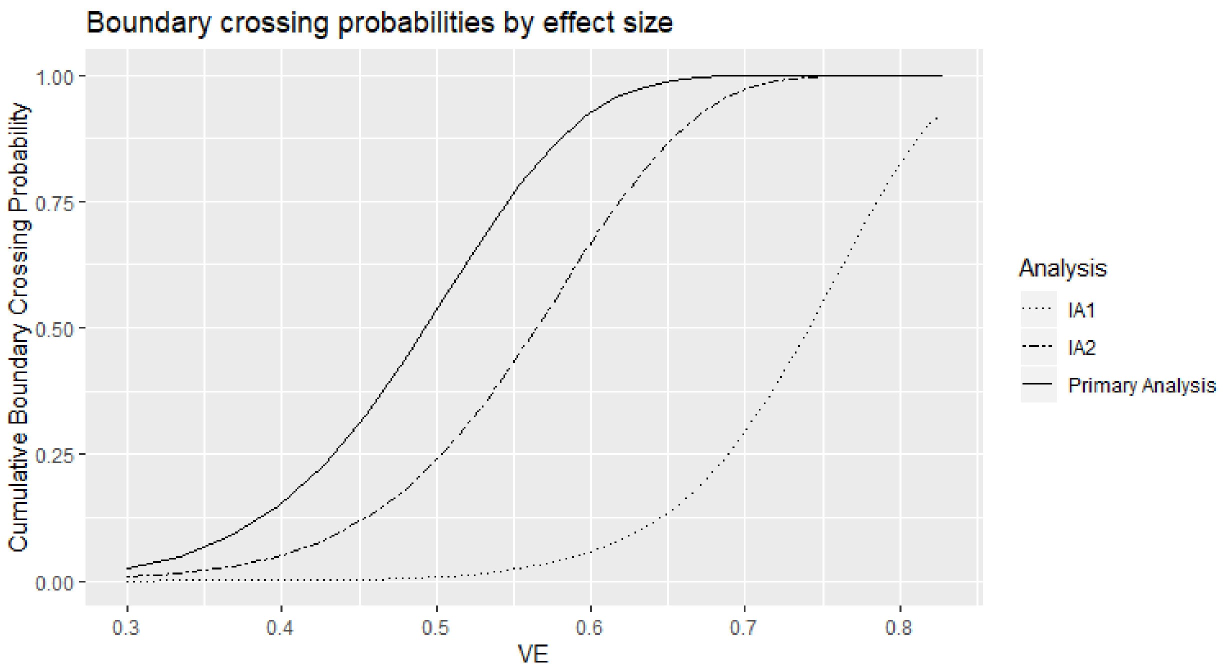

The Sponsor may adjust the size of the study or duration of follow-up based on the blinded review of the total number of cases of COVID-19 accrued during the study, in addition to estimated percentages of study participants with serologic evidence of SARS-CoV-2 infection at baseline.

##### 9.3.1. Power for Selected Secondary Efficacy Endpoints

For the secondary objective on VE against virologically confirmed SARS-CoV-2 infection or COVID-19 regardless of symptomology or severity (COV-INF), the study will have ≥ 90% power to demonstrate the VE is above 30% (to reject null hypothesis VE ≤ 30%) at 1-sided alpha of 2.5% if the true VE to prevent COV-INF is 60% because every COVID-19 endpoint is necessarily a COV-INF endpoint.

For the secondary objective on VE against severe COVID-19, Table 7 provides power of demonstrating VE based on a total of 30 and 60 events under different scenarios of true VE and VE criteria.

**Table 7: Power of Demonstrating Vaccine Efficacy Against Severe COVID-19**

| True VE | Total No. of Severe COVID-19 Cases | Severe COVID-19 (Secondary Objective) | |
|---|---|---|---|
| | | To Reject Null Hypothesis | Power |
| 60% | 30 | VE≤ 0% | 70.90% |
| | | VE≤ 10% | 60.30% |
| | | VE≤ 20% | 47.50% |
| 70% | | VE≤ 0% | > 90% |
| | | VE≤ 10% | 85.30% |
| | | VE≤ 20% | 76.60% |
| 80% | | VE≤ 0% | > 90% |
| | | VE≤ 10% | > 90% |
| | | VE≤ 20% | > 90% |
| 60% | 60 | VE≤ 0% | > 90% |
| | | VE≤ 10% | 88.10% |
| | | VE≤ 20% | 76.60% |
| 70% | | VE≤ 0% | > 90% |
| | | VE≤ 10% | > 90% |
| | | VE≤ 20% | > 90% |
| 80% | | VE≤ 0% | > 90% |
| | | VE≤ 10% | > 90% |
| | | VE≤ 20% | > 90% |

Abbreviation: VE = vaccine efficacy.

#### 9.4. Analysis Populations

Analysis populations for statistical analyses are Randomization Set, Full Analysis Set (FAS), modified intent-to-treat (mITT) set, PP Set, Immunogenicity Subset, Solicited Safety Set, and Safety Set, as shown in Table 8.

**Table 8: Populations for Analyses**

| Population | Description |
|---|---|
| Randomization Set | All participants who are randomized, regardless of the participants' treatment status in the study. |
| Full Analysis Set (FAS) | All randomized participants who received at least one dose of IP.<br>Participants will be analyzed according to the group to which they were randomized. |
| Modified Intent-to-Treat (mITT) Set | All participants in the FAS who had no immunologic or virologic evidence of prior COVID-19 (ie, negative NP swab test at Day 1, and/or bAb against SARS-CoV-2 nucleocapsid below LOD or LLOQ) at Day 1 before the first dose of IP.<br>Participants will be analyzed according to the group to which they were randomized. |
| Per-protocol (PP) Set | All participants in the mITT Set who received planned doses of IP per schedule and have no major protocol deviations, as determined and documented by Sponsor prior to database lock and unblinding, that impact critical or key study data.<br>Participants will be analyzed according to the group to which they were randomized. |
| Immunogenicity Subset | All participants in the FAS who were sampled into a subset for characterizing mRNA-1273 immunogenicity and had a valid immunogenicity test result prior to the first dose of IP and at least 1 valid result after the first dose of IP. Participants in the subset who had major protocol deviations that impact critical or key immunogenicity or study data may be excluded from the Immunogenicity Subset. The details of the Immunogenicity Subset will be documented prior to analysis of immunogenicity data. |
| Solicited Safety Set | The Solicited Safety Set consists of all randomized participants who received at least one dose of IP and contributed any solicited AR data.<br>The Solicited Safety Set will be used for the analyses of solicited ARs and participants will be included in the treatment group corresponding to the IP that they actually received. |
| Safety Set | All randomized participants who received at least one dose of IP. The Safety Set will be used for all analyses of safety except for the solicited ARs.<br>Participants will be included in the treatment group corresponding to the IP that they actually received. |

#### 9.5. Statistical Analyses

This section provides a summary of the planned statistical analyses of the primary and secondary endpoints.

The overall Type I error rate for the primary endpoint at the IAs and the primary analysis is strictly controlled at 2.5% (1-sided) based on the Lan-DeMets O'Brien-Fleming approximation spending function (see Section 9.6 for details). The primary efficacy results that will be considered statistically significant after consideration of the strategy for controlling the Type I error as described in Section 9.6. Statistical significance of the primary efficacy endpoint can be achieved at either one of the IAs or at the primary analysis. A sequential/hierarchical testing procedure will be used to control type 1 error rate over the primary efficacy endpoint and the secondary efficacy endpoints. Secondary efficacy endpoints will only be tested when the primary efficacy endpoint achieves statistical significance. Multiplicity adjustments among the secondary efficacy endpoints may be performed for secondary efficacy endpoints, in that case, will be specified in the SAP.

No formal multiple comparison adjustments will be employed for multiple safety endpoints or multiple efficacy endpoints. Nominal p-values and confidence intervals (CIs) may be computed for other efficacy analyses without controlling for multiplicity as a measure of VE.

##### 9.5.1. Efficacy Analyses

Efficacy analyses will be performed using the FAS, mITT and PP populations, and participants will be included in the treatment group to which they are randomized. The primary analysis population will be the PP Set.

Table 9 summarizes the analysis approach for primary and secondary efficacy endpoints. Sensitivity analysis methods are described for each endpoint as applicable.

**Table 9: Statistical Analysis Methods of Efficacy Endpoints**

| Endpoint | Statistical Analysis Methods |
|---|---|
| **Primary endpoint:**<br>Vaccine Efficacy (VE) of mRNA-1273 to prevent COVID-19 | • Primary analysis: VE will be estimated with 1 - HR (mRNA-1273 vs placebo) using a Cox proportional hazard regression model with treatment group as a fixed effect and adjust for stratification factor based on the PP Set, with cases counted starting 14 days after the second dose of IP.<br>• Analysis using the same model based on the mITT Set.<br>• Sensitivity analysis using the same model based on the PP Set, with cases counted starting either immediately after the second dose of IP or immediately after the first dose of IP. |

| Endpoint | Statistical Analysis Methods |
|---|---|
| | • Subgroup analysis of the primary efficacy endpoint will be performed to assess consistency of VE, such as in the age groups ≥ 18 and < 65 years and ≥ 65 years.<br>• Supportive analysis of VE to be estimated with 1 - ratio of incidence rates with 95% CI using the exact method conditional upon the total number of cases.<br>• Supportive analysis of cumulative incidence VE . |
| **Secondary endpoints:**<br>• Vaccine efficacy of mRNA-1273 to prevent severe COVID-19<br>• Vaccine efficacy of mRNA-1273 to prevent serologically confirmed SARS-CoV-2 infection or COVID-19 regardless of symptomatology or severity<br>• Vaccine efficacy of mRNA-1273 to prevent COVID-19 using a secondary definition of symptoms<br>• Vaccine efficacy of mRNA-1273 to prevent death due to COVID-19<br>• Vaccine efficacy of mRNA-1273 to prevent COVID-19 after the first dose of IP<br>• Vaccine efficacy of mRNA-1273 to prevent asymptomatic SARS-CoV-2 infection | Similar analysis method as for the primary endpoint analysis.<br>For each of the secondary endpoints:<br>• Primary analysis: VE will be estimated with 1 - HR (mRNA-1273 vs placebo) using a Cox proportional hazard regression model with treatment group as a fixed effect and adjusting for stratification factor based on the PP Set, with cases counted starting 14 days after the second dose of IP.<br>• Analysis using the same model based on the mITT Set.<br>• Sensitivity analyses with cases counted starting immediately after the second dose of IP, 14 days after the first dose of IP, immediately after the first dose of IP, and immediately after randomization.<br>• Vaccine efficacy and 95% CI based on the case incidence will be estimated with 1 - ratio of incidence rates using the exact method conditional upon the total number of cases. |
| • Vaccine efficacy of mRNA-1273 to prevent COVID-19 in all study participants, regardless of evidence of prior SARS-CoV-2 infection | The FAS population will be used for this secondary objective, using similar analysis methods as for the primary endpoint analysis.<br>• Primary analysis: VE will be estimated with 1 - HR (mRNA-1273 vs placebo) using a Cox proportional hazard regression model with treatment group as a fixed effect and adjusting for stratification factor based on the FAS, with cases counted starting 14 days after the second dose of IP.<br>• Sensitivity analyses with cases counted starting immediately after the second dose of IP, 14 days after the first dose of IP, immediately after the first dose of IP, and immediately after randomization. |

###### 9.5.1.1. Efficacy Analysis on Primary Endpoint

To assess the primary efficacy endpoint of VE of mRNA-1273 in preventing the first occurrence of COVID-19 from 14 days after second dose of IP, Cox proportional hazards regression will be used to estimate proportional hazards VE (PH VE), measured by one minus the HR (mRNA-1273 vs. placebo), with a 2-sided score-based 95% CI and 2-sided p-value for testing $H_0$: VE ≤ 30%.

Vaccine efficacy is defined as the percent reduction in the hazard of the primary endpoint (mRNA-1273 vs placebo). The VE will be estimated using one minus the HR (mRNA-1273 vs placebo) estimand. A stratified Cox proportional hazard model will be used to assess the magnitude of the treatment group difference (ie, HR) between mRNA-1273 and placebo. The HR and its 95% CI from the stratified Cox model with Efron's method of tie handling and with treatment group as covariate will be reported. The same stratification factors used for randomization will be applied to the stratified Cox model.

For the primary efficacy endpoints, participants without documented COVID-19 will be censored at the last study assessment date. Potential intercurrent events may include: 1) death unrelated to COVID-19 and 2) early COVID-19 up to 14 days after second study dose.

In the estimand of the primary analysis on the primary endpoint, a hypothetical strategy will be used for death unrelated to COVID-19; a hypothetical strategy will also be used for early COVID-19, where the time to COVID-19 will be censored at the onset day of early case. The details of intercurrent event description and estimand strategies are presented in Section 11.4.1.

For the primary efficacy analysis, cases will be counted starting 14 days after the second vaccination. Sensitivity analyses with cases counted immediately after the second vaccination, and after randomization will also be carried out.

Analyses of the primary endpoint will be also performed based on the mITT Set using the same methods described above.

For the primary efficacy analysis, cases will be counted starting 14 days after the second dose of IP. Sensitivity analyses with cases counted starting immediately after the second dose of IP and starting immediately after randomization will also be carried out.

Subgroup analysis of the primary efficacy endpoint will be performed in selected subgroups, such as age groups ≥18 and <65 year and ≥65 years to assess consistency of VE as described in Section 9.5.5.

As a supportive analysis, VE will also be estimated by one minus the infection rate ratio, where the number of cases (ie, participants with first occurrence of COVID-19) will be used and the CI will be computed using the exact method conditional upon the total number of cases. Cumulative incidence VE, one minus the ratio of cumulative incidences (mRNA-1273 vs placebo) of COVID--19, may also be assessed, the cumulative incidence for each arm will be estimated using a covariate adjustment method based on Zeng (2004) that makes use of baseline characteristics.

Additional analysis to evaluate VE against COVID-19 incorporating duration and presence/severity of symptoms will also be performed; the details will be provided in the SAP.

###### 9.5.1.2. Efficacy Analysis on Secondary Endpoint

For each of the below secondary objectives:

- Vaccine efficacy to prevent severe COVID-19
- Vaccine efficacy to prevent serologically confirmed SARS-CoV-2 infection or COVID-19 regardless of symptomology or severity
- Vaccine efficacy to prevent COVID-19 using a broad definition of symptoms
- Vaccine efficacy to prevent death caused by COVID-19
- Vaccine efficacy to prevent asymptomatic SARS-CoV-2 infection

For each of the above secondary objectives, the same Cox proportional hazard model described above for the primary objective will be applied using the PP Set, with cases counted starting 14 days after the second dose of IP. Sensitivity analyses with cases counted starting after the second dose of IP, 14 days after the first dose of IP, after the first dose of IP, and after randomization will also be performed.

The same model will be applied using the mITT population with cases counted starting 14 days after the second dose of IP.

Vaccine efficacy will be estimated with 1- ratio of incidence rates with the 95% CI using the exact method conditional upon the total number of cases.

Vaccine efficacy to prevent COVID-19 after the first dose of IP

The same Cox proportional hazard model described above for the primary objective will be applied using the PP Set, with cases counted starting 14 days after the first dose of IP.

Vaccine efficacy to prevent COVID-19 regardless of prior SARS-CoV-2 infection

The FAS will be used for analysis to evaluate VE to prevent COVID-19 regardless of prior SARS-CoV-2 infection. The same methods described above for the primary objective will be applied with cases counted starting 14 days after the second dose of IP. Sensitivity analyses with cases counted starting after the second dose of IP, 14 days after the first dose of IP, the first dose of IP, and randomization will also be performed.

Vaccine efficacy will be estimated with 1- ratio of incidence rates with the 95% CI using the exact method conditional upon the total number of cases.

##### 9.5.1.3. Long-term Efficacy Analysis

Long-term efficacy will be evaluated after the primary analysis by including data collected in the Open-Label Observational Phase. Long-term efficacy data will be summarized descriptively by treatment cohort described in Table 10 without cohort comparison. The long-term efficacy endpoints are listed in Table 11 with the same case definitions specified in Table 1. In the primary approach, cases will be counted starting 14 days after the second dose of IP for participants in treatment cohorts of mRNA-1273 and Placebo, or starting 14 days after the second dose of mRNA-1273 for participants in the Placebo-mRNA-1273 Cohort. Sensitivity analyses with cases starting from the second dose, 14 days after the first dose, or first dose of IP for participants in the cohorts of mRNA-1273 and placebo, or mRNA-1273 for participants in the Placebo-mRNA-1273 Cohort will be provided. Incidences of cases assessed by numbers, rates, and 2-sided 95% CI based on the exact method adjusting for person-time will be summarized by treatment cohort. The Kaplan-Meier analysis will be used to estimate cumulative incidences of time to first cases by treatment cohort. Long-term efficacy analysis will be performed using FAS for the case of COVID-19 regardless of prior SARS-CoV-2 infection, and long-term efficacy analysis of other cases will be conducted using the PP Set and mITT Set.

In addition, efficacy analyses based on data collected in the Open-Label Observational Phase only will be provided in the SAP.

**Table 10: Treatment Cohorts for the Long-term Efficacy Analyses**

| Treatment Cohort | Description |
|---|---|
| **mRNA-1273** | Participants randomized to mRNA-1273 in the Blinded Phase. |
| **Placebo** | Participants randomized to Placebo in the Blinded Phase and did not crossover to mRNA-1273 in the Open-Label Observational Phase. |
| **Placebo-mRNA-1273** | Participants randomized to Placebo in the Blinded phase and crossed over to mRNA-1273 in the Open-Label Observational Phase. |

**Table 11: Long-term Efficacy Endpoints**

| Long-term Efficacy Endpoint |
|---|
| Cases of COVID-19 starting 14-days after the second injection of:<br>• IP for participants in the mRNA-1273 Cohort and the Placebo Cohort,<br>or<br>• mRNA-1273 for participants in the Placebo-mRNA-1273 Cohort. |

| **Long-term Efficacy Endpoint** |
|---|
| Cases of severe COVID-19 starting 14-days after the second injection of:<br>• IP for participants in the mRNA-1273 Cohort and the Placebo Cohort,<br>or<br>• mRNA-1273 for participants in the Placebo-mRNA-1273 Cohort. |
| Cases of either COVID-19 or SARS-CoV-2 infection starting 14-days after the second injection of:<br>• IP for participants in the mRNA-1273 Cohort and the Placebo Cohort,<br>or<br>• mRNA-1273 for participants in the Placebo-mRNA-1273 Cohort. |
| Cases with a secondary (less restrictive) definition of COVID-19 starting 14-days after the second injection of:<br>• IP for participants in the mRNA-1273 Cohort and the Placebo Cohort,<br>or<br>• mRNA-1273 for participants in the Placebo-mRNA-1273 Cohort. |
| Cases of death due to a cause directly attributed to a complication of COVID-19, starting 14-days after the second injection of:<br>• IP for participants in the mRNA-1273 Cohort and Placebo Cohort,<br>or<br>• mRNA-1273 for participants in the Placebo-mRNA-1273 Cohort. |
| Cases of COVID-19 starting 14-days after the first injection of:<br>• IP for participants in the mRNA-1273 Cohort and the Placebo Cohort,<br>or<br>• mRNA-1273 for participants in the Placebo-mRNA-1273 Cohort. |
| Regardless of evidence of prior SARS-CoV-2 infection determined by serologic titer against SARS-CoV-2 nucleocapsid, cases of COVID-19 starting 14-days after the second injection of:<br>• IP for participants in the mRNA-1273 Cohort and the Placebo Cohort,<br>or<br>• mRNA-1273 for participants in the Placebo-mRNA-1273 Cohort. |
| Cases of SARS-CoV-2 infection in the absence of symptoms defining COVID-19 starting 14-days after the second injection of:<br>• IP for participants in the mRNA-1273 Cohort and the Placebo Cohort,<br>or<br>• mRNA-1273 for participants in the Placebo-mRNA-1273 Cohort. |

##### 9.5.2. Safety Analyses

All safety analyses will be based on the Safety Set, except summaries of solicited ARs, which will be based on the Solicited Safety Set. All safety analyses will be provided by treatment group, and by treatment cohort as applicable, unless otherwise specified. Pregnancies and their known outcomes will be summarized (Section 8.3.5).

###### 9.5.2.1. Adverse Events

Safety and reactogenicity will be assessed by clinical review of all relevant parameters.

The number and percentage of participants with any solicited local AR, with any solicited systemic AR, and with any solicited AR during the 7-day follow-up period after each dose (Blinded Phase only) will be provided. A 2-sided 95% exact CI using the Clopper-Pearson method will be also provided for the percentage of participants with any solicited AR for each treatment group.

Number and percentage of participants with unsolicited AEs, SAEs, MAAEs, Grade 3 or higher ARs and AEs, and AEs leading to discontinuation from IP or withdrawal from the study will be summarized. Unsolicited AE will be presented by MedDRA preferred term and system organ class.

Number of events of solicited ARs, unsolicited AEs/SAEs, and MAAEs will be reported in summarization tables accordingly.

For all other safety parameters, descriptive summary statistics will be provided, and Table 12 summarizes analysis strategy for safety parameters. Further details will be described in the SAP.

**Table 12: Analysis Strategy for Safety Parameters**

| Safety Endpoint | Number and Percentage of Participants, Number of Events | 95% CI |
|---|---|---|
| Any Solicited AR (overall and by local, systemic) | X | X |
| Any Unsolicited AE | X | |
| Any SAE | X | |
| Any Unsolicited MAAE | X | |
| Any Unsolicited Treatment-Related AE | X | |
| Any Treatment-Related SAE | X | |
| Discontinuation due to AE | X | |
| Any Grade 3 and above AE | X | |
| Any Treatment-Related Grade 3 and above AE | X | |

Notes: 95% CI using the Clopper-Pearson method, X = results will be provided. Unsolicited AEs will be summarized by System Organ Class and Preferred Term coded by MedDRA.

###### 9.5.2.2. Baseline Descriptive Statistics

Demographic variables and baseline characteristics will be summarized by treatment group, and by treatment cohort as applicable, by descriptive statistics (mean, standard deviation for continuous variable, and number and percentage for categorical variables).

##### 9.5.3. Immunogenicity Analyses

The secondary immunogenicity endpoints will be analyzed using the Immunogenicity Subset by treatment group, by treatment cohort as applicable, and by baseline SARS-CoV-2 serostatus, unless otherwise specified. Details for immunogenicity analyses on data collected in the Open-Label Observational Phase will be provided in the SAP.

The SAP will describe the complete set of immunogenicity analyses, including the approach to sample individuals into an Immunogenicity Subset for characterizing mRNA-1273 immunogenicity and assessing immunological correlates of risk and protection.

Data from quantitative immunogenicity assays will be summarized for each treatment group using positive response rates and geometric means with 95% CIs, for each timepoint for which an assessment is performed. Data from qualitative (ie, yielding a positive or negative result) assays will be summarized by tabulating the frequency of positive responses for each assay by group at each timepoint that an assessment is performed. Analyses will focus on the 2 key immunogenicity time points and the change in marker response between them: Day 1 before the first dose of IP and Day 57 (28 days after the second dose of IP). The SAP will describe the complete set of immunogenicity analyses.

Quantitative levels or GMT of specific bAb with corresponding 95% CI at each timepoint and GMFR of specific bAb with corresponding 95% CI at each post-baseline timepoint over pre-dose baseline at Day 1 will be provided by study arm. Descriptive summary statistics including median, minimum, and maximum will also be provided.

GMT of specific nAb with corresponding 95% CI at each timepoint and GMFR of specific nAb with corresponding 95% CI at each post-baseline timepoint over pre-dose baseline at Day 1 will be provided by study arm. Descriptive summary statistics including median, minimum, and maximum will also be provided. For summarizations of group values, antibody values reported as below the LLOQ will be replaced by 0.5 × LLOQ. Values that are greater than the ULOQ will be converted to the ULOQ.

The number and percentage of participants with a fold rise $\geq 2$, $\geq 3$, and $\geq 4$ of serum SARS-CoV-2-specific nAb titers and participants with seroconversion due to vaccination from baseline will be provided with 2-sided 95% CI using the Clopper-Pearson method at each post-baseline timepoint.

Seroconversion due to vaccination at a participant level may be defined as a change from below the LLOQ to equal or above LLOQ, or at least 4-fold rise in terms of neutralizing antibody or vaccine antigen-specific bAb in participants with pre-existing bAb or nAb. The definition of seroconversion will be finalized and documented in the SAP prior to analyzing the immunogenicity data.

The GMT of specific nAb for each group and the geometric mean ratio (GMR) of mRNA-1273 versus placebo with corresponding 2-sided 95% CI will be estimated at each study timepoint using an analysis of covariance (ANCOVA) model with the treatment group and baseline values, if applicable, as explanatory variables, the analysis may adjust for the stratification factor (Table 13).

**Table 13: Immunogenicity Endpoints and Statistical Methods**

| Endpoint | Statistical Analysis Methods |
|---|---|
| Specific bAb and nAb titers/values | • GMT of each group, GMR (mRNA-1273 vs. Placebo)<br>• GMT estimated by the ANCOVA model |
| Fold rise | • GMFR - descriptive statistics<br>• Binomial endpoints of fold rise ≥2, 3, and 4, and seroconversion due to vaccination - the Clopper-Pearson method |

##### 9.5.4. Exploratory Analyses

The endpoint of viral infection kinetics will be assessed by determining the number of days until testing of nasal swab or saliva samples becomes negative after COVID-19 is established.

The endpoint of duration of symptoms will be assessed by determining the total number of days that a study participant with COVID-19 remains symptomatic through daily assessments after diagnosis.

Vaccine efficacy to prevent all-cause mortality will be assessed by similar analysis methods as used for the primary endpoint analysis, using PP Set, mITT Set, and FAS. All deaths, regardless of cause from the time of randomization, will be included. If the number of deaths becomes large enough to warrant analysis, the same Cox proportional hazard model described above for the primary objective will be applied using the PP Set, the mITT Set, and the FAS. Death, regardless of cause, from randomization will be included.

This endpoint of BOD is defined based on the post SARS-CoV-2 infection follow-up. A BOD score will be used to reflect the severity of symptoms with maximum score at COVID-19 death (Table 14). All other analyses of exploratory endpoints will be described in the SAP before database lock.

**Table 14: Burden of Disease Score**

| Patient State | BOD Score |
|---|---|
| Uninfected/Asymptomatic infection | 0 |
| Symptomatic without hospitalization | 1 |
| Hospitalization | 2 |
| Death | 3 |

###### 9.5.5. Subgroup Analyses

To determine whether the VE is consistent across various subgroups, the VE and its 95% CI may be estimated using the similar model within each category of the following classification variables.

- Age groups: ≥ 18, < 65, and ≥ 65
- Age and health risk for severe disease: ≥ 18 and < 65 and not at risk; ≥ 18 and < 65 and at risk, and ≥ 65
- Sex (female, male)
- Race
- At risk for severe COVID-19 illness (Section 6.2.1.1)

Subgroup analysis for the long-term efficacy may be performed descriptively by treatment cohort within above categories.

##### 9.6. Interim Analyses

There are 2 planned IAs at 35% and 70% of total target cases across the 2 treatment groups. The primary objective of the IAs is for early detection of reliable evidence that VE is above 30%. The Lan-DeMets O'Brien-Fleming approximation spending function is used for calculating efficacy bounds and to preserve the (1-sided) 0.025 false positive error rate over the IAs and the primary analysis (when the target number of cases have been observed), relative to the hypothesis:

$H_0^{efficacy}$: HR ≥ 0.7 (equivalently, proportional hazards VE ≤ 0.3).

There is no intention to stop the study early if the efficacy has been demonstrated at any of the IAs. If efficacy is demonstrated at an IA, the subsequent IA or primary analysis will be considered supportive in nature. The DSMB will review the IA results and make recommendations to the Sponsor in terms of study results reporting and unblinding based on the boundaries of early efficacy as described in this section, safety data, and data external to this study. In addition to possible early efficacy at IAs, the DSMB will monitor for non-efficacy and vaccine harm; the guiding principles (non-binding) is provided in Section 8.4.2, and the details will be provided in the SAP.

Table 15 summarizes the timing, number of cases and decision guidance at each IA and primary analysis.

The first IA will occur when approximately 35% of the total cases have been observed (across both treatment groups). The study will be considered positive at the first IA if the p-value for rejecting HR ≥ 0.7 is less than 0.0002 based on the Lan-DeMets O'Brien-Fleming approximation spending function. This corresponds to an observed HR of approximately 0.259, or an observed VE approximately 0.741.

The second IA will occur when approximately 70% of the total cases have been observed. The study will be considered positive (VE has been demonstrate) if the p-value for rejecting HR ≥ 0.7 is less than 0.0073 based on the Lan-DeMets O'Brien-Fleming approximation spending function. This corresponds to an observed HR of approximately 0.435, or an observed VE of approximately 0.565.

The primary analysis will be performed when approximately 151 cases have been observed in the study. The study will be considered positive at the primary analysis if the 1-sided p-value for rejecting HR ≥ 0.7 is less than 0.0227. This corresponds to an observed HR of approximately 0.505 or observed VE of approximately 0.495.

**Table 15: Interim Boundaries Using O'Brien-Fleming Spending Function, Calculation Based on the PP Set for the Primary Efficacy Endpoint**

| Information Fraction (% of total #cases) | Number of Cases | Nominal Alpha | Efficacy Boundary Rejecting H0: VE ≤ 30% | Cum Prob (crossing efficacy boundary if the true VE = 60%) |
|---|---|---|---|---|
| IA1 35% | 53 | 0.0002 | VE ≥ 0.741<br>(HR ≤ 0.259) | 4.6% |
| IA2 70% | 106 | 0.0073 | VE ≥ 0.565<br>(HR ≤ 0.435) | 61.5% |
| Primary analysis 100% | 151 | 0.0227 | VE ≥ 0.495<br>(HR ≤ 0.505) | 90.0% |

Abbreviations: HR = hazard ratio; IA: interim analysis; LB = lower boundary; PP = per-protocol; VE = vaccine efficacy.

Department of Health and Human Services (DHHS), Food and Drug Administration, Center for Biologics Evaluation and Research (US). Guidance for industry: Toxicity grading scale for healthy adult and adolescent volunteers enrolled in preventative vaccine clinical trials. September 2007 [cited 2019 Apr 10] [10 screens]. Available from:

https://www.fda.gov/downloads/BiologicsBloodVaccines/GuidanceComplianceRegulatoryInformation/Guidances/Vaccines/ucm091977.pdf.

Feldman RA, Fuhr R, Smolenov I, Mick Ribeiro A, Panther L, Watson M, et al. mRNA vaccines against H10N8 and H7N9 influenza viruses of pandemic potential are immunogenic and well tolerated in healthy adults in phase 1 randomized clinical trials. Vaccine. 2019 May 31;37(25):3326-3334.

Fulginiti VA, Eller JJ, Downie AW, Kempe CH. Altered reactivity to measles virus. Atypical measles in children previously immunized with inactivated measles virus vaccines. JAMA. 1967 202:1075-80.

Johnson RF, Bagci U, Keith L, Tang X, Mollura DJ, Zeitlin L, et al. 3B11-N, a monoclonal antibody against MERS-CoV, reduces lung pathology in rhesus monkeys following intratracheal inoculation of MERS-CoV Jordan-n3/2012. Virology. 2016 Mar;490:49-58.

Kim Y, Lee H, Park K, Park S, Lim JH, So MK, et al. Selection and characterization of monoclonal antibodies targeting middle east respiratory syndrome coronavirus through a human synthetic fab phage display library panning. Antibodies (Basel). 2019 Jul 31;8(3).

Rüggeberg JU, Gold MS, Bayas JM, Blum MD, Bonhoeffer J, Friedlander S, et al. Anaphylaxis: case definition and guidelines for data collection, analysis, and presentation of immunization safety data. Vaccine. 2007;25(31):5675-84.

Thomas SJ and Yoon IK. A review of Dengvaxia®: development to deployment. Hum Vaccin Immunother. 2019;15(10), 2295-2314.

Wang L, Shi W, Joyce MG, Modjarrad K, Zhang Y, Leung K, et al. Evaluation of candidate vaccine approaches for MERS-CoV. Nat Commun. 2015 Jul 28;6:7712.

Wang L, Shi W, Chappell JD, Joyce MG, Zhang Y, Kanekiyo M, et al. Importance of neutralizing monoclonal antibodies targeting multiple antigenic sites on the middle east respiratory syndrome coronavirus spike glycoprotein to avoid neutralization escape. J Virol. 2018 Apr 27;92(10).

Widjaja I, Wang C, van Haperen R, Gutiérrez-Álvarez J, van Dieren B, Okba NMA, et al. Towards a solution to MERS: protective human monoclonal antibodies targeting different domains and functions of the MERS-coronavirus spike glycoprotein. Emerg Microbes Infect. 2019;8(1):516-30.

World Health Organization (WHO). Dengue vaccine: WHO position paper, September 2018 – recommendations. Vaccine. 2019 Aug;37(35):4848-4849

World Health Organization (WHO). 2020 (Data reported as of 2020, May 28). Coronavirus disease 2019 (COVID-19) Situation Report – 129. Retrieved from: https://www.who.int/docs/default-source/coronaviruse/situation-reports/20200528-covid-19-sitrep-129.pdf?sfvrsn=5b154880_2. (accessed 2020 May 29).

Wrapp D, Wang N, Corbett KS, Goldsmith JA, Hseih CL, Abiona O, et al. Cryo-EM structure of the 2019-nCoV spike in the prefusion conformation. Science. 2020;367:1260-3.

Yu X, Zhang S, Jiang L, Cui Y, Li D, Wang D, et al. Structural basis for the neutralization of MERS-CoV by a human monoclonal antibody MERS-27. Sci Rep. 2015 Aug 18;5:13133.

Zeng D. 2004. Estimating marginal survival function by adjusting for dependent censoring using many covariates. The Annals of Statistics, 32(4), 1533-1555.

Zent O, Arras-Reiter C, Broeker M, Hennig R. Immediate allergic reactions after vaccinations – a post-marketing surveillance review. Eur J Pediatr. 2002;161(1):21-25.

### 11. SUPPORTING DOCUMENTATION AND OPERATIONAL CONSIDERATIONS

#### 11.1. APPENDIX 1: Schedules of Events

If a participant cannot attend a study site visit (scheduled or unscheduled, with the exception of Screening, Day 1, Day 29, Participant Decision clinic visit, OL-D1, and OL-D29), a home visit is acceptable if performed by appropriately delegated study site staff or a home healthcare service provided by the Sponsor. If neither a participant visit to the study site nor a home visit to the participant is possible (with the exception of Screening, Day 1, Day 29, Participant Decision clinic visit, OL-D1, and OL-D29), a safety phone call should be performed that includes the assessments scheduled for the biweekly safety phone calls (Table 16).

The Supplemental SoE (Table 21) and the Modified Supplemental SoE (Table 22) are intended to occur in addition to the original Schedules of Events, as applicable (Table 16, Table 17, Table 18, and Table 19) and therefore there is a possibility for study visits to overlap. If visits overlap according to respective visit windows, a single visit may be done with the combined study procedures completed once.

**Table 16: Schedule of Events (Vaccination Phase, Day 1 – Day 57)**

| Visit Number | 0 | 1 | | | | 2 | | | | 3 |
|---|---|---|---|---|---|---|---|---|---|---|
| Type of Visit | C | C | SC | SC | SC | C | SC | SC | SC | C |
| Month/Weekly Timepoint | | M0 | | | | M1 | | | | M2 |
| **Study Visit Day** | D0[1] (Screening) | D1 (Baseline) | 7 days after D1 D8 | 14 days after D1 D15 | 21 days after D1 D22 | D29 | 7 days after D29 D36 | 14 days after D29 D43 | 21 days after D29 D50 | D57 |
| **Window Allowance (Days)** | -28 | | +3 | +3 | +3 | -3/+7 | +3 | +3 | +3 | -3/+7 |
| **Days Since Most Recent Vaccination** | - | 0 | 7 | 14 | 21 | 28/0 | 7 | 14 | 21 | 28 |
| ICF, demographics, concomitant medications, medical history | X | | | | | | | | | |
| Confirm participant meets inclusion and exclusion criteria | X | X | | | | | | | | |
| Physical examination[2] | X | X | | | | X | | | | X |
| Pregnancy testing[3] | X | X | | | | X | | | | |
| Randomization | | X | | | | | | | | |
| **Dosing** | | | | | | | | | | |
| Study injection (including 30-minute post-dosing observation period) | | X | | | | X | | | | |
| **Efficacy Assessment** | | | | | | | | | | |
| Surveillance for COVID-19/Unscheduled Visit[4] | | X | X | X | X | X | X | X | X | X |
| Nasopharyngeal swab[5] | | X | | | | X | | | | |
| **Immunogenicity Assessment** | | | | | | | | | | |
| Blood for immunologic analysis[5] | | X | | | | X | | | | X |

| **Visit Number** | 0 | 1 | | | | 2 | | | | 3 |
|---|---|---|---|---|---|---|---|---|---|---|
| **Type of Visit** | C | C | SC | SC | SC | C | SC | SC | SC | C |
| **Month/Weekly Timepoint** | | M0 | | | | M1 | | | | M2 |
| **Study Visit Day** | D0[1] (Screening) | D1 (Baseline) | 7 days after D1 D8 | 14 days after D1 D15 | 21 days after D1 D22 | D29 | 7 days after D29 D36 | 14 days after D29 D43 | 21 days after D29 D50 | D57 |
| **Window Allowance (Days)** | -28 | | +3 | +3 | +3 | -3/+7 | +3 | +3 | +3 | -3/+7 |
| **Days Since Most Recent Vaccination** | - | 0 | 7 | 14 | 21 | 28/0 | 7 | 14 | 21 | 28 |
| **Safety Assessments** | | | | | | | | | | |
| eDiary activation for recording solicited adverse reactions (7 days)[6] | | X | | | | X | | | | |
| Review of eDiary | | | X | | | | X | | | |
| Follow-up Safety[7] | | | X | X | X | | X | X | X | |
| Recording of Unsolicited AEs (Blinded Phase only) | | X | X | X | X | X | X | X | X | X |
| Recording of MAAEs, AE leading to withdrawal and concomitant medications relevant to or for the treatment of the MAAE[8] | | X | X | X | X | X | X | X | X | X |
| Recording of SAEs and concomitant medications relevant to or for the treatment of the SAE[8] | X | X | X | X | X | X | X | X | X | X |
| Recording of concomitant medications and non-study vaccinations[8] | | X | X | X | X | X | X | X | X | X |

Abbreviations: AE = adverse event; C = clinic visit; D = day; eDiary= electronic diary; ICF = informed consent form; M = month; MAAE = medically attended AE; SAE = serious adverse event; SC = safety (phone) call.

Note: In accordance with FDA Guidance on Conduct of Clinical Trials of Medical Products during the COVID-19 Pandemic (FDA March 2020), investigators may convert study site visits to telemedicine visits with the approval of the Sponsor. All scheduled study visits should be completed within the respective visit windows. If the participant is not able to come on site for a study site visit as a result of the COVID-19 Pandemic (self-quarantine or disruption of study site activities following business continuity plans and/or local government mandates for “stay at home” or “shelter in place”), a safety call to the participant should be made in place of the study site visit. The safety call should encompass all scheduled visit assessments that can be completed remotely, such as assessment for AEs and concomitant medications (eg, as defined in scheduled biweekly safety phone calls). Home visits will be permitted for all non-dosing visits except for Screening if a participant cannot come to the study site as a result of the COVID19 pandemic.

1. Day 0 and Day 1 may be combined the same day. Additionally, the Day 0 visit may be performed over multiple visits if within the 28-day screening window.
2. Physical examination: a full physical examination, including vital signs, height, and weight, will be performed at Screening and Day 1. Body mass index will be calculated at the Screening Visit (Day 0) only. Symptom-directed physical examinations will be performed on Day 29 and Day 57. On each dosing day before injection, the arm receiving the injection should be examined and the associated lymph nodes should be evaluated. Any clinically significant finding identified during a study visit should be reported as a MAAE. Vital signs are to be collected pre- and post-dosing on days of injection (Day 1 and Day 29). When applicable, vital sign measurements should be performed before blood collection. Participants who are febrile (body temperature ≥ 38.0°C/100.4°F) before dosing on Day 1 or Day 29 must be rescheduled within the relevant window period to receive the injection. Afebrile participants with minor illnesses can be enrolled at the discretion of the investigator.
3. Pregnancy test at Screening and Day 1 and before the second vaccination will be a point-of-care urine test. At the discretion of the investigator a pregnancy test either via blood or point-of-care urine test can be performed. Follicle-stimulating hormone level may be measured to confirm menopausal status at the discretion of the investigator.
4. Participants with symptoms of COVID-19 will be asked to return within 72 hours or as soon as possible to the study site for an unscheduled visit, to include an NP swab sample (for RT-PCR testing) and other clinical evaluations. If a study site visit is not possible, a home visit may be arranged to collect the NP sample and conduct clinical evaluations. If a home visit is not possible, the participant will be asked to submit a saliva sample to the study site by a Sponsor-approved method. At this visit, NP swab and blood samples will be collected to evaluate for the presence of SARS-CoV-2 infection. NP swab samples will also be tested for the presence of other respiratory pathogens. In addition, the study site may collect an additional respiratory sample for SARS-CoV-2 testing to be able to render appropriate medical care for the study participant as determined by local standards of care. Additionally, clinical information will be carefully collected to evaluate the severity of the clinical case. It is important to note that some of the symptoms of COVID-19 overlap with solicited systemic ARs that are expected after vaccination with mRNA-1273 (eg, myalgia, headache, fever, and chills). During the first 7 days after vaccination, when these solicited ARs are common, investigators should use their clinical judgement to decide if an NP swab should be collected. The collection of an NP swab prior to the Day 1 and Day 29 vaccination can help ensure that cases of COVID-19 are not overlooked. Any study participant reporting respiratory symptoms during the 7-day period after vaccination should be evaluated for COVID-19.
5. Sample must be collected prior to dosing on days of injection (Day 1 and Day 29).
6. The participant will record entries in the eDiary approximately 30 minutes after dosing while at the study site, with instruction provided by study staff. Study participants will continue to record in the eDiary each day after they leave the study site, preferably in the evening, on the day of dosing and for 6 days following. Any solicited AR that is ongoing beyond Day 7 will be reported until resolution. Adverse reactions recorded in eDiaries beyond Day 7 should be reviewed either via phone call or at the following study visit. Participants will be given thermometers to record their temperatures and rulers to measure any injection site reactions.
7. Trained study personnel will call all participants to collect information relating to any unsolicited AEs through Day 57 (including any signs and symptoms of COVID-19), MAAEs, AEs leading to withdrawal, SAEs, information on concomitant medications associated with those events, and any non-study vaccinations.
8. All concomitant medications and non-study vaccinations will be recorded through 28 days after each injection; all concomitant medications relevant to or for the treatment of an SAE or MAAE will be recorded from Screening through the final visit (Day 759).

**Table 17: Schedule of Events (Surveillance Phase, Day 64 – Day 394)**

| **Visit Number** | | | | | | 4 | | | | | | 5 |
|---|---|---|---|---|---|---|---|---|---|---|---|---|
| **Type of Visit** | eD | SC | SC | SC | SC | C | SC | SC | SC | SC | SC | C |
| **Month (M)/Weekly (W) Timepoint** | W | M3 | M4 | M5 | M6 | M7 | M8 | M9 | M10 | M11 | M12 | M13 |
| **Study Visit Day** | D64 [1] | D85[1] | D119 [1] 90 days after D29 | D149[1] | D179[1] | D209[1] 180 days after D29 | D239[1] | D269[1] | D299[1] | D329[1] | D359[1] | D394[1] 365 days after D29 |
| **Window Allowance (Days)** | ±2 | ±3 | ±3 | ±3 | ±3 | ±14 | ±3 | ±3 | ±3 | ±3 | ±3 | ±14 |
| **Days Since Most Recent Vaccination** | - | 56 | 90 | 120 | 150 | 180 | 210 | 240 | 270 | 300 | 330 | 365 |
| Physical examination[2] | | | | | | X | | | | | | X |
| **Efficacy Assessments** | | | | | | | | | | | | |
| eDiary activation for surveillance for COVID-19/Unscheduled Visit | X | | | | | | | | | | | |
| eDiary Weekly Prompts for surveillance for COVID-19 | -------------------Weekly eDiary Prompts (Day 64 through Day 387) ----------- |  |  |  |  |  |  |  |  |  |  |  |
| Surveillance for COVID-19/Unscheduled Visit[3] | | X | X | X | X | X | X | X | X | X | X | X |
| **Immunogenicity Assessment** | | | | | | | | | | | | |
| Blood for immunologic analysis | | | | | | X | | | | | | X |
| **Safety Assessments** | | | | | | | | | | | | |
| eDiary activation for safety follow-up | X[4] | | | | | | | | | | | |
| eDiary Weekly Prompts for safety follow-up | --------------Weekly eDiary Prompts (Day 64 through Day 387) ---------------- |  |  |  |  |  |  |  |  |  |  |  |

| Visit Number | | | | | | 4 | | | | | | 5 |
|---|---|---|---|---|---|---|---|---|---|---|---|---|
| Type of Visit | eD | SC | SC | SC | SC | C | SC | SC | SC | SC | SC | C |
| Month (M)/Weekly (W) Timepoint | W | M3 | M4 | M5 | M6 | M7 | M8 | M9 | M10 | M11 | M12 | M13 |
| **Study Visit Day** | D64 [1] | D85[1] | D119 [1] 90 days after D29 | D149[1] | D179[1] | D209[1] 180 days after D29 | D239[1] | D269[1] | D299[1] | D329[1] | D359[1] | D394[1] 365 days after D29 |
| **Window Allowance (Days)** | ±2 | ±3 | ±3 | ±3 | ±3 | ±14 | ±3 | ±3 | ±3 | ±3 | ±3 | ±14 |
| **Days Since Most Recent Vaccination** | - | 56 | 90 | 120 | 150 | 180 | 210 | 240 | 270 | 300 | 330 | 365 |
| Follow-up Safety[5] | | X | X | X | X | X | X | X | X | X | X | X |
| Recording of MAAEs, AE leading to withdrawal and concomitant medications relevant to or for the treatment of the MAAE[6] | | X | X | X | X | X | X | X | X | X | X | X |
| Recording of SAEs and concomitant medications relevant to or for the treatment of the SAE[6] | | X | X | X | X | X | X | X | X | X | X | X |
| Recording of concomitant medications and non-study vaccinations[6,7] | | X | X | X | X | X | X | X | X | X | X | X |

Abbreviations: AE = adverse event; C = clinic visit; D = day; eD = electronic diary; IRB = institutional review board; M = month; MAAE = medically attended AE; SAE = serious adverse event; SC = safety (phone) call; W = week.

Note: In accordance with FDA Guidance on Conduct of Clinical Trials of Medical Products during the COVID-19 Pandemic (FDA March 2020), investigators may convert study site visits to telemedicine visits with the approval of the Sponsor.

1. All scheduled study visits should be completed within the respective visit windows. If the participant is not able to come on site for a study site visit as a result of the COVID-19 Pandemic (self-quarantine or disruption of study site activities following business continuity plans and/or local government mandates for "stay at home" or "shelter in place"), a safety phone call to the participant should be made in place of the study site visit. The safety call should encompass all scheduled visit assessments that can be completed remotely, such as assessment for AEs and concomitant medications (eg, as defined in scheduled safety phone calls). Home visits will be permitted for all non-dosing visits except for Screening if a participant cannot come to the study site as a result of the COVID-19 pandemic. Home visits must be permitted by the site IRB and the participant via informed consent and have prior approval from the Sponsor (or its designee).
2. Symptom-directed physical examinations may be performed at the discretion of the investigator.

3. Participants with symptoms of COVID-19 will be asked to return within 72 hours or as soon as possible to the study site for an unscheduled visit, to include an NP swab sample (for RT-PCR testing) and other clinical evaluations. If a study site visit is not possible, a home visit may be arranged to collect the NP sample and conduct clinical evaluations. If a home visit is not possible, the participant will be asked to submit a saliva sample to the study site by a Sponsor-approved method. At this visit, NP swab and blood samples will be collected to evaluate for the presence of SARS-CoV-2 infection. NP swab samples will also be tested for the presence of other respiratory pathogens. In addition, the study site may collect an additional respiratory sample for SARS-CoV-2 testing to be able to render appropriate medical care for the study participant as determined by local standards of care. Additionally, clinical information will be carefully collected to evaluate the severity of the clinical case.
4. The eDiary Safety Follow-up Prompts will be triggering off of Day 61 to take into consideration the (-3 day) window allowance at Day 29. This first Safety Follow-Up prompt at D61 will follow the same 7-day surveillance per protocol.
5. Trained study personnel will call all participants to collect information relating to any sign/symptoms of COVID-19, MAAEs, AEs leading to withdrawal, SAEs, information on concomitant medications associated with those events, and any non-study vaccinations.
6. All concomitant medications relevant to or for the treatment of an SAE or MAAE will be recorded from Screening through the final visit (Day 759).
7. One additional ePRO prompt will be sent to participants specifically to solicit the collection of information regarding participant's history of facial injections or dermal fillers, for cosmetic or medical indications such as migraine headaches.

**Table 18: Schedule of Events (Surveillance Phase, Day 401 – Day 759)**

| **Visit Number** | | | | | | | | | | | | 6 |
|---|---|---|---|---|---|---|---|---|---|---|---|---|
| **Type of Visit** | SC | SC | SC | SC | SC | SC | SC | SC | SC | SC | SC | C |
| **Month (M)/Weekly (W) Timepoint** | M14 | M15 | M16 | M17 | M18 | M19 | M20 | M21 | M22 | M23 | M24 | M25 |
| **Study Visit Day** | D424[1] | D454[1] | D484[1] | D514[1] | D544[1] | D574[1] | D604[1] | D634[1] | D664[1] | D694[1] | D724[1] | D759[1] 365 days after Year 1 |
| **Window Allowance (Days)** | ±3 | ±3 | ±3 | ±3 | ±3 | ±3 | ±3 | ±3 | ±3 | ±3 | ±3 | ±14 |
| **Days Since Most Recent Vaccination** | | | | | | | | | | | | 730 |
| Physical examination[2] | | | | | | | | | | | | X |
| eDiary Weekly Prompts for surveillance for COVID-19 |  | □--------Weekly eDiary Prompts (Day 401 through Day 752) --------------□ |  |  |  |  |  |  |  |  |  |  |
| Surveillance for COVID-19/Unscheduled Visit[3] | X | X | X | X | X | X | X | X | X | X | X | X |
| Blood for immunologic analysis | | | | | | | | | | | | X |
| eDiary Weekly Prompts for safety follow-up |  | □--------Weekly eDiary Prompts (Day 401 through Day 752) --------------□ |  |  |  |  |  |  |  |  |  |  |
| Follow-up Safety[4] | X | X | X | X | X | X | X | X | X | X | X | X |
| Recording of MAAEs, AE leading to withdrawal and concomitant medications relevant to or for the treatment of the MAAE[5] | X | X | X | X | X | X | X | X | X | X | X | X |
| Recording of SAEs and concomitant medications relevant to or for the treatment of the SAE[5] | X | X | X | X | X | X | X | X | X | X | X | X |
| Recording of concomitant medications and non-study vaccinations[5] | X | X | X | X | X | X | X | X | X | X | X | X |

Abbreviations: AE = adverse event; C = clinic visit; D = day; eD = electronic diary; IRB = institutional review board; M = month; MAAE = medically attended AE; SAE = serious adverse event; SC = safety (phone) call.

Note: In accordance with FDA Guidance on Conduct of Clinical Trials of Medical Products during the COVID-19 Pandemic (FDA March 2020), investigators may convert study site visits to telemedicine visits with the approval of the Sponsor.

1. All scheduled study visits should be completed within the respective visit windows. If the participant is not able to come on site for a study site visit as a result of the COVID-19 Pandemic (self-quarantine or disruption of study site activities following business continuity plans and/or local government mandates for "stay at home" or "shelter in place"), a safety phone call to the participant should be made in place of the study site visit. The safety phone call should encompass all scheduled visit assessments that can be completed remotely, such as assessment for AEs and concomitant medications (eg, as defined in scheduled safety phone calls). Home visits will be permitted for all non-dosing visits except for Screening if a participant cannot come to the study site as a result of the COVID-19 pandemic. Home visits must be permitted by the site IRB and the participant via informed consent and have prior approval from the Sponsor (or its designee).
2. Symptom-directed physical examinations may be performed at the discretion of the investigator.
3. Participants with symptoms of COVID-19 will be asked to return within 72 hours or as soon as possible to the study site for an unscheduled visit, to include an NP swab sample (for RT-PCR testing) and other clinical evaluations. If a study site visit is not possible, a home visit may be arranged to collect the NP sample and conduct clinical evaluations. If a home visit is not possible, the participant will be asked to submit a saliva sample to the study site by a sponsor-approved method. At this visit, NP swab and blood samples will be collected to evaluate for the presence of SARS-CoV-2 infection. NP swabs will also be tested for the presence of other respiratory pathogens. In addition, the study site may collect an additional NP sample for SARS-CoV-2 testing to be able to render appropriate medical care for the study participant as determined by local standards of care. Additionally, clinical information will be carefully collected to evaluate the severity of the clinical case.
4. Trained study personnel will call all participants to collect information relating to any MAAEs, AEs leading to withdrawal, SAEs, information on concomitant medications associated with those events, and any non-study vaccinations.
5. All concomitant medications relevant to or for the treatment of an SAE or MAAE will be recorded from Screening through the final visit (Day 759).

**Table 19: Schedule of Events (Convalescent Period, Starting with the Illness Visit)**

| Unscheduled Visit | 1 | | | | | | | | 2 |
|---|---|---|---|---|---|---|---|---|---|
| **Type of Visit** | C/H | | | | | | | | C/H |
| **Daily Timepoint** | D1 | D2- D6 | D7 | D8-D13 | D14 | D15-D20 | D21 | D22-D27 | D28 |
| **Window Allowance (Days)** | - | ±1 | ±1 | ±1 | ±1 | ±1 | ±1 | ±1 | +7 |
| **Safety Assessments** | | | | | | | | | |
| Symptom-directed Physical examination[1] | X | | | | | | | | X |
| Follow-up Safety[2] | X | | | | | | | | X |
| Recording of MAAEs, AE leading to withdrawal and concomitant medications relevant to or for the treatment of the MAAE[3] | X | | | | | | | | X |
| Recording of SAEs and concomitant medications relevant to or for the treatment of the SAE[3] | X | | | | | | | | X |
| Recording of concomitant medications and non-study vaccinations[3] | X | | | | | | | | X |
| **Efficacy Assessments** | | | | | | | | | |
| Daily Telemedicine Visit[4] |  | ------------------------------------------Daily------------------------- |  |  |  |  |  |  |  |
| Respiratory illness sample[5] | X | | | | | | | | |
| Blood sample for immunologic assessment of SARS-CoV-2 infection[6] | X | | | | | | | | X |
| Saliva sample | | D3[7], D5[7] | D7[7] | D9[7] | D14[7] | | D21[7] | | X[8] |

Abbreviations: C = clinic visit; CLIA = Clinical Laboratory Improvement Amendments; D = day; H = home visit; MAAE = medically attended AE; SAE = serious adverse event

Note: In accordance with FDA Guidance on Conduct of Clinical Trials of Medical Products during the COVID-19 Pandemic (FDA March 2020), investigators may convert study site visits to telemedicine visits with the approval of the Sponsor.

1. Physical examination: a symptom-directed physical examination, including vital signs will be performed at the initial visit to confirm the diagnosis (denoted as D1 in this table) and at the Convalescent Visit (28 days after diagnosis [D28 in this table]).
2. Trained study personnel will call all participants to collect information relating to any AEs, MAAEs, AEs leading to withdrawal, SAEs, information on concomitant medications associated with those events, and any non-study vaccinations. All safety events will be followed until resolution.

3. All concomitant medications relevant to or for the treatment of an SAE or MAAE will be recorded from Screening through the final visit (Day 759).
4. Participants will have daily telemedicine visits (via video or phone) for 14 days (or until symptoms resolve, whichever is longer). Telemedicine visits may be performed by medically qualified staff appropriately delegated by the investigator. During the telemedicine visit (preferably done in the evening) the participant will be asked to verbally report the severity of each symptom, their highest body temperature and lowest oxygen saturation for that day and the investigator will determine if medical attention is required due to worsening of COVID-19. The participant will also be reminded to collect a saliva sample (or nasal swab) and return it to the study site, on the appropriate days.
5. Participants with symptoms of COVID-19 will be asked to return within 72 hours or as soon as possible to the study site or medically qualified staff from the site will conduct a home visit as soon as possible to collect an NP swab sample (for RT-PCR) and collect a blood sample for immunologic assessment of SARS-CoV-2 infection and evaluate for COVID-19. NP swab sample will also be tested for the presence of other respiratory infections. In addition, the study site may collect an additional respiratory sample for SARS-CoV-2 testing to be able to render appropriate medical care for the study participant as determined by local standards of care. Additionally, clinical information will be carefully collected to evaluate the severity of the clinical case. If the RT-PCR test from the NP swab sample from the Illness Visit is negative for SARS-CoV-2, the participant will exit the Convalescent Period and resume the study schedule (Table 16, Table 17, and Table 18).
6. This can be a home visit if necessary.
7. Participants will collect their own saliva sample using the saliva collection tubes provided, or a nasal swab, on 3, 5, 7, 9, 14, and 21 days after the initial Illness Visit, and return them to the study site according to Sponsor instructions. Participants who are pending a central laboratory PCR may exit from the convalescent period based on a negative PCR result from a CLIA-certified or CLIA-certified waiver local laboratory at the investigator’s discretion.
8. At this visit, a saliva (or nasal swab) sample will be collected.

**Table 20: Part B: Participant Decision Clinic Visit**

| | **All Participants** | | | | |
|---|---|---|---|---|---|
| Sign revised Informed Consent Form | X | | | | |
| Confirm participant's request to be unblinded or not to be unblinded[a] | X | | | | |
| Nasopharyngeal swab | X | | | | |
| Blood for immunologic analysis | X | | | | |
| Counselling the importance of public health measures[b] | X | | | | |
| | | **Open-label Cohort** | | | |
| **Participant Status after Participant Decision Clinic Visit** | **Blinded Cohort** | **Previously Receiving mRNA-1273** | **Remaining on Placebo** | **Placebo Requesting mRNA-1273** | **mRNA-1273 who received 1 dose ONLY** |
| Continue with original Schedules of Events, as applicable (Table 16, Table 17, Table 18, and Table 19) | X | X | X | X | X |
| Supplemental Schedule of Events: Open-Label Days 1-57 (Table 21)[c] | -- | -- | -- | X | -- |
| Modified Supplemental Schedule of Events: Open-Label Days 1-57 (Table 22)[c] | -- | -- | -- | -- | X |

[a] At the point when the mRNA-1273 vaccine is no longer available for study use, any participant who schedules a Participant Decision Visit after this time-point will be unblinded (with no option to stay blinded), but will not be offered m-RNA-1273 study vaccine.

[b] All participants are counselled about the importance of continuing other public health measures to limit the spread of disease including physical-social distancing, wearing a mask, and hand-washing.

[c] The Supplemental and the Modified Supplemental Schedule of Events are intended to occur in addition to the original Schedules of Events, as applicable (Table 16, Table 17, Table 18, and Table 19) and therefore there is a possibility for study visits to overlap. If visits overlap according to respective visit windows, a single visit may be done with the combined study procedures completed once.

#### Table 21: Supplemental Schedule of Events: Open-Label Observational Phase - Placebo Participants who Request to Receive mRNA-1273

**NOTE: Supplemental Schedule of Events: Open-Label Observational Phase is to be used ONLY for participants who received placebo during the blinded phase of this study (Part A), and request to receive mRNA-1273. These participants will comply with this Supplemental SoE (Table 21) in addition to the original SoEs as applicable (Table 16, Table 17, Table 18, and Table 19). As this Supplemental SoE is intended to occur in addition to the original Schedules of Events, there is a possibility for study visits to overlap. If visits overlap according to respective visit windows, a single visit may be done with the combined study procedures completed once.**

| **Visit Number** | 0 | OL-1 | | OL-2 | | OL-3 |
|---|---|---|---|---|---|---|
| **Type of Visit** | C | C | SC | C | SC | C |
| **Study Visit Day** | Participant Decision Visit[1] and OL-D1[1] | | 7 days after OL-D1 OL-D8 | OL-D29 | 7 days after OL-D29 OL-D36 | OL-D57 |
| **Window Allowance (Days)** | ? | | +3 | -3/+14 | +3 | -3/+14 |
| **Days Since Most Recent Vaccination** | 0 | | 7 | 28/0 | 7 | 28 |
| Confirm signing of ICF, concomitant medications, medical history | X | | | | | |
| Confirm participant meets inclusion and exclusion criteria | X | | | | | |
| Physical examination[2] | X | | | X | | X |
| Pregnancy testing[3] | X | | | X | | |
| **Immunogenicity Assessment** | | | | | | |
| Blood for immunologic analysis[5] | X | | | | | X |
| **Dosing** | | | | | | |
| Study injection (including 30-minute post-dosing observation period[6]) | X | | | X | | |
| **Efficacy Assessment** | | | | | | |

**NOTE: Supplemental Schedule of Events: Open-Label Observational Phase is to be used ONLY for participants who received placebo during the blinded phase of this study (Part A), and request to receive mRNA-1273. These participants will comply with this Supplemental SoE (Table 21) in addition to the original SoEs as applicable (Table 16, Table 17, Table 18, and Table 19). As this Supplemental SoE is intended to occur in addition to the original Schedules of Events, there is a possibility for study visits to overlap. If visits overlap according to respective visit windows, a single visit may be done with the combined study procedures completed once.**

| **Visit Number** | 0 | OL-1 | | OL-2 | | OL-3 |
|---|---|---|---|---|---|---|
| **Type of Visit** | C | C | SC | C | SC | C |
| **Study Visit Day** | Participant Decision Visit[1] and OL-D1[1] | | 7 days after OL-D1 OL-D8 | OL-D29 | 7 days after OL-D29 OL-D36 | OL-D57 |
| **Window Allowance (Days)** | ? | | +3 | -3/+14 | +3 | -3/+14 |
| **Days Since Most Recent Vaccination** | 0 | | 7 | 28/0 | 7 | 28 |
| Surveillance for COVID-19/Unscheduled Visit[4] | X | | X | X | X | X |
| Nasopharyngeal swab[5] | X | | | | | |
| **Safety Assessments** | | | | | | |
| Follow-up Safety[7] | | | X | | X | |
| Recording of MAAEs, AE leading to withdrawal and concomitant medications relevant to or for the treatment of the MAAE[8] | X | | X | X | X | X |
| Recording of SAEs and concomitant medications relevant to or for the treatment of the SAE[8] | X | | X | X | X | X |
| Recording of concomitant medications and non-study vaccinations[8] | X | | X | X | X | X |

Abbreviations: AE = adverse event; C = clinic visit; D = day; eDiary = electronic diary; ICF = informed consent form; OL = open-label; MAAE = medically attended AE; SAE = serious adverse event; SC = safety (phone) call.

Note: In accordance with FDA Guidance on Conduct of Clinical Trials of Medical Products during the COVID-19 Pandemic (FDA March 2020), investigators may convert study site visits to telemedicine visits with the approval of the Sponsor. All scheduled study visits should be completed within the respective visit windows. If the participant is not able to come on site for a study site visit as a result of the COVID-19 Pandemic (self-quarantine or disruption of study site activities following business continuity plans and/or local government mandates for "stay at home" or "shelter in place"), a safety call to the participant should be made in place of the study site visit. The safety call should encompass all scheduled visit assessments that can be completed remotely, such as assessment for AEs and concomitant medications (eg, as defined in scheduled biweekly safety phone calls). Home visits will be permitted for all non-dosing visits except for Screening if a participant cannot come to the study site as a result of the COVID19 pandemic.

1. Day 0 and OL-D1 visit may be performed on 2 separate visits.
2. Symptom-directed physical examination will be performed at the Participant Decision Visit and on OL-D29. On each dosing day before injection, the arm receiving the injection should be examined and the associated lymph nodes should be evaluated. Any clinically significant finding identified during a study visit should be reported as a MAAE. Vital signs are to be collected pre- and post-dosing on days of injection (OL-D1 and OL-D29). Participants who are febrile (body temperature ≥ 38.0°C/100.4°F) before dosing (OL-D1 and OL-D29) must be rescheduled within the relevant window period to receive the injection. Afebrile participants with minor illnesses can be enrolled at the discretion of the investigator.
3. Pregnancy test at the Participant Decision Visit and OL-D29 will be a point-of-care urine test. At the discretion of the investigator, a pregnancy test either via blood or point-of-care urine test can be performed. Follicle-stimulating hormone level may be measured to confirm menopausal status at the discretion of the investigator.
4. Participants with symptoms of COVID-19 will be asked to return within 72 hours or as soon as possible to the study site for an unscheduled visit, to include an NP swab sample (for RT-PCR testing) and other clinical evaluations. If a study site visit is not possible, a home visit may be arranged to collect the NP sample and conduct clinical evaluations. If a home visit is not possible, the participant will be asked to submit a saliva sample to the study site by a Sponsor-approved method. At this visit, NP swab and blood samples will be collected to evaluate for the presence of SARS-CoV-2 infection. NP swab samples will also be tested for the presence of other respiratory pathogens. In addition, the study site may collect an additional respiratory sample for SARS-CoV-2 testing to be able to render appropriate medical care for the study participant as determined by local standards of care. Additionally, clinical information will be carefully collected to evaluate the severity of the clinical case. It is important to note that some of the symptoms of COVID-19 overlap with solicited systemic ARs that are expected after vaccination with mRNA-1273 (eg, myalgia, headache, fever, and chills). During the first 7 days after vaccination, when these solicited ARs are common, investigators should use their clinical judgement to decide if an NP swab should be collected. The collection of an NP swab prior to the Participant Decision Visit can help ensure that cases of COVID-19 are not overlooked. Any study participant reporting respiratory symptoms during the 7-day period after vaccination should be evaluated for COVID-19.
5. Sample must be collected prior to unblinding and injection at the Participant Decision Visit.
6. Post-dosing, participants will have a 30-minute observation period.
7. Trained study personnel will call all participants to collect information relating to any MAAEs (including any signs and symptoms of COVID-19), AEs leading to withdrawal, SAEs, information on concomitant medications associated with those events, and any non-study vaccinations.
8. All concomitant medications relevant to or for the treatment of an SAE or MAAE will be recorded from Screening through the final visit (Day 759).

**Table 22: Modified Supplemental Schedule of Events: Open-Label Observational Phase – mRNA-1273 Participants who ONLY received 1 dose of mRNA-1273**

**NOTE: Modified Supplemental Schedule of Events: Open-Label Observational Phase is to be used ONLY for participants who ONLY received 1 dose of mRNA-1273 during the blinded phase of this study (Part A), and consented to receive mRNA-1273 (see Section 4.1.2 for details). These participants will comply with this SoE (Table 22) in addition to the original SoEs as applicable (Table 16, Table 17, Table 18, and Table 19). As this Modified Supplemental SoE is intended to occur in addition to the original Schedules of Events, there is a possibility for study visits to overlap. If visits overlap according to respective visit windows, a single visit may be done with the combined study procedures completed once.**

| **Visit Number** | 0 | OL-1 | | OL-2 |
|---|---|---|---|---|
| **Type of Visit** | C | C | SC | C |
| **Study Visit Day** | Participant Decision Visit[1] and OL-D1[1] | | 7 days after OL-D1 OL-D8 | OL-D29 |
| **Window Allowance (Days)** | - | | +3 | -3/+14 |
| **Days Since Most Recent Vaccination** | 0 | | 7 | 28/0 |
| Confirm signing of ICF, concomitant medications, medical history | X | | | |
| Confirm participant meets inclusion and exclusion criteria | X | | | |
| Physical examination[2] | X | | | X |
| Pregnancy testing[3] | X | | | |
| **Immunogenicity Assessment** | | | | |
| Blood for immunologic analysis[5] | X | | | X |
| **Dosing** | | | | |
| Study injection (including 30-minute post-dosing observation period[6]) | X | | | |

**NOTE: Modified Supplemental Schedule of Events: Open-Label Observational Phase is to be used ONLY for participants who ONLY received 1 dose of mRNA-1273 during the blinded phase of this study (Part A), and consented to receive mRNA-1273 (see Section 4.1.2 for details). These participants will comply with this SoE (Table 22) in addition to the original SoEs as applicable (Table 16, Table 17, Table 18, and Table 19). As this Modified Supplemental SoE is intended to occur in addition to the original Schedules of Events, there is a possibility for study visits to overlap. If visits overlap according to respective visit windows, a single visit may be done with the combined study procedures completed once.**

| **Visit Number** | 0 | OL-1 | | OL-2 |
|---|---|---|---|---|
| **Type of Visit** | C | C | SC | C |
| **Study Visit Day** | Participant Decision Visit[1] and OL-D1[1] | | 7 days after OL-D1 OL-D8 | OL-D29 |
| **Window Allowance (Days)** | - | | +3 | -3/+14 |
| **Days Since Most Recent Vaccination** | 0 | | 7 | 28/0 |
| **Efficacy Assessment** | | | | |
| Surveillance for COVID-19/Unscheduled Visit[4] | X | | X | X |
| Nasopharyngeal swab[5] | X | | | |
| **Safety Assessments** | | | | |
| Follow-up Safety[7] | | | X | |
| Recording of MAAEs, AE leading to withdrawal and concomitant medications relevant to or for the treatment of the MAAE[8] | X | | X | X |
| Recording of SAEs and concomitant medications relevant to or for the treatment of the SAE[8] | X | | X | X |
| Recording of concomitant medications and non-study vaccinations[8] | X | | X | X |

Abbreviations: AE = adverse event; C = clinic visit; D = day; eDiary = electronic diary; ICF = informed consent form; OL = open-label; MAAE = medically attended AE; SAE = serious adverse event; SC = safety (phone) call.

Note: In accordance with FDA Guidance on Conduct of Clinical Trials of Medical Products during the COVID-19 Pandemic (FDA March 2020), investigators may convert study site visits to telemedicine visits with the approval of the Sponsor. All scheduled study visits should be completed within the respective visit windows. If the participant is not able to come on site for a study site visit as a result of the COVID-19 Pandemic (self-quarantine or disruption of study site activities following business continuity plans and/or local government mandates for "stay at home" or "shelter in place"), a safety call to the participant should be made in place of the study site visit. The safety call should encompass all scheduled visit assessments that can be completed remotely, such as assessment for AEs and concomitant medications (eg, as defined in scheduled biweekly safety phone calls). Home visits will be permitted for all non-dosing visits except for Screening if a participant cannot come to the study site as a result of the COVID19 pandemic.

1. Day 0 and OL-D1 visit may be performed on 2 separate visits.
2. Symptom-directed physical examination will be performed at the Participant Decision Visit. On dosing day before injection, the arm receiving the injection should be examined and the associated lymph nodes should be evaluated. Any clinically significant finding identified during a study visit should be reported as a MAAE. Vital signs are to be collected pre- and post-dosing on the days of injection (OL-D1). Participants who are febrile (body temperature ≥ 38.0°C/100.4°F) before dosing (OL-D1) must be rescheduled within the relevant window period to receive the injection. Afebrile participants with minor illnesses can be enrolled at the discretion of the investigator.
3. Pregnancy test at the Participant Decision Visit will be a point-of-care urine test. At the discretion of the investigator, a pregnancy test either via blood or point-of-care urine test can be performed. Follicle-stimulating hormone level may be measured to confirm menopausal status at the discretion of the investigator.
4. Participants with symptoms of COVID-19 will be asked to return within 72 hours or as soon as possible to the study site for an unscheduled visit, to include an NP swab sample (for RT-PCR testing) and other clinical evaluations. If a study site visit is not possible, a home visit may be arranged to collect the NP sample and conduct clinical evaluations. If a home visit is not possible, the participant will be asked to submit a saliva sample to the study site by a Sponsor-approved method. At this visit, NP swab and blood samples will be collected to evaluate for the presence of SARS-CoV-2 infection. NP swab samples will also be tested for the presence of other respiratory pathogens. In addition, the study site may collect an additional respiratory sample for SARS-CoV-2 testing to be able to render appropriate medical care for the study participant as determined by local standards of care. Additionally, clinical information will be carefully collected to evaluate the severity of the clinical case. It is important to note that some of the symptoms of COVID-19 overlap with solicited systemic ARs that are expected after vaccination with mRNA-1273 (eg, myalgia, headache, fever, and chills). During the first 7 days after vaccination, when these solicited ARs are common, investigators should use their clinical judgement to decide if an NP swab should be collected. The collection of an NP swab prior to the Participant Decision Visit can help ensure that cases of COVID-19 are not overlooked. Any study participant reporting respiratory symptoms during the 7-day period after vaccination should be evaluated for COVID-19.
5. Sample must be collected prior to unblinding and injection at the Participant Decision Visit.
6. Post-dosing, participants will have a 30-minute observation period.
7. Trained study personnel will call all participants to collect information relating to any MAAEs (including any signs and symptoms of COVID-19), AEs leading to withdrawal, SAEs, information on concomitant medications associated with those events, and any non-study vaccinations.
8. All concomitant medications relevant to or for the treatment of an SAE or MAAE will be recorded from Screening through the final visit (Day 759).

#### 11.2. APPENDIX 2: Study Governance Considerations

##### 11.2.1. Regulatory and Ethical Considerations

This study will be conducted in accordance with the protocol and with the following:

- Consensus ethical principles derived from international guidelines including the Declaration of Helsinki and Council for International Organizations of Medical Sciences (CIOMS) International Ethical Guidelines.
- Applicable ICH GCP Guidelines.
- Applicable laws and regulatory requirements.
- The protocol, protocol amendments, ICF, Investigator Brochure, and other relevant documents (eg, advertisements) must be submitted to an IRB/IEC by the investigator and reviewed and approved by the IRB/IEC before the study is initiated.
- Any amendments to the protocol will require IRB/IEC approval before implementation of changes made to the study design, except for changes necessary to eliminate an immediate hazard to study participants.
- The investigator will be responsible for the following:

– Providing written summaries of the status of the study to the IRB/IEC annually or more frequently in accordance with the requirements, policies, and procedures established by the IRB/IEC

– Notifying the IRB/IEC of SAEs or other significant safety findings as required by IRB/IEC procedures

– Providing oversight of the conduct of the study at the site and adherence to requirements of 21 CFR, ICH guidelines, the IRB/IEC, European regulation 536/2014 for clinical studies (if applicable), and all other applicable local regulations.

##### 11.2.2. Study Monitoring

Before an investigational site can enter a participant into the study, a representative of Moderna or its representatives will visit the investigational study site to:

- Determine the adequacy of the facilities.
- Discuss with the investigator(s) and other personnel their responsibilities with regard to protocol adherence, and the responsibilities of Moderna or its representatives. This will be documented in a Clinical Study Agreement between Moderna, designated CRO, and the investigator.

According to ICH GCP guideline, the Sponsor of the study is responsible for ensuring the proper conduct of the study with regard to protocol adherence and validity of data recorded on the eCRFs. The study monitor's duties are to aid the investigator and Moderna in the maintenance of complete, accurate, legible, well-organized, and easily retrievable data. The study monitor will advise the investigator of the regulatory necessity for study-related monitoring, audits, IRB/IEC review, and inspection by providing direct access to the source data/documents. In addition, the study monitor will explain to and interpret for the investigator all regulations applicable to the clinical evaluation of an IP as documented in ICH guidelines.

It is the study monitor's responsibility to inspect the eCRFs and source documentation throughout the study to protect the rights of the participants; to verify adherence to the protocol; to verify completeness, accuracy, and consistency of the data; and to confirm adherence of study conduct to any local regulations. Details will be outlined in the Clinical Monitoring Plan. During the study, a monitor from Moderna or a representative will have regular contacts with the investigational site, for the following:

- Provide information and support to the investigator(s).
- Confirm that facilities remain acceptable.
- Confirm that the investigational team is adhering to the protocol, that the data are being accurately recorded in the eCRFs, and that IP accountability checks are being performed.
- Perform source data verification. This includes a comparison of the data in the eCRFs with the participant's medical records at the hospital or practice, and other records relevant to the study. This will require direct access to all original records for each participant (eg, clinical charts or electronic medical record system).
- Record and report any protocol deviations not previously sent.
- Confirm AEs and SAEs have been properly documented on eCRFs and confirm any SAEs have been forwarded to the SAE Hotline, and those SAEs that met criteria for reporting have been forwarded to the IRB/IEC.

The monitor will be available between visits if the investigator(s) or other staff needs information or advice.

In accordance with FDA Guidance on Conduct of Clinical Trials of Medical Products during the COVID-19 Pandemic (FDA March 2020), investigators may convert study site visits to telemedicine visits with the approval of the Sponsor (Section 8 for Procedures).

The DSMB will also have responsibility for safety monitoring (Section 8.4.1).

##### 11.2.3. Audits and Inspections

Moderna, their designee(s), the IRB/IEC, or regulatory authorities will be allowed to conduct site visits to the investigational facilities for the purpose of monitoring or inspecting any aspect of the study. The investigator agrees to allow Moderna, their designee(s), the IRB/IEC, or regulatory authorities to inspect the IP storage area, IP stocks, IP records, participant charts and study source documents, and other records relative to study conduct.

Authorized representatives of Moderna, a regulatory authority, and any IRB/IEC may visit the site to perform audits or inspections, including source data verification. The purpose of a Sponsor audit or inspection is to systematically and independently examine all study-related activities and documents to determine whether these activities were conducted, and data were recorded, analyzed, and accurately reported according to the protocol, ICH GCP E6R2, and any applicable regulatory requirements. The investigator should contact Moderna immediately if contacted by a regulatory agency about an inspection.

The Principal Investigator must obtain IRB approval for the investigation. Initial IRB approval, and all materials approved by the IRB for this study including the participant consent form and recruitment materials must be maintained by the investigator and made available for inspection.

##### 11.2.4. Financial Disclosure

The investigator is required to provide financial disclosure information to allow the Sponsor to submit the complete and accurate certification or disclosure statements required under 21 CFR 54. In addition, the investigator must provide the Sponsor with a commitment to promptly update this information if any relevant changes occur during the course of the investigation and for 1 year following the completion of the study.

The Sponsor, the CRO, and the study site are not financially responsible for further testing or treatment of any medical condition that may be detected during the screening process. In addition, in the absence of specific arrangements, the Sponsor, the CRO, and the study site are not financially responsible for further treatment of the disease under study.

##### 11.2.5. Recruitment Procedures

Advertisements to be used for the recruitment of study participants, and any other written information regarding this study to be provided to the participant should be submitted to the Sponsor for approval. All documents must be approved by the IRB.

##### 11.2.6. Informed Consent Process

The informed consent document(s) must meet the requirements of 21 CFR 50, local regulations, ICH guidelines, Health Insurance Portability and Accountability Act (HIPAA) requirements, where applicable, and the IRB/IEC or study center. All consent documents will be approved by the appropriate IRB/IEC. The actual ICF used at each center may differ, depending on local regulations and IEC / IRB requirements. However, all versions must contain the standard information found in the sample ICF provided by the Sponsor. Any change to the content of the ICF must be approved by the Sponsor and the IEC / IRB prior to the form being used.

If new information becomes available that may be relevant to the participant's willingness to continue participation in the study, this will be communicated to him / her in a timely manner. Such information will be provided via a revised ICF or an addendum to the original ICF.

The investigator or his/her representative will explain the nature of the study to the participant and answer all questions regarding the study.

The investigator is responsible for ensuring that the participant fully understands the nature and purpose of the study. Information should be given in both oral and written form whenever possible.

No participant should be obliged to participate in the study. The participant must be informed that participation is voluntary. Participants, their relatives, guardians, or (if applicable) legal representatives must be given ample opportunity to inquire about details of the study. The information must make clear that refusal to participate in the study or withdrawal from the study at any stage is without any prejudice to the participant's subsequent care.

The participant must be allowed sufficient time to decide whether they wish to participate. The participant must be made aware of and give consent to direct access to his/her source medical records by study monitors, auditors, the IRB/IEC, and regulatory authorities. The participant should be informed that such access will not violate participant confidentiality or any applicable regulations. The participant should also be informed that he/she is authorizing such access by signing the ICF.

A copy of the ICF(s) must be provided to the participant.

A participant who is rescreened is not required to sign another ICF if the rescreening occurs within 28 days from the previous ICF signature date.

The ICF will also explain that excess serum from immunogenicity testing may be used for future research, which may be performed at the discretion of the Sponsor to further characterize the immune response to SARS-CoV-2, additional assay development, and the immune response across CoV.

##### 11.2.7. Protocol Amendments

No change or amendment to this protocol may be made by the investigator or the Sponsor after the protocol has been agreed to and signed by all parties unless such change(s) or amendment(s) has (have) been agreed upon by the investigator or the Sponsor. Any change agreed upon will be recorded in writing, and the written amendment will be signed by the investigator and the Sponsor. Institutional review board approval is required prior to the implementation of an amendment, unless overriding safety reasons warrant immediate action, in which case the IRB(s)/IEC(s) will be promptly notified.

Any modifications to the protocol or the ICF, which may impact the conduct of the study, potential benefit of the study, or may affect participant safety, including changes of study objectives, study design, participant population, sample sizes, study procedures, or significant administrative aspects will require a formal amendment to the protocol. Such amendment will be released by the Sponsor, agreed by the investigator(s), and approved by the relevant IRB(s)/IEC(s) prior to implementation. A signed and dated statement that the protocol, any subsequent relevant amended documents and the ICF have been approved by relevant IRB(s)/IEC(s) must be provided to the Sponsor before the study is initiated.

Administrative changes of the protocol are minor corrections and/or clarifications that have no effect on the way the study is to be conducted. These administrative changes will be released by the Sponsor, agreed by the investigator(s), and notified to the IRB(s)/IEC(s).

##### 11.2.8. Protocol Deviations

The noncompliance may be either on the part of the participant, the investigator, or the study site staff. As a result of deviations, corrective actions are to be developed by the site and implemented promptly.

It is the responsibility of the site investigator to use continuous vigilance to identify and report deviations to the Sponsor or its designee. All deviations must be addressed in study source documents, reported to study monitor. Protocol deviations must be sent to the reviewing IRB/IEC per their policies. The site investigator is responsible for knowing and adhering to the reviewing IRB/IEC requirements.

##### 11.2.9. Data Protection

Participants will be assigned a unique identifier by the Sponsor. Any participant records or datasets that are transferred to the Sponsor will contain the identifier only; participant names or any information which would make the participant identifiable will not be transferred.

The participant must be informed that his/her personal study-related data will be used by the Sponsor in accordance with local data protection law. The level of disclosure must also be explained to the participant.

The participant must be informed that his/her medical records may be examined by Clinical Quality Assurance auditors or other authorized personnel appointed by the Sponsor, by appropriate IRB/IEC members, and by inspectors from regulatory authorities.

Individual participant medical information obtained as a result of this study is considered confidential, and disclosure to third parties is prohibited. Information will be accessible to authorized parties or personnel only. Medical information may be given to the participant's physician or to other appropriate medical personnel responsible for the participant's well-being. Each participant will be asked to complete a form allowing the investigator to notify the participant's primary health care provider of his/her participation in this study.

All laboratory specimens, evaluation forms, reports, and other records will be identified in a manner designed to maintain participant confidentiality. All records will be kept in a secure storage area with limited access. Clinical information will not be released without the written permission of the participant, except as necessary for monitoring and auditing by the Sponsor, its designee, relevant regulatory authority, or the IRB.

The investigator and all employees and coworkers involved with this study may not disclose or use for any purpose other than performance of the study, any data, record, or other unpublished, confidential information disclosed to those individuals for the purpose of the study. Prior written agreement from the Sponsor or its designee must be obtained for the disclosure of any confidential information to other parties.

##### 11.2.10. Sample Retention and Future Biomedical Research

The retention period of laboratory samples will be 15 years, or as permitted by local regulations, to address further scientific questions related to mRNA-1273 or anti-respiratory virus immune response. In addition, identifiable samples can be destroyed at any time at the request of the participant. During the study, or during the retention period, in addition to the analysis outlined in the study endpoints, exploratory analysis may be conducted, using other antibody-based methodologies, on any remaining blood or serum samples, including participants who provide samples for screening, but are not subsequently enrolled. These analyses would extend the search for other potentially relevant biomarkers to investigate the effect of mRNA-1273, as well as to determine how changes in the markers may relate to exposure and clinical outcomes. A decision to perform such exploratory research may arise from new scientific findings related to the drug class or disease, as well as reagent and assay availability.

##### 11.2.11. Data Quality Assurance and Quality Control

Data collection is the responsibility of the clinical study staff at the site under the supervision of the site investigator. The investigator is responsible for ensuring the accuracy, completeness, legibility, and timeliness of the data reported.

- All participant data relating to the study will be recorded eCRF unless transmitted to the Sponsor or designee electronically (eg, laboratory data). The investigator is responsible for verifying that data entries are accurate and correct by physically or electronically signing the eCRF.
- The investigator must maintain accurate documentation (source data) that supports the information entered in the eCRF.
- The investigator must permit study-related monitoring, audits, IRB/IEC review, and regulatory agency inspections and provide direct access to source data documents.
- Monitoring details describing strategy (eg, risk-based initiatives in operations and quality such as Risk Management and Mitigation Strategies and Analytical Risk-Based Monitoring), methods, responsibilities and requirements, including handling of noncompliance issues and monitoring techniques (central, remote, or on-site monitoring) are provided in the study Monitoring Plan, Centralized Monitoring Plan, and Risk Management Plan.
- The Sponsor or designee is responsible for the data management of this study including quality checking of the data.
- The Sponsor assumes accountability for actions delegated to other individuals (eg, Contract Research Organizations).
- Study monitors will perform ongoing source data verification to confirm that data entered into the CRF by authorized site personnel are accurate, complete, and verifiable from source documents; that the safety and rights of participants are being protected; and that the study is being conducted in accordance with the currently approved protocol and any other study agreements, ICH GCP, and all applicable regulatory requirements.
- Records and documents, including signed ICFs, pertaining to the conduct of this study must be retained by the investigator for 15 years after study completion unless local regulations or institutional policies require a longer retention period. No records may be destroyed during the retention period without the written approval of the Sponsor. No records may be transferred to another location or party without written notification to the Sponsor.

Quality assurance (QA) includes all the planned and systematic actions that are established to ensure that the clinical study is performed, and the data are generated, documented (recorded), and reported according to ICH GCP and local/regional regulatory standards.

A QA representative from the Sponsor or qualified designee, who is independent of and separated from routine monitoring, may periodically arrange inspections/audits of the clinical study by reviewing the data obtained and procedural aspects. These inspections may include on-site inspections/audits and source data checks. Direct access to source documents is required for the purpose of these periodic inspections/audits.

##### 11.2.12. Data Collection and Management

This study will be conducted in compliance with ICH CGP guidelines. This study will also be conducted in accordance with the most recent version of the Declaration of Helsinki.

This study will use electronic data collection (to collect data directly from the investigational site using eCRFs). The investigator is responsible for ensuring that all sections of each eCRF are completed promptly and correctly and that entries can be verified against any source data.

Study monitors will perform source document verification to identify inconsistencies between the eCRFs and source documents. Discrepancies will be resolved in accordance with the principles of GCP. Detailed study monitoring procedures are provided in the Clinical Monitoring Plan.

AEs will be coded with the most current available version of MedDRA. Concomitant medications will be coded using WHO – Drug Reference List.

##### 11.2.13. Source Documents

Source documents are original documents or certified copies, and include, but are not limited to, diaries, medical and hospital records, screening logs, informed consent / assent forms, telephone contact logs, and worksheets. Source documents provide evidence for the existence of the participant and substantiate the integrity of the data collected. Source documents are filed at the investigator's site.

Data reported on the CRF or entered in the eCRF that are transcribed from source documents must be consistent with the source documents or the discrepancies must be explained. The investigator may need to request previous medical records or transfer records, depending on the study. Also, current medical records must be available.

Moderna or its designee requires that the investigator prepare and maintain adequate and accurate records for each participant treated with the IP. Source documents such as any hospital, clinic, or office charts and the signed ICFs are to be included in the investigator's files with the participant's study records.

##### 11.2.14. Retention of Records

The Principal Investigator must maintain all documentation relating to the study for a period of at least 2 years after the last marketing application approval or, if not approved, 2 years following the discontinuance of the test article for investigation. If this requirement differs from any local regulations, the local regulations will take precedence unless the local retention policy is less than 2 years.

If it becomes necessary for Moderna or the regulatory authority to review any documentation relating to the study, the investigator must permit access to such records. No records will be destroyed without the written consent of the Sponsor, if applicable. It is the responsibility of the Sponsor to inform the investigator when these documents no longer need to be retained.

##### 11.2.15. Study and Site Closure

If the study is prematurely terminated or suspended, the Sponsor shall promptly inform the investigators, the IECs/IRBs, the regulatory authorities, and any CRO(s) used in the study of the reason for termination or suspension, as specified by the applicable regulatory requirements. The investigator shall promptly inform the participant and should assure appropriate participant therapy and/or follow-up.

The Sponsor or designee reserves the right to close the study site or terminate the study at any time for any reason at the sole discretion of the Sponsor.

The investigator may initiate study-site closure at any time, provided there is reasonable cause and sufficient notice is given in advance of the intended termination.

Reasons for the early closure of a study site by the Sponsor or investigator may include but are not limited to:

- Continuation of the study represents a significant medical risk to participants.
- Failure of the investigator to comply with the protocol, the requirements of the IRB/IEC or local health authorities, the Sponsor's procedures, or GCP guidelines.
- Inadequate recruitment of participants by the investigator.
- Discontinuation of further mRNA-1273 development.

Study sites will be closed upon study completion. A study site is considered closed when all required documents and study supplies have been collected and a study-site closure visit has been performed.

##### 11.2.16. Publication Policy

The results of this study may be published or presented at scientific meetings. If this is foreseen, the investigator agrees to submit all manuscripts or abstracts to the Sponsor before submission. This allows the Sponsor to protect proprietary information and to provide comments.

The Sponsor will comply with the requirements for publication of study results. In accordance with standard editorial and ethical practice, the Sponsor will generally support publication of multicenter studies only in their entirety and not as individual site data. In this case, a coordinating investigator will be designated by mutual agreement.

Authorship will be determined by mutual agreement and in line with International Committee of Medical Journal Editors authorship requirements.

The clinical study plan and the results of the study will be published on www.ClinicalTrials.gov in accordance with 21 CFR 50.25(c). The results of and data from this study belong to Moderna.

#### 11.3. APPENDIX 3: Contraceptive Guidance

##### Woman of Childbearing Potential (WOCBP)

Females of childbearing potential are those who are considered fertile following menarche and until becoming postmenopausal unless permanently sterile (see below). If fertility is unclear (eg, amenorrhea in adolescents or athletes) and a menstrual cycle cannot be confirmed before first dose of study treatment, additional evaluation should be considered.

Women in the following categories are not considered WOCBP:

1. Premenarchal
2. Premenopausal, surgically sterile female with 1 of the following:
    a. Documented complete hysterectomy
    b. Documented bilateral salpingectomy
    c. Documented bilateral oophorectomy

For individuals with permanent infertility due to an alternate medical cause other than the above, (eg, mullerian agenesis, androgen insensitivity), investigator discretion should be applied in determining study entry.

Note: Documentation can come from the site personnel's: review of the participant's medical records, medical examination, or medical history interview.

3. Postmenopausal female

- A postmenopausal state is defined as no menses for 12 months without an alternative medical cause. The following age-specific requirements apply:
    - Women <50 years of age would be considered postmenopausal if they have been amenorrheic for 12 months or more following cessation of exogenous hormonal treatments and if they have luteinizing hormone and FSH levels in the postmenopausal range for the institution.
    - Women ≥ 50 years of age would be considered postmenopausal if they have been amenorrheic for 12 months or more, had radiation-induced menopause with last menses >1 year ago or had chemotherapy-induced menopause with last menses > 1 year ago.
- A high FSH level in the postmenopausal range may be used to confirm a postmenopausal state in women not using hormonal replacement therapy (HRT).
- Females on HRT and whose menopausal status is in doubt will be required to use one of the non-estrogen hormonal highly effective contraception methods if they wish to continue their HRT during the study. Otherwise, they must discontinue HRT to allow confirmation of postmenopausal status before study enrollment.

### 11.4. APPENDIX 4: Statistical Appendices

#### 11.4.1. Estimands and Estimand Specifications

**Table 23: Intercurrent Event Types**

| Label | Intercurrent Event Type | Comment |
|---|---|---|
| IcEv1 (death without confirmation of cases, ie, unrelated death) | Unrelated death without documented confirmed COVID-19 | Participants in PP Set who die due to reasons unrelated to COVID-19 without confirmation of being a case will all be included in statistical analysis. |
| IcEv2 (early COVID-19) | COVID-19 starting up to 14 days after the second dose of IP | Participants in PP Set who experience early COVID-19 up to 14 days after the second dose of IP will all be included in statistical analysis. |

Abbreviations: IcEv: intercurrent event; IP = investigational product; PP: per-protocol.

**Table 24: Primary Objective and Estimands with Rationale for Strategies to Address Intercurrent Events for Per-Protocol Analysis**

| **Objective: To demonstrate the efficacy of mRNA-1273 to prevent COVID-19** | |
|---|---|
| **Estimand Description** | Vaccine efficacy will be measured using 1 – HR (mRNA-1273/Placebo) of COVID-19 from 14 days after second dose of IP in adults. A hypothetical strategy will be used for deaths unrelated to COVID-19, and a hypothetical strategy will also be used for early COVID-19 in participants in the PP Set. |
| **Target Population** | Adults aged 18 years and older in circumstances at a high risk of SARS-CoV-2 infection but without medical conditions that pose additional risk of developing severe disease.<br>The population excludes those previously infected or vaccinated for SARS-CoV-2 or with a medical condition, on treatment that poses additional risks (including those requiring immunosuppressants or immune-modifying drugs), or SARS-CoV-2 pre-positive. |
| **Variable/Endpoint** | Time to COVID-19 disease, censoring at early discontinuation, early COVID-19, or last assessment for an event not being observed, whichever comes earlier. |
| **Treatment Condition(s)** | **Test:** mRNA-1273<br>**Reference:** Placebo |
| **Estimand Label** | **Estimand 1** |
| **Population-Level Summary** | Vaccine efficacy defined as 1 - HR of mRNA-1273/Placebo |
| **Intercurrent Event Strategy** | |
| **IcEv1 (Unrelated death):** | Hypothetical |
| **IcEv2 (early infection):** | Hypothetical |
| **Rationale for Strategy(s)** | Hypothetical: unrelated death without confirmation of COVID-19 will be censored at the time of death as if there is no event, handled with independent censoring.<br>Hypothetical: early case in PP Set will be censored at the time of case onset if it is not a case, handled with independent censoring. |

##### 11.4.2. Statistical Methods and Sensitivity Analyses

**Table 25: Summary of Statistical Methods and Sensitivity Analyses**

| Estimand Label | Estimand Description | Main Estimation | | | Sensitivity Analysis |
|---|---|---|---|---|---|
| | | Analysis Set | Imputation/Data/Censoring Rules | Analysis Model/Method | |
| Estimand 1 | See Table 24 | PP | Participants who did not develop COVID-19, ongoing or complete the study will be censored at the last assessment.<br>Participants who discontinue early or die without COVID-19 will be censored at the date of discontinuation or death respectively.<br>Participants who is a confirmed case prior to 14 days after the second dose of IP will be censored on the date of confirmation. | The (1 - HR) with 95% CI will be estimated using Cox proportional hazard regression analysis with treatment arm as a fixed effect stratified by randomization stratification factors. And the null hypothesis will be tested using Wald Chi-square method. | As described in Section 9.5.1 with cases counted starting after the second dose of IP |

#### 11.5. APPENDIX 5: Protocol Amendment History

The document history table for this protocol and the Protocol Amendment Summary of Changes Table for the current Amendment 8 is located directly before the Table of Contents.

A description of Amendment 7, Amendment 6, Amendment 5, Amendment 4, Amendment 3, Amendment 2, and Amendment 1 is presented in this appendix.

**Amendment 7, 10 Feb 2021**

**Main Rationale for the Amendment:**

The purpose of this amendment is to collect safety information on suspected cases of anaphylaxis and to include participant history of facial injections or dermal fillers in the eDiary.

The summary of changes table provided here describes the major changes made in Amendment 7 relative to Amendment 6, including the sections modified and the corresponding rationales. The synopsis of Amendment 7 has been modified to correspond to changes in the body of the protocol.

**Summary of Major Changes from Protocol Amendment 6 to Protocol Amendment 7:**

| Section # and Name | Description of Change | Brief Rationale |
|---|---|---|
| Title Page, Protocol Approval Page, Headers, Protocol Amendment Summary of Changes | Updated the protocol version and date. | To reflect the new version and date of the protocol. |
| Sections 1.1 (Synopsis), 6.2.2 (Administration of Investigational Product), 6.2.8.1 (Planned Unblinding) | Clarified that study site personnel who were blinded during the Blinded Phase will be unblinded **at the participant level** at the Participant Decision clinic visit. | To clarify that the site personnel is being unblinded only at the Participant Decision clinic visit. |
| Sections 1.1 (Synopsis), 4.1.2 (Part B, the Open-label Observational Phase) | Clarified that with regards to EUA eligibility in Part B, the CDC-EUA guidance specifications will supersede eligibility criteria in the protocol. | To clarify that the CDC-EUA guidance specifications supersede protocol specified criteria for EUA eligibility in Part B. |
| Section 4.1 (General Design) | Corrected the number of scheduled clinic visits and added mention of the Participant Decision Visit. | To clarify the number of scheduled visits. |
| Section 4.1.2 (Part B, the Open-Label Observational Phase) | Updated text to clarify requirements for Part B subjects who are unblinded and received only 1 dose of mRNA-1273 during Part A. | Clarification of requirements for Part B subjects who received only 1 dose of mRNA-1273 during Part A. |

| Section # and Name | Description of Change | Brief Rationale |
|---|---|---|
| Section 6.2.4 (Packaging and Labeling) | Updated to indicate that vaccine product is packaged in a 10R glass vial with a 6-dose, 5.0-mL or 10-dose, 6.3-mL fill volume. | To include IP presentation for additional supplies that may be used in Part B of the study. |
| Section 6.2.5 (Storage) | Updated to specify storage requirements for the 6-dose and 10-dose mRNA vaccine. | To include IP storage conditions for additional supplies that may be used in Part B of the study. |
| Sections 6.4.2 (Concomitant Medications and Therapies), 7.2 (Discontinuation of Study Treatment), 8.2.2 (Use of Electronic Diaries), 8.2.6 (Blood Sampling Volumes, Table 4) | Clarified requirements for Part A and Part B. | Clarification of concomitant medication, eDiary, and blood sampling procedures during Part A and Part B. |
| Section 8.2.2 (Use of Electronic Diaries), Appendix 1 (Schedule of Events, Table 17 footnote 7). | Clarified that eDiary prompts to surveil for weekly COVID-19 symptoms will be performed for all participants in Part A and Part B.<br><br>Added eDiary prompt to solicit collection of participant history of facial injections or dermal fillers. | Clarification.<br><br>To assure all concomitant medications are recorded. |
| Section 8.2.4 (Physical Examination) | Removed body mass index from the Participant Decision Visit. | Clarification. |
| Section 8.2.6 (Blood Sampling Volumes) | Updated maximum planned volume of blood collection for the complete study to include a range. | Clarification. |
| Section 8.3.2 (Medically Attended Adverse Events) | Added suspected cases of anaphylaxis as an MAAE and requirement to report as an SAE. | Sponsor collecting anaphylaxis information. |
| Section 9.4 (Analysis Populations, Table 8); Synopsis | Clarified Immunogenicity Subset | Clarification |
| Section 9.5.1.1 (Efficacy Analysis on Primary Endpoint), Appendix 4 (Statistical Appendices, Table 23 and Table 24) | Updated intercurrent events and estimands. | Clarification of estimand. |
| Section 9.5.1.3 (Long-term Efficacy Analysis); Synopsis | Updated case count parameters. | Provide long-term efficacy analysis strategy relevant to change in study design. |

| Section # and Name | Description of Change | Brief Rationale |
|---|---|---|
| Appendix 1 (Schedule of Events, Table 21 footnote 2, and Table 22 footnote 2) | Updated physical examination to symptom-directed physical examination and removed height and weight requirements from the Open-Label Clinic Visits. | Clarification |

**Amendment 6, 23 Dec 2020**

**Main Rationale for the Amendment:**

The purpose of this amendment is to inform all ongoing study participants of the availability of and eligibility criteria of any COVID-19 vaccine made available under an EUA and to offer participants who originally received placebo in this study the potential benefit of vaccination against COVID-19, given that the primary efficacy endpoint for mRNA-1273 against COVID-19 was met per the protocol-defined IA.

The summary of changes table provided here describes the major changes made in Amendment 6 relative to Amendment 5, including the sections modified and the corresponding rationales. The synopsis of Amendment 6 has been modified to correspond to changes in the body of the protocol.

**Summary of Major Changes from Protocol Amendment 5 to Protocol Amendment 6:**

| Section # and Name | Description of Change | Brief Rationale |
|---|---|---|
| Title Page, Protocol Approval Page, Headers, Protocol Amendment Summary of Changes | Updated the protocol version and date | To reflect the new version and date of the protocol |
| Sections 1.1 (Synopsis), 1.2 (Schema), 4.1.2 (Part B, the Open-Label Observational Phase), 6.2.8.1 (Planned Unblinding), 7.3.1 (Participant Withdrawal), and 11.1 (Schedules of Events) | Added a "Participant Decision clinic visit". | This visit provides the opportunity for study site personnel to discuss with and offer to participants, the choice to be unblinded, as well as offering to participants who originally received placebo, the choice to receive active vaccination with mRNA-1273 and possible vaccination against COVID-19. Participants will also sign a revised Informed Consent Form at this visit. |

| Section # and Name | Description of Change | Brief Rationale |
|---|---|---|
| Sections 1.1 (Synopsis), 1.2 (Schema), 4.1 (General Design), 4.2 (Scientific Rationale for Study), 6.2 (Method of Randomly Assigning Participants to Treatment Groups [Part A], Blinded Phase Only), and 11.1 (Schedules of Events) | Changes to study design that split the study into a Blinded Phase and an Open-Label Observational Phase. | These changes accommodate the breaking of the blind and offering of open-label vaccination to all participants. This change also distinguishes the Open-Label Supplemental Schedule of Events (Vaccination Phase) for participants who received placebo, and who meet EUA eligibility, and request to receive active vaccine. |
| Sections 1.1 (Synopsis), 1.2 (Schema), 4.1.2 (Part B, the Open-Label Observational Phase), and 11.1 (Schedules of Events) | Added a Supplemental SoE and a Modified Supplemental SoE | These provide paths for participants who choose to be unblinded and receive mRNA-1273, and for participants who previously received only 1 dose of blinded mRNA-1273 during the study (Blinded Phase), to transition into the Open-Label Phase of the study. |
| Section 2.2.2 (Clinical Studies) | Updated status of ongoing clinical studies, including this study (mRNA-1273-P301). | The status of the 3 clinical studies (one Phase 1, one Phase 2a, and this Phase 3 study) have changed since Amendment 5. In addition, the results of the interim analyses in this study of the primary efficacy endpoint (prevention of COVID-19 infection), a major secondary endpoint (prevention of severe COVID-19), and safety and reactogenicity endpoints are now available and are provided here. These results provide the justification for offering participants the opportunity to receive active IP (mRNA-1273) and the potential benefit of vaccination against COVID-19. |

| Section # and Name | Description of Change | Brief Rationale |
|---|---|---|
| Section 1.1 (Synopsis) and 9.5.1.3 (Long-term Efficacy Analysis) | Addition of long-term efficacy analyses | Allows for the analysis of the long-term efficacy of mRNA-1273 in the following treatment cohorts:<br>• mRNA-1273 Cohort: Participants randomized to mRNA-1273 in the Blinded Phase.<br>• Placebo Cohort: Participants randomized to Placebo in the Blinded Phase and did not crossover to mRNA-1273 in the Open-label Phase.<br>• Placebo-mRNA-1273 Cohort: Participants randomized to Placebo in the Blinded phase and crossed over to mRNA-1273 in the Open-label Phase. |
| Section 2.3.3 (Overall Benefit/Risk Conclusion) | Updated the Benefit/Risk Assessment | Based on the interim results from this pivotal Phase 3 study, mRNA-1273 prevents COVID-19 and severe COVID-19. The demonstrated clinical benefit of mRNA-1273 is supported by evidence of a robust immune response both in terms of binding antibodies (bAbs) and neutralizing antibodies (nAbs) as well as the induction of CD4+ T-cells with a Th-1 dominant phenotype. |
| Section 11.1 | Added footnote to Table 16 | To clarify that the eDiary Safety Prompts will be triggering off on Day 61 to take into consideration the (-3 day) window allowance on Day 29. |

**Amendment 5, 11 Nov 2020**

**Main Rationale for the Amendment:**

The main purpose of this amendment is to clarify that the eDiary prompts for safety surveillance will be weekly and to add Month 19 safety call.

The summary of changes table provided here describes the major changes made in Amendment 5 relative to Amendment 4, including the sections modified and the corresponding rationales. The synopsis of Amendment 5 has been modified to correspond to changes in the body of the protocol.

**Summary of Major Changes from Protocol Amendment 4 to Protocol Amendment 5:**

| Section # and Name | Description of Change | Brief Rationale |
|---|---|---|
| Title Page, Protocol Approval Page, Headers, Protocol Amendment Summary of Changes | Updated the protocol version and date | To reflect the new version and date of the protocol |
| Section 11.1 | Updated Appendix 1 (Tables 16 and 17) | To clarify the weekly schedule for eDiary prompts through Year 1 and Year 2 of follow-up |
| Section 11.1 | Added Month 19 Safety Call | To clarify that participants will have continuous weekly (eDiary prompts) and monthly (safety calls) through Year 1 and Year 2 of follow-up |

**Amendment 4, 30 Sep 2020**

**Main Rationale for the Amendment:**

The main purpose of this amendment is to increase the upper limit for stratification of enrolled participants considered "at risk" at Screening to 50%.

The summary of changes table provided here describes the major changes made in Amendment 4 relative to Amendment 3, including the sections modified and the corresponding rationales. The synopsis of Amendment 4 has been modified to correspond to changes in the body of the protocol.

**Summary of Major Changes from Protocol Amendment 3 to Protocol Amendment 4:**

| Section # and Name | Description of Change | Brief Rationale |
|---|---|---|
| Title Page, Protocol Approval Page, Headers, Protocol Amendment Summary of Changes | Updated the protocol version and date | To reflect the new version and date of the protocol |
| Section 6.2.1.1 (Stratification) | Increased the upper limit for stratification of enrolled participants considered "at risk" at Screening to up to 50%, from 40%. | To enhance the diversity of the study population by increasing the number of racial and ethnic minority participants in the study. as participants from these communities often have higher rates of comorbidities. |
| Section 9.5.1.1 (Efficacy Analysis on Primary Endpoint) and Section 11.4.1 (Estimands and Estimand Specifications) | Updated the description of an intercurrent event (unrelated death) and its strategy | To align with the description in the Statistical Analysis Plan. |

**Amendment 3, 20 Aug 2020**

**Main Rationale for the Amendment:**

The main purpose of this amendment is to make changes to the protocol in response to feedback from CBER.

The summary of changes table provided here describes the major changes made in Amendment 3 relative to Amendment 2, including the sections modified and the corresponding rationales. Minor editorial or formatting changes are not included in this summary table. The synopsis of Amendment 3 has been modified to correspond to changes in the body of the protocol.

**Summary of Major Changes from Protocol Amendment 2 to Protocol Amendment 3:**

| Section # and Name | Description of Change | Brief Rationale |
|---|---|---|
| Title Page, Protocol Approval Page, Headers, Protocol Amendment Summary of Changes | Updated the protocol version and date | To reflect the new version and date of the protocol |
| Section 5 (Study Population) | Added a sentence to describe the intent to enroll a representative sample of racial and ethnic minority participants in the study | To enhance the diversity of the study population |
| Section 5.2 (Exclusion Criteria) | Added clarification to exclusion criterion #11 to define the parameters based on screening CD4 count and viral load for exclusion of study participants | To clarify the definition of controlled HIV disease in the exclusion criterion such that only participants with well-controlled HIV disease are enrolled in the study |

| Section # and Name | Description of Change | Brief Rationale |
|---|---|---|
| Section 5.2 (Exclusion Criteria) | Removed "topical tacrolimus" from exclusion criterion #12 | No evidence to support any systemic effect of topical tacrolimus to warrant excluding them |
| Section 6.2.1.1 (Stratification) | Added HIV infection to the risk factors at Screening | To stratify participants based on certain risk factors |
| Section 8.2.3 (Demographics/Medical History) | Added collection of risk factors for complications of COVID-19 | To document the diagnosis of any risk factor for complications of COVID19 used for stratification |
| Section 9.3 (Sample Size Determination) | Removed redundant bullet | To remove redundancy in the assumptions listed |
| Section 9.5.2 (Safety Analyses) | Removed safety analysis by serostatus | No added value for this analysis. Other subgroup analyses may be specified in the SAP, as needed. |
| Section 9.5.5 (Subgroup Analyses) | Removed the categories (white, non-white) from the Race Variable | To allow for more refined Race categorization being collected in the eCRF |
| Appendix 11.3 | Removed "cessation of exogenous hormonal therapy" | To allow post-menopausal women to take these medications if needed for the treatment of the symptoms of menopause |
| Appendix 11.3 | Removed "using hormonal contraception" from postmenopausal female with high FSH levels | Not a standard of care for women for treatment of menopausal symptoms |

**Amendment 2, 31 Jul 2020:**

**Main Rationale for the Amendment:**

The main purpose of this amendment is to provide more intensive surveillance of symptoms and severity of cases of COVID-19 after the first dose of investigational product.

The summary of changes table provided here describes the major changes made in Amendment 2 relative to Amendment 1, including the sections modified and the corresponding rationales. Minor editorial or formatting changes are not included in this summary table. The synopsis of Amendment 2 has been modified to correspond to changes in the body of the protocol.

**Summary of Major Changes from Protocol Amendment 1 to Protocol Amendment 2:**

| Section # and Name | Description of Change | Brief Rationale |
|---|---|---|
| Title Page, Protocol Approval Page, Headers, Protocol Amendment Summary of Changes | Updated the protocol version and date | Reflect the new version and date of the protocol |

| Section # and Name | Description of Change | Brief Rationale |
|---|---|---|
| Section 4.1 (General Design), Section 7.2 (Discontinuation of Study Treatment), Section 8.1.1 (Efficacy Assessments Related to COVID-19 and SARS-CoV-2 Infection), Section 8.1.2 (Surveillance for COVID-19 Symptoms),Table 14 (Schedule of Events [Vaccination Phase, Day 1 – Day 57]) | Added a Day 29 NP swab prior to Dose 2 | Improve surveillance for asymptomatic infection prior to Dose 2 to assist in discriminating COVID-19 symptoms from solicited systemic reactions after vaccination |
| Section 4.1 (General Design), Table 15 (Schedule of Events [Surveillance Phase, Day 64 – Day 394]) | Changed the clinic visit at Month 4 to a safety call | Improve participant safety and adherence to protocol |
| Section 4.1 (General Design), Section 8.1.2 (Surveillance for COVID-19 Symptoms), Table 14 Footnote 5 (Schedule of Events [Vaccination Phase, Day 1 – Day 57]) | Clarified symptom duration (> 48 hours) to trigger NP swab collection and investigator judgement whether to obtain an NP swab in the 7 days following vaccination due to overlap of solicited systemic symptoms and COVID-19 | Improve surveillance for cases of COVID-19 |
| Section 4.1 (General Design), Section 8.1.3 (Convalescent Period Starting with the Illness Visit), Section 8.1.4 (Ancillary Supplies for Participant Use) | Removed mention of continuous biometric monitoring | Simplify data management and clinical operations |
| Section 5.1 (Inclusion Criteria), Section | Removed the inclusion criterion regarding male contraception | Requirement not generally applicable for a Phase 3 vaccine study |
| Section 5.2 (Exclusion Criteria), Section 6.4.3 (Concomitant Medications and Vaccines that May Lead to the Elimination of a Participant from Per-Protocol Analyses) | Clarified language around influenza vaccination | Make restrictions on influenza vaccination less restrictive than for other licensed vaccines relative to administration of investigational product |
| Section 5.2 (Exclusion Criteria) | Removed restriction on enrollment of participants with human immunodeficiency virus (HIV) infection | Participants on stable antiretroviral therapy are not excluded, which diversifies the participant group |
| Section 8.3.7 (Time Period and Frequency for Collecting AE and SAE Information), Section 8.3.10 (Reporting Adverse Events) | Added expedited reporting of confirmed COVID-19 cases | Improve surveillance for cases of COVID-19 |

| Section # and Name | Description of Change | Brief Rationale |
|---|---|---|
| Section 8.4.2 (Data and Safety Monitoring Board) | Added an Oversight Group | Clarify responsibility for declaring early efficacy or for taking action to stop, pause, or continue the study |
| Section 8.4.2 (Data and Safety Monitoring Board) | Updated description of monitoring for potential harm, including details on case counting for the purpose of harm monitoring to align with a DSMB analysis plan | Clarify description for harm monitoring, based on FDA feedback |

**Amendment 1, 26 Jun 2020:**

**Main Rationale for the Amendment:**

The main purpose of this amendment is to provide more intensive surveillance of symptoms and severity of cases of COVID-19 after the first dose of investigational product.

The summary of changes table provided here describes the major changes made in Amendment 1 relative to the original protocol, including the sections modified and the corresponding rationales. Minor editorial or formatting changes are not included in this summary table.

**Summary of Major Changes in Protocol Amendment 1:**

| Section # and Name | Description of Change | Brief Rationale |
|---|---|---|
| Title page, Signature page, and header | Updated the protocol version and date | Reflect the new version and date of the protocol |
| Synopsis; Section 4.1, General Design; Section 8.1.3, Convalescent Period; Section 11.1, Schedules of Events, Table 17 | Increased the frequency of telemedicine contacts during the Convalescent Period following the Initial Illness Visit | Improve the amount and quality of COVID-19 symptom data collected during the Convalescent Period |
| Synopsis; Section 4.1, General Design; Section 8.1.3, Convalescent Period; Section 8.1.4, Ancillary Supplies for Participant Use; Section 11.1, Schedules of Events, Table 17 | Added monitoring of oxygen saturation to the Convalescent Period | Improve surveillance for incidence of COVID-19 during the study |

| Section # and Name | Description of Change | Brief Rationale |
|---|---|---|
| Synopsis; Section 8.1.3, Convalescent Period; Section 8.1.4, Ancillary Supplies for Participant Use; Section 11.1, Schedules of Events, Table 17 | Increased the frequency of monitoring for SARS-CoV-2, using saliva as the preferred sample matrix after the Illness Visit during the Convalescent Period | Improve the sensitivity of monitoring the time course of viral shedding during COVID-19 |
| Synopsis; Section 3, Objectives and Endpoints | Added a respiratory sample for hospitalized participants as a matrix for confirming the presence of SARS-CoV-2 | Increase the potential number of evaluable COVID-19 cases |
| Synopsis; Section 3, Objectives and Endpoints | Broadened the definition for seroconversion at a participant level | Included neutralizing antibody (nAb) in addition to binding antibody (bAb) |
| Section 8, Study Assessments and Procedures; Section 8.1.3, Convalescent Period; Section 8.1.4, Ancillary Supplies for Participant Use; Section 8.2.2, Use of Electronic Diaries; Section 11.1, Schedules of Events, Table 17 | Eliminated paper diaries, substituting an instruction card listing symptoms and a severity grading system to enhance the quality of data obtained by telemedicine contacts | Reduce fomite transmission of SARS-CoV-2 and increase frequency of investigative staff interactions with participants during the Convalescent Period |
| Section 8.1.2, Surveillance for COVID-19 Symptoms | Decreased the number of symptoms (to one of the 11 CDC symptoms) that would result in an Illness Visit | Increase the likelihood of capturing all COVID-19 cases in the earliest stage of disease |
| Section 8.2.2, Use of Electronic Diaries | Expanded the scope of eDiary prompts and data collected during the Surveillance Phase | Increase the likelihood of capturing all COVID-19 cases in the earliest stage of disease |

Signature Page for VV-CLIN-002035 v3.0

| | |
|---|---|
| Approval | Allison August<br>Medical<br>29-Mar-2021 14:17:57 GMT+0000 |

Signature Page for VV-CLIN-002035 v3.0
